# Genetic analyses identify shared genetic components related to autoimmune and cardiovascular diseases

**DOI:** 10.1101/2024.09.01.24310190

**Authors:** Jun Qiao, Minjing Chang, Miaoran Chen, Yuhui Zhao, Jiawei Hao, Pengwei Zhang, Ruixin Zhou, Liuyang Cai, Feng Liu, Xiaoping Fan, Siim Pauklin, Rongjun Zou, Zhixiu Li, Yuliang Feng

**Affiliations:** Department of Pharmacology, Joint Laboratory of Guangdong-Hong Kong Universities for Vascular Homeostasis and Diseases, School of Medicine, Southern University of Science and Technology, Shenzhen, China; School of Public Health and Emergency Management, School of Medicine, Southern University of Science and Technology, Shenzhen, China; Department of Nephrology, Shanxi Kidney Disease Institute, Second Hospital of Shanxi Medical University, Taiyuan, China; Department of Rheumatology, Shanxi Key Laboratory of Immunomicroecology, Second Hospital of Shanxi Medical University, Taiyuan, Shanxi, China; Botnar Research Centre, Nuffield Department of Orthopaedics, Rheumatology and Musculoskeletal Sciences, University of Oxford, Headington, Oxford, UK; Department of Cardiovascular Surgery, Guangdong Provincial Hospital of Chinese Medicine, the Second Affiliated Hospital of Guangzhou University of Chinese Medicine, the Second Clinical College of Guangzhou University of Chinese Medicine, Guangzhou, China; State Key Laboratory of Dampness Syndrome of Chinese Medicine, Guangzhou, China; Guangdong Provincial Key Laboratory of TCM Emergency Research, Guangzhou, China

## Abstract

**Objectives:** Autoimmune diseases (ADs) play a significant and intricate role in the onset of cardiovascular diseases (CVDs). Our study aimed to elucidate the shared genetic etiology between Ads and CVDs.

**Methods:** We conducted genome-wide pleiotropy analyses to investigate the genetic foundation comprehensively and shared etiology of six ADs and six CVDs. We analyze the genetic architecture and genetic overlap between these traits. Then, SNP-level functional annotation identified significant genomic risk loci and potential causal variants. Gene-level analyses explored shared pleiotropic genes, followed by pathway enrichment analyses to elucidate underlying biological mechanisms. Finally, we assess potential causal pathways between ADs and CVDs.

**Results:** Despite negligible overall genetic connections, our results revealed a significant genetic overlap between ADs and CVDs, indicating a complex shared genetic architecture spread throughout the genome. The shared loci implicated several genes, including *ATXN2*, *BRAP*, *SH2B3*, *ALDH2* (all located at 12q24.11-12), *RNF123*, *MST1R*, *RBM6*, and *UBA7* (all located at 3p21.31), all of which are protein-coding genes. Top biological pathways enriched with these shared genes were related to the immune system and intracellular signal transduction.

**Conclusions:** The extensive genetic overlap with mixed effect directions between ADs and CVDs indicates a complex genetic relationship between these diseases. It suggests overlapping genetic risk may contribute to shared pathophysiological and clinical characteristics and may guide clinical treatment and management.

## Introduction

Cardiovascular diseases (CVDs) constitute the leading cause of death worldwide^1^, with various factors contributing to their development. Autoimmune diseases (ADs), as a common cause, play a prominent and complex part in the onset of CVDs. When the recognition of the immune system goes wrong and inflammation goes out of control, it will lead to overactivity in immune activation and damage normal organs and tissues; the cardiovascular system is often injured more severely than other systems^2^. The influence of chronic inflammation and immune cell activation as ADs’ frequent biological pathways significantly contributes to the pathogenesis and progression of CVDs, such as pro-and anti-inflammatory cytokines in atherosclerotic plaque stability^3,4^ and myocardial dysfunctions^5^. Moreover, a population-based cohort study proved that the incidence of CVDs among patients with autoimmune disease was 10-15 times higher than those without an autoimmune disease, and the combination of more ADs means a higher risk of CVD^6^. As complex polygenic diseases, ADs and CVDs display strong phenotypic heterogeneity that many convergent processes, including genetic variation factors, might cause. Still, there has been no sufficient understanding of these diseases so far.

The high frequency of comorbidity observed in complex diseases is primarily driven by shared genetic architecture. Recent genome-wide association studies (GWASs) have identified numerous genetic risk loci for various ADs and CVDs, demonstrating significant interconnectedness due to shared loci effects. For example, IRF8, STAT4, IL19, and SRP54-AS1^7–9^ have been identified as potential shared genetic risk loci between systemic lupus erythematosus (SLE) and CVDs. However, most "causal" genetic variants or loci remain undiscovered at genome-wide significance across ADs and CVDs. Furthermore, genetic studies exploring genome-wide genetic correlations between specific ADs and CVDs have shown that results vary based on the methodology used. While polygenic risk scores (PRS) have identified significant correlations among some conditions, linkage disequilibrium score regression (LDSC) has not produced significant results. A significant genetic correlation estimated with LDSC requires consistent effect directions among shared variants across phenotypes. However, genetic correlation is inadequate in capturing polygenic overlap in cases where shared variants exhibit a combination of different effect directions. Capturing this "missing dimension" of genetic overlap, regardless of effect directions, is crucial to comprehensively understanding the shared genetic underpinnings between ADs and CVDs. Even with minimal genetic correlation, genetic overlap may suggest shared molecular mechanisms, offering a thorough understanding of the shared genetic landscape between ADs and CVDs.

The shared genetic basis may be explained by genetic variants that impact multiple complex phenotypic traits through vertical and/or horizontal pleiotropy^10^. Specifically, Mendelian Randomization (MR), an approach predominantly based on vertical pleiotropy, explores causal relationships between ADs and CVDs. However, only a few trait pairs have been consistently linked causally in multiple studies, often with conflicting results. For example, investigations into the causal connections between SLE and coronary artery disease (CAD), as well as between rheumatoid arthritis (RA) and atrial fibrillation (AF), have produced contradictory outcomes^11–14^. This inconsistency highlights the limitations of MR in deciphering the genetic mechanisms of these diseases, particularly its underutilization of genome-wide markers and vulnerability to the influence of heritable confounders affecting the link between exposure and outcome. With the advancement of statistical tools and a broadening understanding of genetic mechanisms, more and more studies are exploring diseases’ shared genetic underpinnings through horizontal pleiotropy. For example, recent research on the genetic overlap between gastrointestinal tract diseases and psychiatric disorders has demonstrated that pleiotropic genetic determinants, widely distributed across the genome, may define a shared genetic architecture at the levels of SNPs, loci, genes, and pathways^15^. Furthermore, horizontal pleiotropy has revealed evidence of shared genetic effects between ADs and allergic diseases, suggesting novel strategies for elucidating the genetic bases, biology, and therapeutic targets of complex immune-related traits^16^.

This genome-wide pleiotropic association study leveraging large-scale data and various techniques that capture shared genetic background was used to increase coverage of human interactome mapping of ADs and CVDs and novel perspectives for SNP-to-gene and pathway. We investigated shared genetic mechanisms representing general risk across 6 major ADs (RA, SLE, type 1 diabetes [T1D], ulcerative colitis [UC], Crohn’s disease [CD], primary sclerosing cholangitis [PSC]) and 6 major CVDs (AF, CAD, venous thromboembolism [VTE], heart failure [HF], peripheral artery disease [PAD], stroke) to investigate the shared genetic foundation (including genetic correlation and overlap) and genetic mechanism sequentially. In the vertical pleiotropy study, we focus on the pairwise traits causal associations utilizing the Latent Heritable Confounder MR (LHC-MR) method, which estimates bi-directional causal effects, direct heritabilities, and confounder effects while considering sample overlap. Initially, pleiotropic variants and loci were detected through SNP-level analysis in horizontal analysis, then colocalized loci with strong evidence were identified using pairwise colocalization analysis. Further analyses at the gene level were conducted to pinpoint pleiotropic genes, which included parallel position-specific mapping and tissue-specific enrichment analysis. Biological pathways across trait pairs will promote and reshape the understanding of ADs and CVDs, offering valuable knowledge for preventing and treating comorbidity. Finally, we consulted the Drug Gene Interaction Database (DGIdb) to verify whether these and neighboring genes were recognized as drug targets, thus facilitating novel interventions.

## Results

### Genetic correlation between ADs and CVDs

We used cross-trait linkage disequilibrium (LD) score regression (LDSC) for the calculation SNP-based heritability (*h^2^_SNP_*) and the estimation of genome-wide genetic correlation (*r_g_*) between six major ADs and six major CVDs. Univariate LDSC analysis revealed that the estimated *h^2^_SNP_*for ADs were substantially higher than those for CVDs, approximately 16 greater on average. Specifically, SLE exhibited the highest heritability (*h^2^_SNP_*= 0.576, SE = 0.081), while T1D displayed the lowest (*h^2^_SNP_* = 0.033, SE = 0.004). Among six major CVDs analyzed, CAD exhibited relatively high heritability (*h^2^_SNP_*=0.034, SE=0.002), approximately six times greater than that of PAD, which had the lowest heritability (*h^2^_SNP_* = 0.006, SE=0.001) (Fig. 1a and Supplementary Table 2a). Bivariate LDSC analysis revealed positive significant genetic correlations in 9 out of 36 trait pairs between ADs and CVDs (*P* < 0.05), with coefficients ranging from 0.087 to 0.207. Of these, only the trait pair of UC and VTE the stringent Bonferroni correction threshold (*r_g_* = 0.117, SE=0.036, *P* = 1.10×10^-3^) (Fig. 1b and Supplementary Table 2b).

**Fig. 1:**
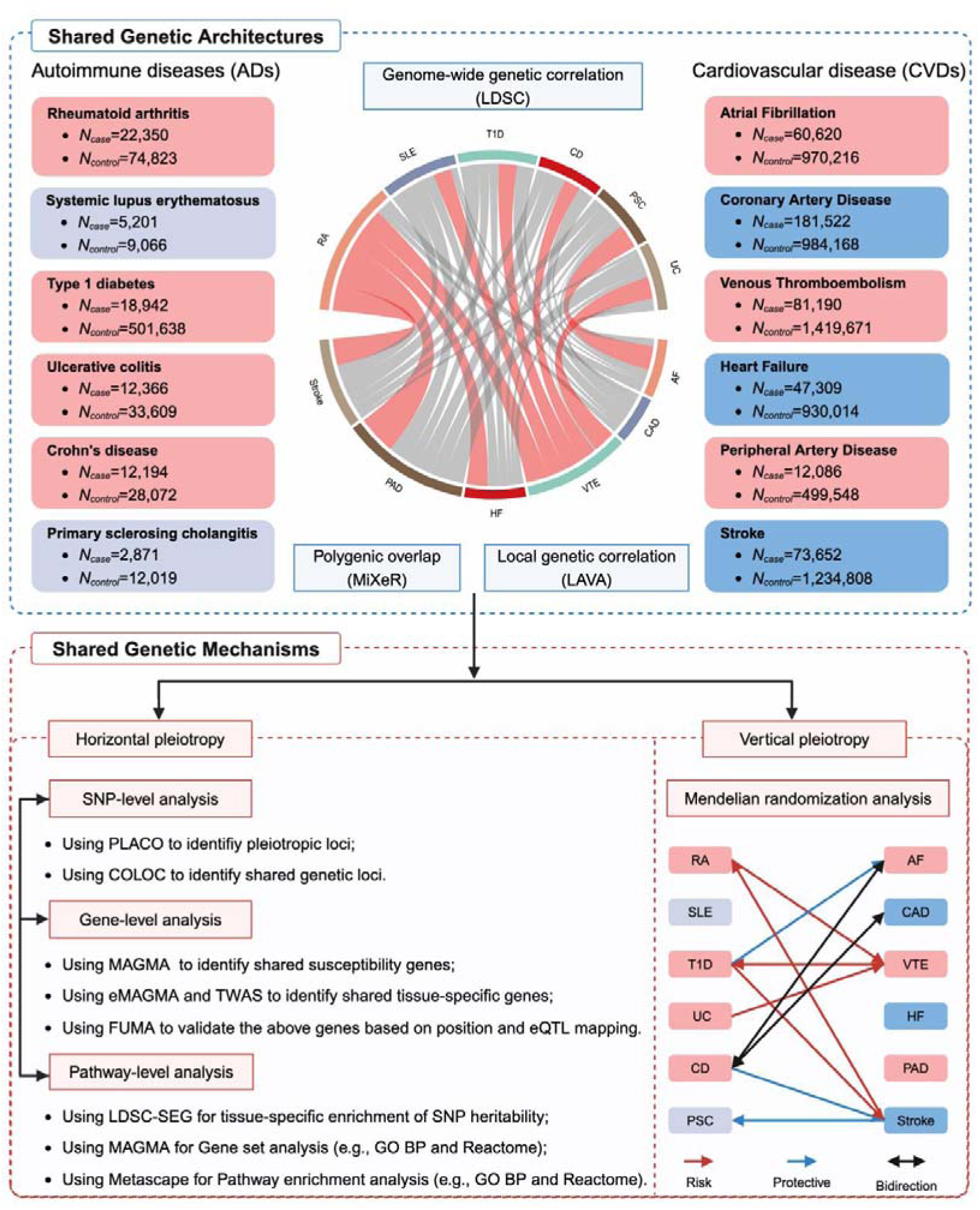
Workflow for the study of six autoimmune diseases and six cardiovascular diseases. From different perspectives, a comprehensive pleiotropic analysis was conducted on six autoimmune diseases (ADs) and six cardiovascular diseases (CVDs). Utilizing large GWAS datasets from individuals of European ancestry, we initially characterize the genetic architecture and overlap between these diseases at genome-wide, polygenic, and local levels. We then apply novel statistical tools to discern distinct forms of genetic pleiotropy, specifically vertical and horizontal pleiotropy. Our analysis commences with SNP-level functional annotation, identifying significant genomic risk loci and potential causal variants. This is complemented by gene-level analyses investigating shared pleiotropic genes, thereby deepening our understanding of the genetic bases of these conditions. Pathway enrichment analyses further illuminate the underlying biological mechanisms, paving the way for identifying therapeutic targets. Finally, we assess potential causal pathways between ADs and CVDs, focusing on capturing evidence of vertical pleiotropy. This comprehensive pleiotropic analysis enabled us to identify shared genetic backgrounds, enhancing coverage of human interactome mapping for ADs and CVDs while also providing novel insights into SNP-to-gene relationships and pathway associations. The diagram was generated using BioRender (www.biorender.com) and has been included with permission for publication. Rheumatoid arthritis, RA; Systemic lupus erythematosus, SLE; Type 1 diabetes, T1D; Crohn’s disease, CD; Ulcerative colitis, UC; Primary sclerosing cholangitis, PSC; AF, Atrial fibrillation; CAD, Coronary artery disease; VTE, Venous thromboembolism; HF, Heart failure; PAD, Peripheral artery disease.

While the genetic correlation coefficient *r_g_* quantifies the genetic correlation between two traits, it may not differentiate between genetic overlap caused by a mixture of concordant and discordant effects and the total lack of genetic overlap, which could lead to an *r_g_* value close to zero in both situations. Consequently, more than LDSC analysis is needed to capture the complex dimensions of genetic overlap fully. To address this, we have employed recently established statistical methods, including the causal mixture modeling approach (MiXeR) and Local Analysis of [co]Variant Annotation (LAVA), to comprehensively characterize the genetic overlap between ADs and CVDs beyond mere genetic correlation.

### Genetic overlap between ADs and CVDs

MiXeR identified genetic overlap regardless of the direction of effect, which complements genetic correlation to provide a more thorough understanding of the genetic relationships among phenotypes. MiXeR considers differences in polygenicity to determine which phenotypes may have shared genetic variants. Univariate MiXeR analyses revealed that CAD (N = 1.795K, SD = 0.101K) and HF (N = 2.231K, SD = 0.317K) exhibited higher polygenicity, while SLE and PSC displayed lower polygenicity, suggesting a polygenicity pattern distinct from *h^2^_SNP_* estimates in ADs and CVDs. Bivariate MiXeR analyses showed weak to moderate but distinct patterns of genetic overlap between ADs and CVDs, with the Dice coefficients ranging from 0.021 to 0.307 (Supplementary Table 3a). For example, consistent with the strongest positive genome-wide genetic correlation (*r_g_* = 0.211, SE = 0.027) and a positive genetic correlation of shared variants (*r_g_s* = 0.890, SE = 0.087), a pronounced genetic overlap was observed between RA and PAD. This was reflected by a Dice coefficient of 0.238 (SD = 0.038), with 0.117K shared variants (SD = 0.019K) accounting for 22.1% of the variants affecting RA and 25.7% of the variants affecting PAD, respectively. Despite the lack of significant *r_g_* in the LDSC analyses, RA and CAD demonstrated extensive genetic overlap, evidenced by a Dice coefficient of 0.208 (SD = 0.033). This suggests a mixed direction of effect between RA and CAD, further validated by a significant proportion (59.5%) of shared variants exhibiting consistent effects. When shared variants have both concordant and discordant effect directions, they nullify each other, masking genetic correlation at the genome-wide level (Fig. 2c). Given the low polygenicity observed in ADs such as SLE and PSC and the high polygenicity in CVDs like CAD and HF, substantial disparities were noted in the number of shared and unique "causal" variants. For example, SLE and CAD shared a relatively low number of variants (N = 0.019K, SD = 0.009K), while there were significantly more unique variants for CAD (N = 1.775K, SD = 0.104K) compared to SLE (N = 0.023K, SD = 0.010K). These unique variants accounted for 45.7% of the variants influencing SLE and only 1.07% of those influencing CAD. Consequently, SLE and CAD exhibited minimal genetic overlap (Dice coefficient = 0.021, SD = 0.011) and demonstrated a *r_g_* approaching zero (*r_g_* = 0.036, SE = 0.031) (Fig. 2b, Supplementary Fig. 1, Supplementary Table 3b). Finally, MiXeR results indicate that the model fits for SLE-AF, UC-PAD, and PSC-Stroke are suboptimal, as evidenced by negative Akaike Information Criterion (AIC) scores. When comparing the best-fit model to the minimum possible overlap (minima), these scores suggest that the shared genetic component between these trait pairs may be less than previously estimated.

**Fig. 2:**
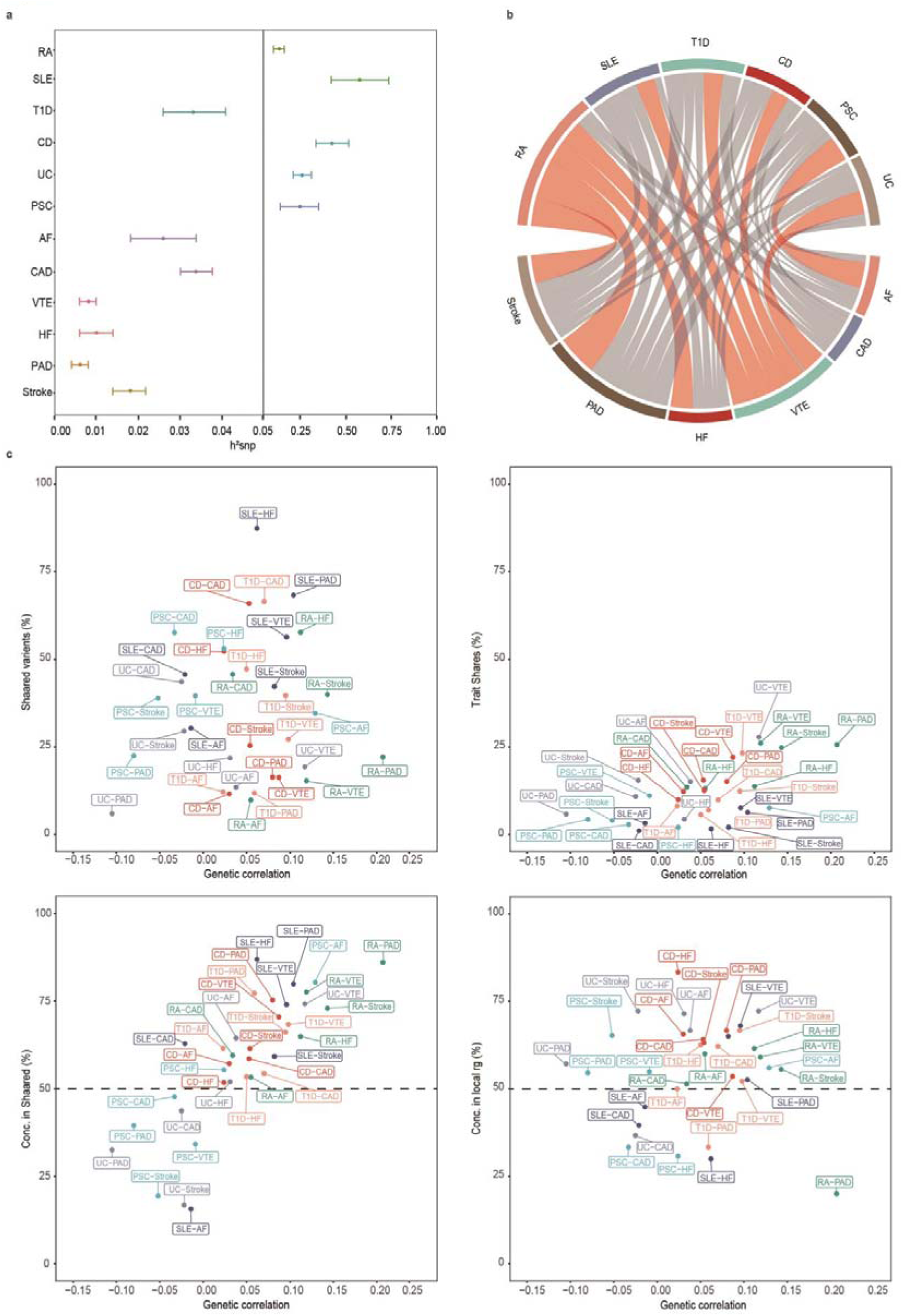
Genetic overlap between six autoimmune diseases and six cardiovascular diseases beyond genome-wide genetic correlation. **(a)** Error-bar plot of the SNP-based heritability (*h_SNP_^2^*) point estimates for six ADs and six CVDs, computed by univariate LDSC. **(b)** Network visualization of the Bonferroni-corrected significant global genetic correlations (*r_g_*) between six ADs and six CVDs, computed by bivariate LDSC. Connections represent significant *r_g_* values, with correlation values along the connections; thicker lines denote stronger correlations. Blue indicates negative correlations, red indicates positive correlations, and dark gray indicates insignificant correlations. The size of the nodes is weighted by the sample size and *h_SNP_^2^* of the given phenotype (size = *h_SNP_^2^* × sqrt (N)). **(c)** Genetic correlation estimated by LDSC (x-axis) against the percentage of AD variants shared with CVDs (first plot), the percentage of CVD variants shared with ADs (second plot), and the percentage of CVD variants shared with ADs that have concordant effect directions (third plot). The fourth plot shows the percentage of local genetic correlations from LAVA with concordant effect directions on the y-axis. RA: Rheumatoid arthritis; SLE: Systemic lupus erythematosus; T1D: Type 1 diabetes; CD: Crohn’s disease; UC: Ulcerative colitis; PSC: Primary sclerosing cholangitis; AF: Atrial fibrillation; CAD: Coronary artery disease; VTE: Venous thromboembolism; HF: Heart failure; PAD: Peripheral artery disease.

### Local genetic correlation between ADs and CVDs

Genetic variance in small genomic regions may be shared by pairs of ADs and CVDs, even without a significant genome-wide genetic correlation. We conducted LAVA analyses to estimate broad local genetic correlations between ADs and CVDs in 2,495 unique genomic regions, further elucidating the direction of the mixed effects observed. Local genetic correlations (*r_g_s*) showed that only 58.7% of nominally significant local *r_g_s* were in the positive direction between the AD-CVD phenotype pair (Supplementary Table 4-5). A mixture of negative and positive local *r_g_s* were observed for each pair, potentially leading to minimal genetic correlations at the genome-wide level. Supporting the MiXeR findings, further evidence of mixed effect directions was evident among RA-CAD (18 positively and 17 negatively correlated loci) and SLE-CAD (19 positively and 29 negatively correlated loci). Interestingly, many loci had negative local genetic correlations between RA and PAD, somewhat divergent from positive genetic correlations (8/10, 80%). After correcting for multiple tests using Bonferroni correction, we also identified 23 loci that exhibited a significant local genetic correlation without a significant global correlation (Fig. 2c, Fig. 5, Supplementary Table 5). Our investigation also identified three regions (LD block 1,841, chr12: 111,592,382-113,947,983; LD block 100, chr1: 113,418,038-114,664,387; LD block 2,048, chr15: 37,962,916-39,238,840) displaying significant correlations for more than one trait pair. Overall, these findings indicate that global genetic correlations cannot fully represent the heterogeneity in genetic associations between phenotypes.

### Shared genetic loci and functional annotation for ADs and CVDs

Despite these advances, the shared genetic mechanisms between ADs and CVDs remain unclear. Uncertainty persists regarding whether the genetic basis observed predominantly reflects horizontal pleiotropy, whereby the same genetic variant affects both traits. At the most fine-grained level of analysis, PLACO analyses are concerned with estimating SNP-level effects on phenotypes and identified 233,00 SNPs with potentially pleiotropic effects across 36 trait pairs between ADs and CVDs. FUMA annotation subsequently clustered these pleiotropic SNPs into 815 lead SNPs and 679 independent genomic risk loci across 208 unique chromosomal regions. Notably, 131 pleiotropic loci were identified across multiple trait pairs, with six of these loci exhibiting genetic signals in more than one-third of the trait pairs, suggesting a potentially broad functional impact of specific genomic regions (Supplementary Fig 2, Supplementary Fig 3, Supplementary Table 6-9). For example, the loci spanning 12q24.1-q24.12 on chromosome 12 overlaps with 30 trait pairs, which does not involve SLE-CAD and any related to AF except for T1D-AF. Notably, rs4766578 at 12q24.12 showed a remarkably consistent degree of pleiotropy across most trait pairs and was located in the binding sequence of the transcription factor HNF4A, a crucial regulatory element of *ALDH2*. This transcription factor has previously been linked to significant health outcomes, including blood pressure, cardiovascular disease, and autoimmune disease. Interestingly, the locus 17q12 on chromosome 17 was jointly associated with all ADs and AF, except for PSC-AF. This locus surrounds SNP rs1008723, located in the intronic region of the gasdermin B gene [*GSDMB*]. *GSDMB* encodes a family of structurally related proteins that play crucial roles, particularly in pyroptosis, a process implicated in the pathogenesis of ADs such as IBD and CVDs due to its involvement in severe cytokine release and inflammation. Overall, 345 SNPs (50.8%) exhibited novel associations with ADs, while 354 SNPs (52.1%) displayed novel associations with CVDs. Notably, 79 of these SNPs are reported for the first time about ADs and CVDs, suggesting potential implications for the immune and cardiovascular systems that warrant further investigation. More than half (51.3%) of the significant SNPs identified by PLACO have opposing genetic effects on the two diseases, suggesting different underlying causes for ADs and CVDs and potentially explaining the weak genetic correlation in the above analyses.

ANNOVAR annotation revealed that out of 679 top lead SNPs, 176 (25.9%) were intergenic variants, 352 (51.8%) were intronic variants, and 39 (5.7%) were exonic variants. Among these exonic variants, the SNP rs10781542, located at the 9q34.3 locus on chromosome 9 (*P_PLACO_*= 1.04×10^-9^ for CD-Stroke), had the highest RDB score of 1a, indicating strong evidence of functionality. Additionally, 49 SNPs (7.2%) had CADD scores above 12.37, with rs601338 at the 19q13.33 locus on chromosome 19 (*P_PLACO_* = 3.77×10^-10^ for T1D-CAD) presenting the highest CADD score of 52, suggesting potential deleterious effects. Further colocalization analysis revealed that 112 (16.5%) out of 679 pleiotropic loci exhibited PP.H4 greater than 0.7, identifying 11 unique SNPs as candidate-shared causal variants. Additionally, 93 (13.7%) pleiotropic loci showed PP.H3 greater than 0.7, suggesting the presence of different causal variants within these loci (Supplementary Fig 4, Supplementary Table 6).

### Candidate pleiotropic genes between ADs and CVDs

Instead of focusing on single SNPs, we conducted a gene-centered pleiotropy analysis by collectively analyzing sets of SNPs located within genes. MAGMA analysis identified 662 pleiotropic genes, of which 191 are unique, located within or overlapping with 679 pleiotropic loci. Notably, 590 genes (89.1%, 119 unique) were detected in at least two trait pairs (Fig. 3, Supplementary Table 10-12). Furthermore, four unique pleiotropic genes were detected in over one-third of the trait pairs, including *ATXN2*, *BRAP*, *ALDH2*, and *SH2B3*, all located at the 12q24.1-q24.12 loci. Ataxin 2 [*ATXN2*] is a polyglutamine protein primarily involved in various biological processes, including RNA translation and cytoskeletal reorganization.

**Fig. 3:**
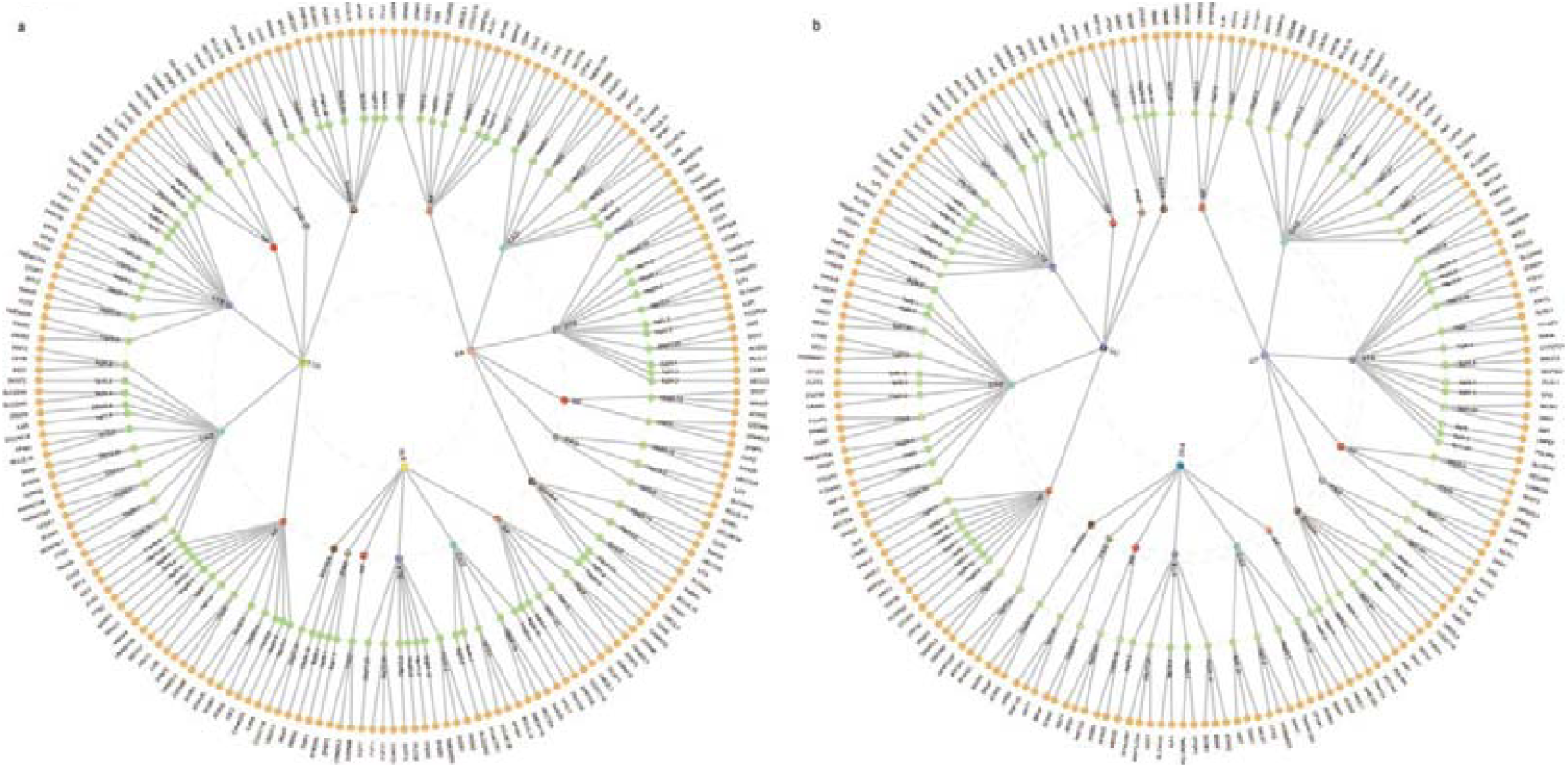
The Overall landscape of the pleiotropic associations across six autoimmune diseases and six cardiovascular diseases. **(a)** A circular dendrogram displays the shared genes among three ADs (first circle: RA, TID, and SLE) and six CVDs (second circle), resulting in 18 pairs. **(b)** Another circular dendrogram displays the genes shared between three ADs (first circle: CD, UC, and PSC) and six CVDs (second circle), also resulting in 18 pairs. In total, 679 shared loci were identified across 36 trait pairs, mapped to 662 significant pleiotropic genes (191 unique) identified through multimarker analysis using GenoMic annotation (MAGMA). For trait pairs with more than three pleiotropic genes, only the top 3 were displayed based on candidate pleiotropic gene prioritization (fourth circle). Rheumatoid arthritis, RA; Systemic lupus erythematosus, SLE; Type 1 diabetes, T1D; Crohn’s disease, CD; Ulcerative colitis, UC; Primary sclerosing cholangitis, PSC; AF, Atrial fibrillation; CAD, Coronary artery disease; VTE, Venous thromboembolism; HF, Heart failure; PAD, Peripheral artery disease.

Recent studies have suggested that ataxin-2 deficiency is associated with dyslipidemia, potentially impacting the normal metabolism of the cardiovascular system. Rare variants in *ATXN2* have been proposed to be related to obesity, insulin-resistance, and diabetes mellitus. Obesity may trigger and maintain a chronic low-level inflammatory state that can worsen autoimmune conditions and their related complications. Inflammatory stimuli increase the expression of BRCA1-associated protein [*BRAP1*], which in turn promotes the release of inflammatory cytokines, thereby elevating the likelihood of atherosclerosis, a key contributor to cardiovascular disease development. Accumulated evidence demonstrates that *BRCA* is rapidly recruited to DNA lesions and plays a crucial role in the DNA damage response, potentially mediating autoimmune and systemic immune-mediated diseases. In addition, these results suggest that 225 pleiotropic genes (34.0%) are novel candidate genes for ADs, while 312 genes (47.1%) are associated with CVDs. Notably, *ATXN2*, *BRAP*, and *SH2B3* were not previously reported to be associated with both traits. A total of 644 genes (97.3%) identified by MAGMA were confirmed using FUMA positional mapping (Supplementary Table 8).

### Tissue-specific pleiotropic genes between ADs and CVDs

We applied stratified LDSC to specifically expressed genes (LDSC-SEG) to connect genetic discoveries to pertinent tissues and cell types, offering an understanding of the role of particular tissue or cellular functions in the genetic basis of a trait. Some of our findings from analyzing gene expression data align with established biological knowledge: immunological traits exhibit immune cell-type enrichments, while cardiovascular traits are strongly enriched in tissues such as the heart’s left ventricle and arterial tissues, including the aorta, coronary artery, and tibial artery. Chromatin data from the Roadmap Epigenomics and ENCODE projects confirmed the multiple-tissue gene expression analysis described above (Fig.4, Supplementary Table 13).

**Fig. 4:**
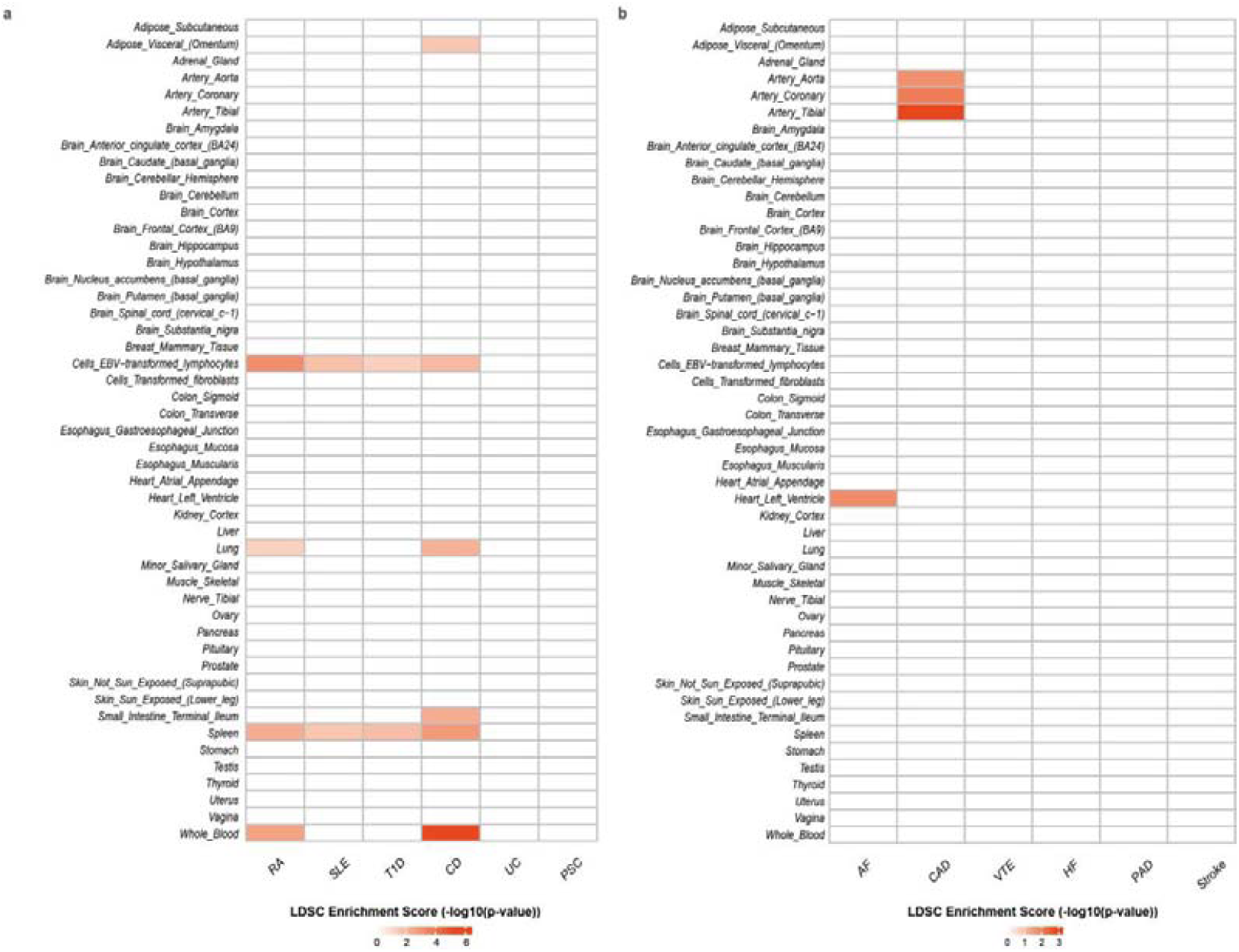
The results of multiple-tissue analysis using gene expression data for six autoimmune diseases and six cardiovascular diseases. Heatmap of tissue type-specific enrichment of single nucleotide polymorphism (SNP) heritability for ADs and CVDs in 53 tissues from GTEx v8, estimated using stratified LDSC applied to specifically expressed genes (LDSC-SEG). The x-axis reflects disease types: ADs **(a)** and CVDs **(b)**, and the y-axis reflects a tissue from the GTEx dataset. Red represents significant enrichment with a *P*-value after FDR correction (FDR < 0.05). The color gradient indicates the magnitude of values, with different colors corresponding to different ranges of values. Dark red represents higher values. RA, Rheumatoid arthritis; SLE, Systemic lupus erythematosus; T1D, Type 1 diabetes; CD, Crohn’s disease; UC, Ulcerative colitis; PSC, Primary sclerosing cholangitis; AF, Atrial fibrillation; CAD, Coronary artery disease; VTE, Venous thromboembolism; HF, Heart failure; PAD, Peripheral artery disease.

**Fig. 5:**
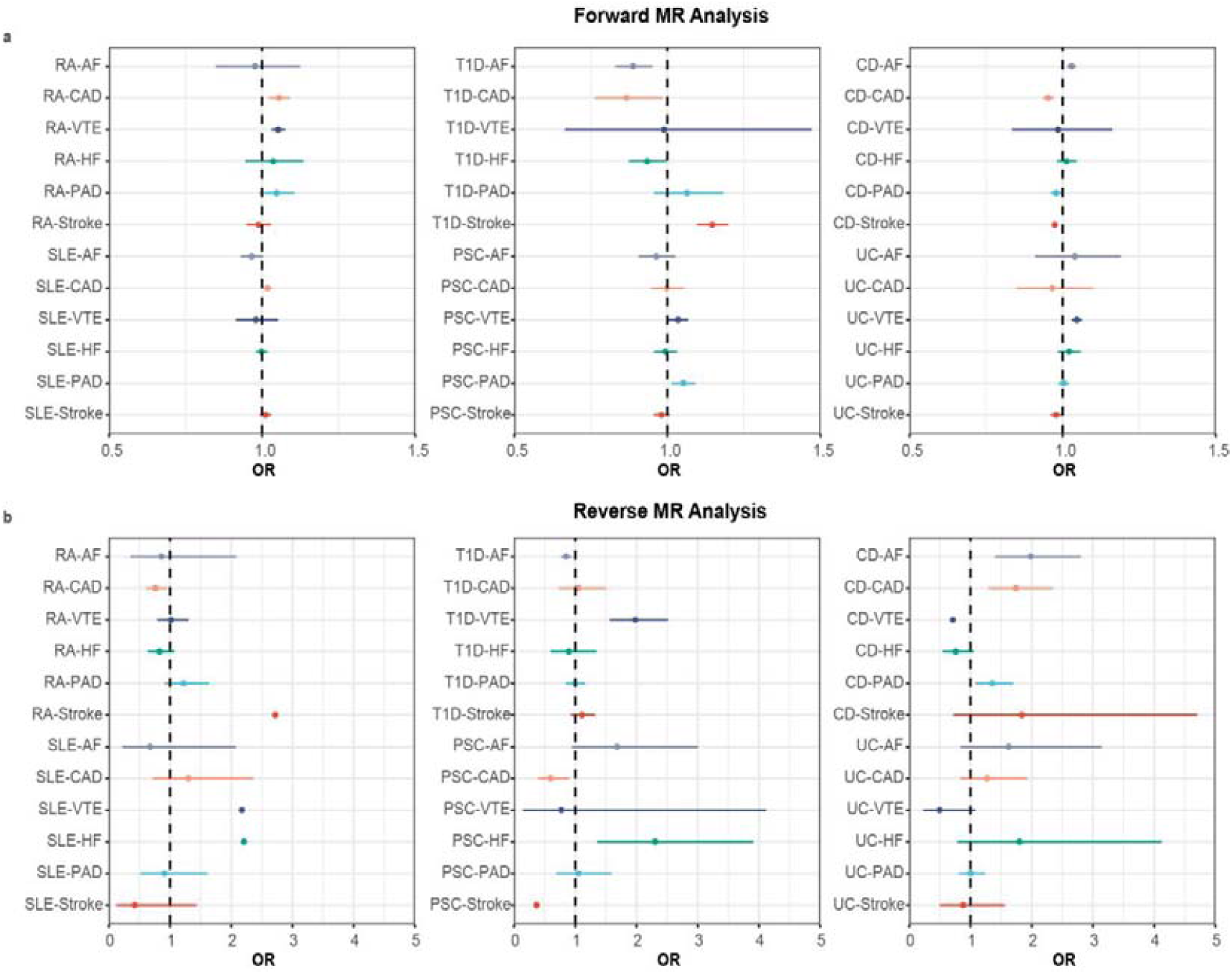
The causal inference between six autoimmune diseases and six cardiovascular diseases. Summary of putative causal relationships between ADs and CVDs identified by LHC-MR. Forest plot of the LHC-MR analysis on the associations between ADs and CVDs. Circles represent the odds ratio (OR) estimate, and the error bars indicate the 95% confidence interval. The top part of the results **(a)** represents the estimated causal effect of ADs on CVDs, while the bottom part **(b)** represents the estimated causal effect of CVDs on ADs. A positive association is indicated by OR > 1, while a negative association is indicated by OR < 1. Note that the first plot was plotted at OR truncated by 1.5 for better visualization, thus excluding SLE-PAD. Abbreviations: RA, Rheumatoid arthritis; SLE, Systemic lupus erythematosus; T1D, Type 1 diabetes; CD, Crohn’s disease; UC, Ulcerative colitis; PSC, Primary sclerosing cholangitis; AF, Atrial fibrillation; CAD, Coronary artery disease; VTE, Venous thromboembolism; HF, Heart failure; PAD, Peripheral artery disease.

While it is commonly assumed that the nearest gene is often the causal gene, this isn’t always true. MAGMA mainly focuses on variants near the gene boundary, potentially overlooking significant variant-gene associations. E-MAGMA, based on MAGMA, integrates genetic and transcriptomic data (e.g., eQTLs) to identify risk genes, thus enhancing the utilization of distal variant-gene associations. Additionally, e-MAGMA could assist in pinpointing actual susceptibility genes in the tissue context by leveraging eQTLs of potentially phenotype-associated tissues. A total of 5,483 pleiotropic tissue-specific genes (550 unique) were identified in at least one tissue (Supplementary Table 14). Ten genes, including *RBM6*, *UBA7*, *MST1R*, *RNF123* (located at 3p21.31), *GSDMB*, *ORMDL3*, *PGAP3* (all located at 17q12-q21.1), *ALDH2*, *TMEM116* and *SH2B3* (all located at 12q24.11-12) were identified in the greater than or equal to one-third of trait pairs. Four genes at 3p21.31 were identified as candidate risk genes for UC, CD, and PSC for three disease-specific conditions. For example, evidence supported the role of RNA binding motif protein 6 [*RBM6*] in IBD by participating in the intestinal immune network for IgA production. Dysregulation of RNA-binding proteins like *RBM6* can lead to aberrant immune responses, significantly contributing to hypertension and thereby increasing the risk of CVDs. Ubiquitin-like modifier activating enzyme 7 [*UBA7*] has been identified as a target gene for IBD-associated variants, which influence immune responses. Given the association between IBD and PSC, *UBA7* may also play a role in PSC pathogenesis. It is also known to activate ISG15, a ubiquitin-like protein, which contributes to heart failure development by regulating cardiac amino acid metabolism and altering cardiomyocyte protein turnover. Transcriptome-wide association Scanning (TWAS) validated the e-MAGMA analyses using single-trait GWAS results (Supplementary Table 15). A total of 45.1% of tissue-specific pleiotropic genes were identified as novel for ADs and 84.9% for CVDs. A total of 30.4% of genes identified by e-MAGMA were confirmed through FUMA eQTL mapping (Supplementary Table 8).

In conclusion, 388 pleiotropic genes (130 unique) were finally identified through the combined use of MAGMA and e-MAGMA, in which *ALDH2* and *SH2B3* were detected in over or equal to one-third of the trait pairs (Supplementary Table 10). Except for *ALDH2* and *SH2B3* in SLE-CAD, located at 12p24.12, *ALDH2* and *SH2B3* for other trait pairs are located at the 12q24.11-12 locus. The aldehyde dehydrogenase two family member [*ALDH2*] significantly inhibited mitophagy during reperfusion, attenuated hypoxia/reoxygenation-induced cardiomyocyte contractile dysfunction, and may serve as a primary target for cardioprotection. Additionally, overexpression of *ALDH2* protects against oxidative stress-induced inflammatory events that lead to cellular or tissue injury and protects ADs. The SH2B adaptor protein 3 [*SH2B3*], an adaptor protein, negatively regulates cytokine signaling and cell proliferation. This function contributes to an increased risk of various autoimmune diseases, potentially due to *SH2B3*’s impact on impairing the adverse selection of immature or transitional self-reactive B cells. Furthermore, *SH2B3* has been implicated in causing heart injury by promoting a proinflammatory response and impairing insulin signaling. Remarkably, the disease-specific *RNF123* and *MST1R* genes at locus 3p21.31 identified in the e-MAGMA analysis were still present. RING finger protein [*RNF123*] plays a significant role in the immune response, mainly through the TLR3/IRF7-mediated pathway that promotes type 1 interferon (IFN) expression, thereby exacerbating chronic inflammation in IBD. Research indicates that *RNF123* can influence the stability of critical proteins involved in the inflammatory response, such as those in the NF-κB signaling pathway, which is also implicated in atherosclerosis and other cardiovascular conditions. Macrophage-stimulating one receptor [*MST1R*], also known as RON receptor tyrosine kinase, is critical in regulating inflammatory responses and tissue repair. Research has shown that *MST1R* is involved in several key signaling pathways that mediate immune responses and fibrotic processes, which are central to the pathogenesis of PSC. Moreover, *MST1R* signaling pathways could intersect with those involved in lipid metabolism and oxidative stress, which are critical in the pathogenesis of CVDs.

### Shared biological mechanisms between ADs and CVDs

Pathway and gene set approaches, by aggregating and analyzing signals at the gene level within functional pathways, reveal the functional and biological characteristics of genes that confer risk for a particular phenotype. Here, we note that the associated genes collectively perturb various nodes in T cell activation and signaling pathways, yet different disease clusters show distinct patterns of genetic associations (Supplementary Table 16a, Supplementary Table 16b). Notably, the gene *SH2B3*, associated with most trait pairs, was significantly enriched in multiple gene sets within the lower layers of ‘intracellular signal transduction,’ including ‘regulation of the MAPK cascade’ and ‘regulation of phosphatidylinositol 3-kinase/protein kinase B signal transduction’. In autoimmune diseases such as RA and SLE, dysregulation of the MAPK cascade can lead to aberrant T-cell activation and inflammatory cytokine production, perpetuating the autoimmune response. Excessive activation of the MAPK cascade can also contribute to the development of atherosclerosis by promoting endothelial cell dysfunction and inflammatory processes within the vascular wall. The PI3K/AKT pathway is integral to various cellular functions, including growth, survival, metabolism, and immune responses. In the lower layers of the ‘innate immune system,’ CD, UC, and PSC were significantly associated with ‘antigen processing and presentation of peptide antigen via MHC class I,’ whose dysregulation can lead to immune responses against self-antigens, contributing to autoimmune pathology. For instance, studies have implicated aberrant MHC class I antigen presentation in CD, where T cells recognize self-peptides as foreign, triggering chronic inflammation in the gastrointestinal tract. Similarly, in UC, defective antigen processing mechanisms may result in the immune system attacking the intestinal lining, exacerbating symptoms.

### Drug-potential target network

Using STRING V.11.5, we identified ten biologically related genes of *SH2B3*, *ATXN2*, *BRAP*, and *ALDH2*, respectively. Subsequent queries in the Drug Gene Interaction Database (DGIdb) revealed that *SH2B3*, *ATXN2*, *ALDH2*, and their associated genes, such as *JAK2* (linked to *SH2B3*), are targeted in various treatments for ADs and CVDs. This discovery is consistent with the known role of *JAK2* in mediating immune responses and regulating T-cell differentiation. We also identified 444 FDA-approved drugs and 858 potential candidates targeting these 28 unique genes. Notably, paclitaxel, an anti-inflammatory drug targeting *JAK2*, demonstrates potential as a therapeutic option for ADs and is already approved for various CVDs, including PAD. Furthermore, the potential for repurposing antitumor drugs like fedratinib to prevent or treat ADs and CVDs merits further exploration in clinical trials, as detailed in Supplementary Table 17.

### Causal relationships between ADs and CVDs

Mendelian randomization analysis could detect causal trait pairs and partially reflect vertical pleiotropy. In the Latent Heritable Confounder MR (LHC-MR) analysis, after correcting for multiple comparisons using Bonferroni correction, we found convincing evidence (P < 2.25×10-7) for causal effects in five trait pairs, including RA-VTE, T1D-AF, T1D-Stroke, CD-Stroke, and UC-VTE (Supplementary Table 18). For example, for a one-unit increase in log odds of UC (equalling a one-unit increase in the prevalence of UC), the odds ratios were 1.046 (95% CI, 1.029, 1.063) for VTE. In the reverse analysis, convincing evidence of genetic causality emerged for three pairs (VTE-T1D, Stroke-RA, and Stroke-PSC). For example, genetic liability to VTE showed a positive association with T1D (OR 1.981; 95% CI, 1.560-2.515). Furthermore, two trait pairs (CD-AF and CD-CAD) hinted at a bidirectional causality. Overall, MR analysis demonstrated strong evidence of genetic causality in 10 trait pairs analyzed in either direction, suggesting that vertical pleiotropy may mediate their relationship.

## Discussion

This genome-wide pleiotropic association study, based on European ancestry, identified shared genetic components across six major ADs and six major CVDs. Notably, most variants associated with CVDs also influenced ADs, reflecting their greater polygenicity and exhibiting weak to moderate patterns of genetic overlap. Additionally, we discovered significant local genetic correlations between ADs and CVDs within specific regions, including three regions with notable correlations across multiple traits. Our analysis also uncovered 131 pleiotropic loci, with six of these loci showing genetic signals in more than one-third of the trait pairs and consistently exhibiting the same effect direction for both traits. Our gene-level pleiotropy analysis utilized two mapping strategies: position and eQTL mapping. This approach identified extensive co-inheritance across 338 genes, representing 87.1% of the 388 critical pleiotropic genes detected across various trait pairs. Notably, pleiotropic genes located at 12q24.11-q24.12, such as *BRAP* (18/36), *ATXN2* (18/36), *SH2B3* (17/36), and *ALDH2* (15/36), as well as *GSDMB* (10/36) at 17q12 and *RNF123* (12/36) at 3p21.31, both influenced over 1/4 trait pairs, underscoring their profound impact on shared genetic associations. Further biological mechanisms analysis suggested that the MAPK cascade and the PI3K/AKT signaling pathway might be involved in common underlying genetic vulnerabilities. Moreover, the LHC-MR results indicated causal effects between Ads and CVDs in either direction for 10 trait pairs, supporting evidence of vertical pleiotropy. Moreover, *SH2B3*, *ATXN2*, *ALDH2* and its functionally related genes, such as JAK2 related to *SH2B3*, were identified as therapeutic targets for both ADs and CVDs. Collectively, these findings provide deeper insights into the shared genetic architectures of ADs and CVDs, implicating novel molecular mechanisms and potential therapeutic targets.

Given the complex genetic architecture of ADs and CVDs and their prevalent comorbidities, examining their shared genetic underpinnings is crucial. Genome-wide genetic correlations, quantified using LDSC, revealed weak to moderate associations between ADs and CVDs, with nine trait pairs surpassing the significance threshold. However, only the association between UC and VTE remained significant under a stringent Bonferroni correction. To discern between mixtures of concordant and discordant genetic effects versus an absence of genetic overlap, we employed MiXeR and LAVA. These analytical tools shed light on the polygenic overlap and local genetic correlations, thus deepening our understanding of the shared genetic foundations between ADs and CVDs. Notably, the genetic interactions between RA and CAD provide a compelling case study for examining the influence of confounding factors. MiXeR analysis indicated significant polygenic overlap between RA and CAD, with most RA risk variants also affecting CAD and exhibiting concordant effect directions. Additionally, LAVA analysis uncovered 18 positively and 17 negatively correlated regions between RA and CAD, corroborating the extensive genetic overlaps identified by MiXeR. These results indicate that approximately equal proportions of positive and negative correlation regions may obscure the true extent of genome-wide genetic correlations, potentially leading to an underestimation of the genetic linkages between ADs and CVDs. This phenomenon is also observed in weak or insignificant genome-wide genetic correlations, such as those between SLE and CAD, where minimal genetic overlap and a correlation nearing zero were observed. This minimal correlation likely stems from most variants being unique to SLE, exerting negligible effects on CAD. Further, LAVA analysis identified 19 positive and 29 negative correlated regions, which may have masked the genome-wide genetic correlation. These comprehensive analyses highlight the complexity of trait correlations and suggest that comorbidities may arise more from the distribution and direction of highly pleiotropic variants than phenotype-specific ones, enriching our understanding of the underlying polygenic overlap and the intricate genetic interplay between ADs and CVDs.

Within a sophisticated genetic framework, several mechanisms may influence the effects of genetic variations on ADs and CVDs, including horizontal and vertical pleiotropy. The PLACO method assesses horizontal pleiotropy by evaluating shared risk variants across these diseases at the SNP level. Among the 679 pleiotropic loci annotated from 23,300 pleiotropic SNPs across 36 trait pairs, 345 (50.8%) revealed novel associations with ADs and 354 (52.1%) with CVDs. Several genetic loci, including 17q12 (*GSDMB*), 3p21.31 (*RNF123*), and 12q24.11-q24.12 (*SH2B3*, *ATXN2*, *BRAP*, *ALDH2*), play critical roles in the interplay between ADs and CVDs. For example, Gasdermin B (*GSDMB*) at 17q12, part of the gasdermin family, is instrumental in pyroptosis—a form of cell death integral to inflammatory responses. Dysregulation of pyroptosis can cause severe inflammation, leading to significant tissue damage and organ dysfunction^17^, factors central to the pathogenesis of ADs such as T1D, IBD, and PSC^18^. Additionally, these mechanisms potentially contribute to cardiovascular diseases through pathways linked to atherosclerosis^19^. Another pivotal protein, RING finger protein 123 (*RNF123* or *KPC1*), targets *SOCS1* for proteasomal degradation, thereby enhancing TLR3/IRF7-mediated type 1 interferon (IFN) expression. While essential for protective immunity, dysregulated IFN expression can induce inflammatory tissue damage. Elevated levels of IFN-α and IL-6 in the colonic mucosa of patients with UC and CD exemplify such pathophysiological implications^20^. Moreover, an IFN-inducible transcriptional signature in children at risk of T1D and pronounced IFN signatures in lupus-related cardiovascular conditions underscore the critical role of type 1 IFN in disease progression. In lupus, type 1 IFN contributes to early atherosclerosis by promoting T-cell migration into arterial walls, macrophage recruitment, and foam cell formation^21^.

Our gene-level pleiotropy analysis utilized two mapping strategies: positional mapping and eQTL mapping. This approach identified extensive co-inheritance across 338 genes, representing 87.1% of the 388 critical pleiotropic genes detected across various trait pairs. Notably, genes located at 12q24.11-q24.12, such as *BRAP*, *ATXN2*, *SH2B3*, and *ALDH2*, as well as *GSDMB* at 17q12 and *RNF123* at 3p21.31, have shown significant pleiotropy, influencing over ten trait pairs each, underscoring their profound impact on genetic associations. For example, the *BRAP* gene (*BRCA-1* associated protein gene) is implicated in cardiovascular disease risk via its interaction with the IKK signalosome, which enhances NF-κB nuclear translocation and triggers the transcription of inflammatory cytokines^22^. Similarly, Human Ataxin-2 (*ATXN2*), a conserved RNA-binding protein, regulates the endocytosis of trophic receptors and growth pathways, affecting mitochondrial precursor proteins and metabolic enzymes. Extensive research links *ATXN2* was linked with ADs (T1D, IBD) and CVDs (Stroke, HF, CAD)^23–25^. Lack of *ATXN2* results in reduced levels of the insulin receptor (*INSR*) in the liver and brain, elevated insulin levels in the pancreas and serum, and an excess accumulation of glycogen and fat—key factors contributing to insulin resistance and cardiovascular disease. However, the specific mechanisms through which *ATXN2* influences inflammation or autoimmune diseases remain elusive and are critical areas for further research.

The 12q24.11-12 locus shows pleiotropic effects across all related trait pairs, except ADs-AF and SLE-CAD, encompassing genes such as *SH2B3*, *ATXN2*, *BRAP*, and *ALDH2*. These genes demonstrate robust linkage evidence through both positional and eQTL mapping. Notably, the SNP rs10744777 within this locus acts as an eQTL for aldehyde dehydrogenase-2 (*ALDH2*) in monocytes. *ALDH2* mitigates ischemia-reperfusion injury in rat models and *H9C2* cells under hypoxia-reoxygenation by downregulating *PINK1* and *PRKN* expression, highlighting its protective role in CVDs^26–28^. The *ALDH2*\*1/*2 genotype, associated with various leukopenias, may reduce the risk of ADs^29^. Overexpression of *ALDH2* in human peripheral blood mononuclear cells bolsters oxidative stress resistance by metabolizing 4-HNE and diminishing intracellular reactive oxygen species (ROS), crucial in maintaining redox homeostasis. Such overexpression is posited to protect against ADs by attenuating oxidative stress, a primary factor in chronic inflammation. Additional research links differential *ALDH2* expression to increased CD8+ T cell infiltration, exacerbating apoptosis, adverse ventricular remodeling, and myocardial function deterioration^30^. Another gene, *SH2B3*, is implicated in autoimmune diabetes in adults and CAD through mechanisms involving B-cell proliferation and eosinophil counts, which may induce cardiomyocyte death^31^. The SNP rs3184504 in *SH2B3* exhibits significant pleiotropic effects associated with both immunological disorders and CVDs. Specifically, the rs3184504*A risk allele intensifies activation of the NOD2 recognition pathway, prompting a pro-inflammatory response and disrupting insulin signaling, potentially leading to cardiac injury^32^.

At the pathway level, functional analyses of pleiotropic loci between ADs and CVDs have pinpointed genes that modulate intracellular signal transduction, mainly through the regulation of the MAPK cascade and the PI3K/AKT pathway. The Mitogen-Activated Protein Kinase (MAPK) cascade, essential for cellular functions such as proliferation, differentiation, and apoptosis, incorporates critical kinases like ERK, JNK, and p38. Each kinase has a unique role in cellular signaling. Dysregulation of this cascade can lead to overactive T-cells and excessive release of inflammatory cytokines such as IL-2, IFN-γ, and TNF-α, highlighting its pivotal role in the progression of autoimmune diseases. For example, elevated MAPK activity correlates with increased T-cell activity and heightened autoantibody production in conditions like RA or SLE. Moreover, the overstimulation of the MAPK pathway contributes to the development of atherosclerosis by promoting dysfunction in endothelial cells and fostering inflammation within vascular walls. Proposed molecular mechanisms for MAPK-mediated atherosclerosis suggest that oxidative stress and pro-inflammatory cytokines, like TNF-α and IL-1β—often instigated by hypertension, hyperlipidemia, and smoking—play a central role. Activation of the MAPK pathways can intensify these inflammatory responses, further impairing endothelial function and enhancing vascular damage. Overall, the dysregulation of the MAPK cascade represents a significant factor in perpetuating the autoimmune response and development of atherosclerosis by promoting endothelial cell dysfunction and vascular inflammation, highlighting the potential for novel therapeutic strategies targeting this pathway to treat ADs and CVDs.

Our MR analysis further explored the potential causality between ADs and CVDs, shedding light on the role of vertical pleiotropy in their shared genetic architecture. Using the latest GWAS summary data, we identified robust evidence for the causal effects in either direction of ADs and CVDs across 10 of 36 trait pairs. Notably, RA and UC both demonstrated a positive causal influence on VTE, contrasting with prior research supporting a causal link between UC and VTE risk but did not establish such an association for RA^33^. Interpreting MR estimates involves complexity, as ADs and CVDs are time-varying exposures with considerable polygenic effect variation. Moreover, variability in cohort characteristics, such as disease prevalence and diagnostic criteria, complicates these analyses further. Our results suggest that previous studies may have yet to accurately capture the associations between ADs and CVDs, potentially due to reverse causation, surveillance bias, or unaccounted confounding factors.

Our study acknowledges several limitations. Firstly, our analysis was restricted to GWAS summary data from European ancestry due to constraints in available sample sizes. Although using a consistent pedigree enhances the accuracy of LD score regression, expanding the scope to include cross-ancestry samples and developing statistical methods to assess the generalizability of findings across different ancestry groups are crucial for future research. Secondly, our study focused only on six major ADs and six major CVDs, chosen based on the accessibility and sufficiency of data to ensure adequate statistical power for detecting cross-disorder effects. While these conditions represent a significant portion of the genetic risk architecture for these diseases, our selection needs to be more comprehensive. Expanding the sample size to encompass similar diseases is essential for a deeper understanding of their genetic bases. Finally, our analysis exclusively concentrated on common variants, which are more prone to pleiotropy than rare variants. It was exclusively focused on common variants, which are more prone to pleiotropy than rare ones. Including rare variants, often more specifically associated with particular diseases, along with other types of genetic variations such as undetected SNPs or genetic interactions, could provide a more comprehensive understanding of disease risk.

A large number of previous epidemiological studies have provided evidence of cardiovascular events caused by autoimmune diseases, which raises awareness of the interface between cardiology and rheumatology, which is the field of ‘cardiorheumatology.’ Our study of the genetic associations of ADs and CVDs is more comprehensive genetic research that supplements the knowledge as an overview of the latest advances in ‘cardiorheumatology’ and indicates shared SNP, genes, pathways, and drug targets. We highlighted the importance of early intervention to prevent long-term damage based on the broad shared genetic background and provided new hope for more forward-looking disease management and effective treatments. Therefore, to prevent the onset and progression of cardiovascular events in patients with ADs, several critical clinical interventions are recommended. Routine monitoring of cardiovascular risk factors— including blood pressure, lipid profiles, and glucose levels and annual cardiovascular evaluations to detect early changes. Systemic inflammation is pivotal in damaging the circulatory system but can be effectively managed with disease-modifying antirheumatic drugs (DMARDs) or biological therapies. Patients should also be informed about their increased risk of CVDs linked with ADs and the importance of proactive management. Moreover, it is essential to foster collaboration among rheumatologists, cardiologists, and primary care physicians to provide comprehensive care for these patients.

Our study illuminates the complex genetic relationships that involve various horizontal pleiotropic variants, loci, genes, and pathways throughout the genome. We present robust evidence of shared genetic associations across several loci, notably including SH2B3 and ALDH2 at the 12q24.12 locus. We also identified common biological mechanisms, such as the regulation of the MAPK cascade and the PI3K/AKT signaling pathway, that may contribute to the comorbidities observed between ADs and CVDs. Furthermore, SH2B3, ATXN2, ALDH2, and their functionally related genes have been identified as crucial therapeutic targets for both ADs and CVDs. This study maps the shared genetic foundations of ADs and CVDs and elucidates the mechanisms underlying their comorbidity from a genetic standpoint, offering new insights into the genetic patterns and clinical treatments of these conditions.

## Materials and Methods

### Data sources

Due to differences in linkage disequilibrium (LD) structures across various ancestries, the study cohorts were restricted to individuals of European descent to maintain consistency in genetic analysis. We used the latest and largest GWAS summary statistics for six major ADs and six major CVDs from individuals of European ancestry for each trait (Table 1, Supplementary Table 1).

**Table 1.**
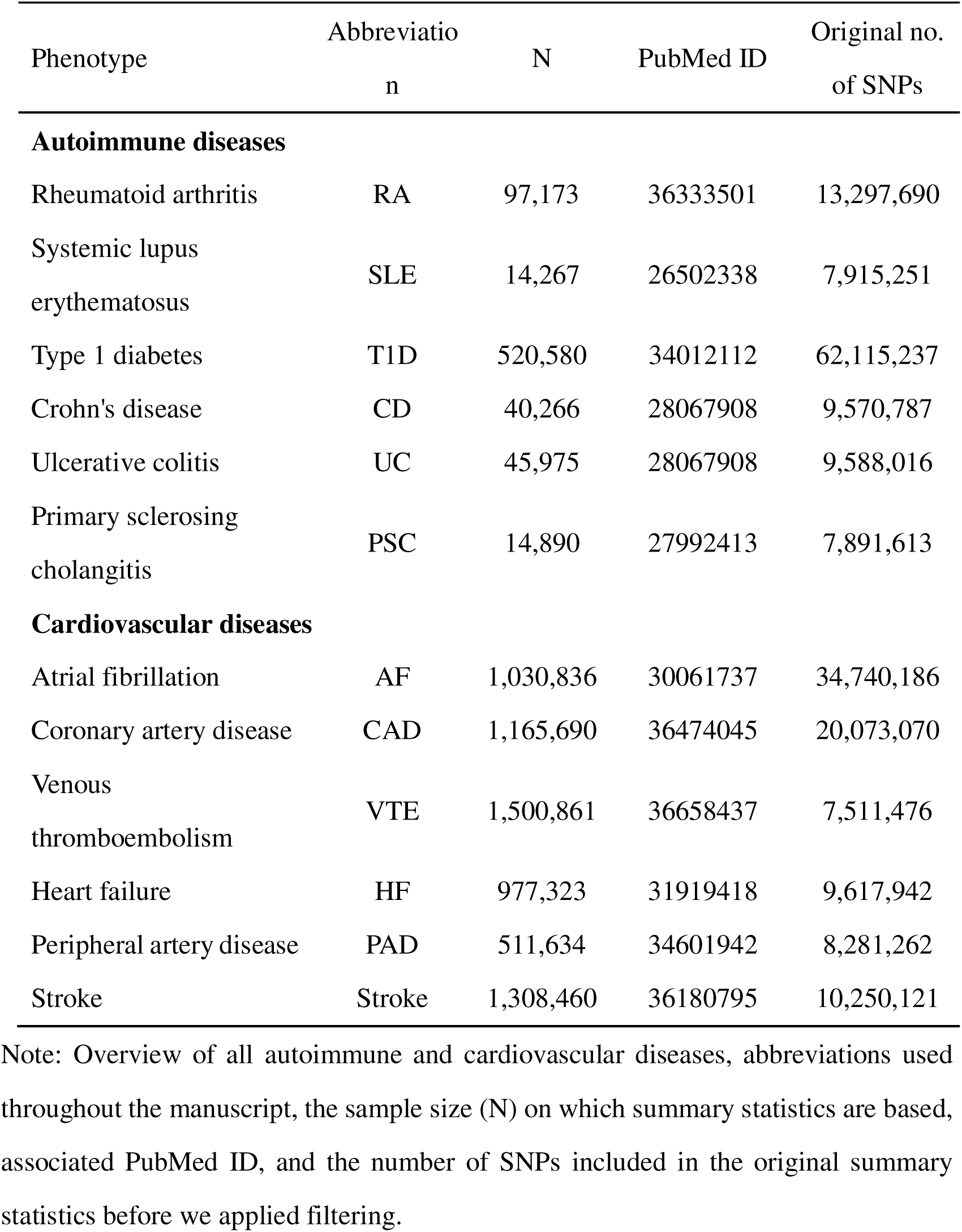
Overview of all autoimmune and cardiovascular diseases included in this study.

GWAS summary statistics for rheumatoid arthritis (RA) were derived from a meta-analysis that included 37 cohorts from Europe, East Asia, Africa, South Asia, and the Arab regions. In this study, we utilized GWAS summary statistics derived solely from individuals of European ancestry, encompassing 22,350 cases and 97,173 controls^34^. GWAS summary statistics for systemic lupus erythematosus (SLE) involved the European subset of the meta-analysis, totaling 5,201 cases and 9,066 controls^35^. GWAS summary statistics for type 1 diabetes (T1D) were obtained from a meta-analysis of 9 large GWAS focusing on European ancestry populations, which included 18,942 cases and 501,638 controls^36^. The international IBD genomics consortium provided summary statistics for Crohn’s disease (CD) and ulcerative colitis (UC) on a population of European Ancestry, comprising 25,042 clinically ascertained cases (12,194 Crohn’s disease and 12,366 ulcerative colitis), and 34,915 controls^37^. The IPSCSG consortium provided summary statistics for primary sclerosing cholangitis (PSC), including 2,871 cases and 12,019 controls^34,38^.

We obtained the largest GWAS summary statistics for atrial fibrillation (AF) from a large-scale meta-analysis involving six studies (The Nord-Trøndelag Health Study [HUNT], deCODE, the Michigan Genomics Initiative [MGI], DiscovEHR, UK Biobank, and the Atrial Fibrillation Genetics [AFGen] Consortium), comprising 60,620 cases and 970,216 controls^39^. GWAS summary statistics for coronary artery disease (CAD) were derived from a meta-analysis of two large GWAS, specifically from the CARDIoGRAMplusC4D Consortium and the UK Biobank, totaling 181,522 cases and 984,168 controls of European ancestry^40^. For venous thromboembolism (VTE), we acquired the most extensive summary statistics from a meta-analysis of seven cohorts, including the Copenhagen Hospital Biobank Cardiovascular Disease Cohort (CHB-CVDC), Danish Blood Donor Study (DBDS), deCODE, Intermountain Healthcare, UK Biobank, FinnGen, and the Million Veterans Program Consortium, involving 81,190 cases and 1,419,671 controls^41^. The Heart Failure Molecular Epidemiology for Therapeutic Targets (HERMES) consortium provided GWAS summary statistics for heart failure (HF), combining data from 26 cohort-level studies with 47,309 cases and 930,014 controls^42^. GWAS summary statistics for PAD were extracted from a meta-analysis of 11 independent GWASs comprising 12,086 patients with PAD and 499,548 control participants of European descent^43^. GWAS summary statistics for Stroke from populations of European Ancestry included 73,652 cases identified by ICD codes and 1,234,808 controls^44^.

GWAS summary statistics for ADs and CVDs underwent quality control procedures: (1) alignment with the 1000 Genomes Project v3 European reference for the hg19 genome assembly; (2) exclusion of non-autosomal single nucleotide polymorphisms (SNPs); (3) removal of SNPs that either lack an rs label or are duplicates; and (4) retention of only biallelic SNPs with a minor allele frequency (MAF) greater than 0.01. After these quality control measures, we rigorously screened all summary statistics to ensure uniformity across the datasets. Subsequent analyses included only the 4,286,675 SNPs common to all 12 diseases studied. Table S1 details sample sizes, the number of SNPs in the original summary statistics prior to filtration, and other pertinent information. All GWAS were approved by relevant ethics committees, and written informed consent was obtained from all participants.

### Heritability and genome-wide genetic correlation analysis

To probe the shared genetic architecture for the genome-wide level, we employed cross-trait linkage disequilibrium (LD) score regression (LDSC) to evaluate SNP-based heritability (*h^2^_SNP_*) for each trait and to calculate genome-wide genetic correlations (*r_g_*) between 6 major ADs and 6 major CVDs. LDSC estimates trait heritability and correlations by analyzing GWAS summary data and LD patterns while mitigating confounding factors and population stratification^45^. First, we used univariate LDSC to estimate SNP-based heritability (*h_SNP_^2^*) for each trait, utilizing SNPs from the 1000 Genomes Project Phase 3 European population as the LD reference, excluding the major histocompatibility complex (MHC) region (chr 6: 25-35 Mb). Subsequently, we applied bivariate LDSC to estimate the genetic correlation between traits. Bivariate LDSC calculates the genetic correlation between two traits by multiplying the z-statistic for each variant’s association with both traits and regressing this product against LD scores. The resulting slope (coefficient) indicates the genetic correlation, which ranges from -1 (indicating opposite influences) to +1 (indicating identical influences) for shared genetic variants influencing the traits. A significant slope indicates a strong genetic correlation, while a non-significant slope suggests little to no genetic correlation. The significance threshold was set using Bonferroni correction at *P* < 1.39×10^-3^ (.05/36).

To assess genome annotation’s contribution to trait heritability, we conducted stratified LDSC applied to specifically expressed genes (LDSC-SEG) to evaluates SNP heritability enrichment across tissue-specific gene expression and cell types relevant to diseases. LDSC-SEG uses genotyping and gene expression reference datasets to identify tissue and cell types significantly enriched for variants contributing to the heritability of a trait. The 53 tissue and cell type-specific expression data from the Genotype-Tissue Expression (GTEx) project and 152 from Franke Lab were analyzed jointly, and tissue and cell type-specific chromatin-based annotations from peaks for 6 epigenetic marks, including 93 labels from Encyclopedia of DNA Elements (ENCODE) EN-TEx and 396 from Roadmap Epigenomics database were used respectively for validation. We adjusted the *P*-values for significance using the false discovery rate (FDR) method, with a FDR threshold set at < 0.05 to determine statistical significance for enrichment.

### Polygenic overlap analysis

Genetic correlation reflects the average shared signal across the genome,^46^, which may not apply when genetic effects combine both the same and opposite directions of effect. To accurately quantify genetic overlap beyond genome-wide significance and delineate the unique and shared genetic architectures of two traits, we employed the causal mixture model (MiXeR) to evaluate the total number of shared and unique variants influencing traits (i.e., variants with pure genetic effects not induced by LD)^47^. By estimating the total number of shared genetic variants, MiXeR identifies polygenic overlap (i.e., shared genetic architecture among common variants) beyond what is captured by genetic correlations, regardless of their effect directions. First, univariate MiXeR analyses were performed for each trait to calculate the SNP-based heritability (*h^2^_SNP_*) and polygenicity, defined as the number of genetic variants responsible for 90% of the SNP heritability. The LD structure was established using the genotype reference panel from Phase 3 of the 1000 Genomes Project. For MiXeR analyses, the MHC region was excluded, following standard recommendations. Subsequently, univariate MiXeR analyses construct a bivariate mixture model for pairs of phenotypes: (1) variants not linked to either phenotype, (2) variants impacting solely the first trait, (3) variants impacting solely the second trait, and (4) variants impacting both traits. The proportion of shared SNPs between two phenotypes relative to their total influence was estimated using Dice coefficients and the proportion of variants with matching effects. MiXeR further calculated the genome-wide correlations across all SNPs (*r_g_*) and the correlation of effect sizes within the shared genetic component (*r_g_s*). MiXeR uses the Akaike Information Criterion (AIC) to evaluate model fit by comparing its model to the infinitesimal model. A positive AIC difference supports the MiXeR model of polygenic overlap, indicating that the GWAS data have sufficient power to distinguish the estimated polygenic overlap from models with minimal or maximal overlap.

### Local heritability and genetic correlation analysis

To determine the presence of shared genetic correlation within independent genomic regions, we calculated the local genetic correlations using the Local Analysis of [co]Variant Association (LAVA) method^46^. Employing LAVA allowed us to examine the direction of correlation at each genomic locus, offering a deeper comprehension of the genetic overlap between traits. LAVA complements MiXeR by estimating local genetic correlations (*r_gS_*) across 2,495 semi-independent loci, each around 1 megabase (Mb) in size, which effectively identifies regions with mixed effect directions, even when the overall *r_g_* is minimal. Initially, LAVA conducted univariate tests on each trait and locus to select those with significant local genetic signals, with a *P*-value < 1×10^−4^ as significant. The LD reference panel was used based on the 1000 Genomes Phase 3 European genotype data, excluding complex MHC region. Subsequent bivariate tests were conducted on the selected loci and traits to investigate their local genetic correlations. We used multiple testing corrections to adjust for the number of genomic loci tested in local bivariate analyses with a Bonferroni-corrected significant threshold set at *P*_<_5.43×10^-6^ (.05 / 9203). LAVA accounts for sample overlap by incorporating the genetic covariance intercept from LDSC.

### Pleiotropic analysis

All these approaches above describe genetic sharing, but genetic sharing at the locus level or novel shared variants/loci is not indicated. To identify pleiotropic effects of genetic variants (i.e., horizontal pleiotropy) underlying the genetic correlations and detect potential pleiotropy at the SNP level, we further extended our analysis using the pleiotropic analysis under the composite null hypothesis (PLACO) to identify shared genetic SNPs with concordant or discordant effect directions between the two phenotypes. PLACO considers a composite null hypothesis, positing that a variant is linked to either none or just one of the traits, thereby enhancing the specificity of genetic correlations identified across studies^48^. Therefore, rejecting this composite null hypothesis suggests the presence of pleiotropy, where both phenotypes are associated with the variant. PLACO implements this test using the product of the Z-statistics corresponding to each trait as the test statistic. The null distribution of this statistic is modeled as a mixture distribution, which accounts for the possibility that the variant may not be linked to either or just one of the traits. SNPs with *P_PLACO_*< 5×10^-8^ were identified as significant pleiotropic variants.

### Loci definition and functional annotation

Functional Mapping and Annotation (FUMA) was used to discover independent genomic loci and annotate GWAS results, helping to clarify the shared genetic influences between two traits. FUMA combines data from various biological resources to annotate GWAS results, prioritize genes, and offer interactive visualizations.^49^. Based on the pre-calculated LD structure from the 1000 Genomes European reference panel, SNPs with *P* < 5 × 10^−8^ and r^2^ < 0.6 within 1 Mb were defined as independent significant SNPs. Independent lead SNPs were identified as those with low LD (r^2^ < 0.1) with other SNPs. LD blocks with significant SNPs within 500 kb were combined into a single genomic locus, with the top SNP being the one with the smallest P value in that region. The effect directions were interpreted by comparing the Z-scores of lead SNPs for each locus in the GWAS summary statistics for the trait. A locus was deemed novel to ADs and CVDs if it did not physically overlap with the loci in the original GWAS. ANNOVAR was used to evaluate the proximity of Lead SNPs to genes and their potential effects on gene function through functional annotation. We then utilized Combined Annotation Dependent Depletion (CADD) scores to assess the harmful effects of the SNP on protein function and RegulomeDB scores to predict the regulatory role of the SNP that reflected functions based on expression quantitative trait loci (eQTLs) and chromatin markers. SNPs with a CADD score above 12.37 were classified as possibly harmful, and RDB assigns a score ranging from 1 to 7, where 1 indicates substantial proof of being a regulatory variant and 7 indicates minimal evidence. Additionally, we employed position mapping and cis-eQTL mapping to determine how shared risk loci impact the genes functionally. Gene annotations for each locus are defined by their proximity to the most significant/lead SNPs identified by FUMA. Positional mapping linked shared independent lead SNPs with protein-coding genes based on their proximity within 10 kb in the human reference assembly (GRCh37/hg19). Functional implications of the identified lead variants were evaluated using eQTL mapping.

### Colocalization analysis

To identify potential shared causal variants in each pleiotropic locus, we utilized COLOC to perform colocalization analyses. COLOC utilizes regression coefficients for each SNP and the variance of these coefficients for each trait to assess the probability that the two traits share a common genetic causal variant. COLOC evaluates five posterior probabilities (PPs), each representing a distinct hypothesis about the genetic association with the traits under study: H_0_ indicates no association with either trait; H_1_ and H_2_ suggest an association with one of the traits; H_3_ implies that both traits are associated, but with different causal variants; and H_4_ suggests a shared causal variant influences both traits. The analyses employed default COLOC prior probabilities: p_1_ and p_2_ were each set at 1×10^-4^ for an SNP’s association with the first and second traits, respectively, and p_12_ at 1×10^-5^ for an SNP associated with both traits. A locus was considered colocalized if the posterior probability of H_4_ (PP.H4) exceeded 0.7, with the SNP showing the highest PP.H4 identified as the candidate causal variant.

### Gene-level analysis

To further explore candidate pleiotropic genes, gene-level Multi-marker Analysis of GenoMic Annotation (MAGMA) was employed on genes situated within or overlapping the pleiotropic loci, as identified by PLACO results and single-trait GWAS analyses. MAGMA aggregates SNP-level associations into a single gene-level association signal by first analyzing the individual SNPs in a gene and combining the resulting SNP *P*-values into a gene test statistic. It can thus be used to calculate a *P*-value for each gene, considering factors such as gene size, SNP count per gene, and LD among the markers. SNPs physically located within the gene body or extending to a 10 KB window around each gene were assigned to genes. SNPs were annotated to genes based on the 1000 Genomes Project Phase 3 European population as the reference panel and the human genome Build 37 (GRCh37/hg19) locations for 17,636 protein-coding genes as primary proteins analyzed. The MHC region (chr6: 25-35 Mb) was excluded from the MAGMA analyses for all trait pairs under consideration. We additionally implemented multiple testing corrections to adjust for the number of unique protein-coding genes and the number of trait pairs (threshold: *P*_=_0.05 / no. of proteins_/ no. of trait pairs =_0.05 / 17,636 / 36 = 7.88×10^-8^).

Shared genetic risk variants frequently influenced gene expression in a tissue-specific manner, as evidenced by the expression of quantitative trait loci (eQTLs). MAGMA often struggles to identify functionally relevant genes as it assigns SNPs to the nearest genes, which can overlook distal regulatory effects on gene expression mediated by eQTLs. To explore the biological mechanisms underlying pleiotropic loci across 36 trait pairs more thoroughly, we conducted tissue-specific gene analyses using eQTL-informed MAGMA (e-MAGMA), which utilizes PLACO results. E-MAGMA uses tissue-specific eQTL data to assign risk variants to genes, more accurately reflecting the functional relationships between SNPs and cis-eQTLs within specific tissues^50^. We employed gene expression data from the Genotype-Tissue Expression Project (GTEx) v8, selecting 17 tissues based on previous studies and findings from LDSC-SEG analyses, highlighting their relevance to immune and cardiovascular diseases and the clinical manifestations in the affected organs. We excluded non-coding and duplicated genes from each selected tissue. Our LD reference data was derived from the 1000 Genomes Phase 3 European panel. As with MAGMA, results from e-MAGMA analysis within the MHC region (chr6: 25-35 Mb) were omitted to avoid confounding due to complex LD structures. Tissue-specific *P*-values were computed for each gene across the selected tissues, applying a Bonferroni correction based on the number of tissue-specific protein-coding genes and trait pairs analyzed. For example, the significance threshold for Whole Blood was set at *P* = 2.25×10^-7^ (*P* = 0.05/number of tissue-specific genes/number of trait pairs = 0.05 / 6,162 / 36).

Additionally, we supplemented and validated the results from the e-MAGMA analyses through Transcriptome-Wide Association Scanning (TWAS) using single-trait GWAS results. TWAS utilizes the LD Expression Reference Panel to discern gene-trait associations from GWAS datasets, effectively minimizing the impact of environmental and technical variables on gene expression. Initially, we employed FUSION to calculate tissue-specific gene expression using various prediction models. We then selected gene expression weights from the model demonstrating the best statistical performance and integrated these with GWAS statistics to pinpoint significant associations between gene expression levels and traits. TWAS utilized the same tissues from the GTEx v8 dataset as those assessed in the e-MAGMA study, applying tissue-specific Bonferroni corrections to ensure rigorous determination of statistical significance.

### Pathway enrichment analysis

Gene-set analysis offers a further understanding of the functional and biological processes that contribute to the genetic component of a trait. We performed gene-set enrichment analysis using MAGMA, which employs a regression structure that facilitates the analysis of continuous gene properties and enables the simultaneous analysis of multiple gene sets and other gene attributes. MAGMA initially quantified the association of each gene with the phenotype and estimated correlations between genes. Subsequently, the analysis utilized the gene *P*-values and the gene correlation matrix to perform the gene-set analysis. The Canonical Pathways from the MSigDB database were used for the gene-set analysis, and focused enrichment tests were performed on Gene Ontology biological processes (GO_BP) and Reactome pathway based on genes identified from MAGMA gene-based analysis. Multiple testing was adjusted using a Bonferroni correction, with the threshold set at *P* = 0.05 / (7,744 + 1,654) / 36 = 1.48×10^-7^. Moreover, the Metascape was further utilized to conduct pathway enrichment analysis on genes identified as significant by both MAGMA and e-MAGMA analyses, overlapping multiple trait pairs. Metascape’s functional enrichment analysis allows for identifying biological pathways and processes that are overrepresented in a given gene list, providing insights into the underlying mechanisms of diseases. We employed the default settings of the Metascape, setting the cut-off *P* value as 0.01.

### Drug target analysis

We utilized STRING V.11.5 to query genes associated with ADs and CVDs. STRING assembles protein-protein interaction (PPI) networks, annotating each interaction with a confidence score ranging from 0 to 1, which reflects the strength of both physical and functional associations. We specifically focused on biologically related neighborhood genes, defining them as those with a high confidence score (a combined score excluding ‘text mining score’ greater than 0.7) about our target genes. Subsequent searches in the Drug Gene Interaction Database (DGIdb) assessed whether target genes and their neighboring genes were already recognized as drug targets, facilitating the identification of potential therapeutic interventions.

### Mendelian randomization analysis

Studies on horizontal pleiotropy have underscored potential shared biological mechanisms between trait pairs, yet the causal relationships remain elusive (i.e., vertical pleiotropy). We used the Latent Heritable Confounder MR (LHC-MR) approach to estimate the bidirectional causal effects between ADs and CVDs to address this. LHC-MR estimates trait heritability and calculates bidirectional causal effects using genome-wide variants, accounting for sample overlap. This refined method models potential unmeasured heritable confounders, both genetic and environmental, that may affect both exposure and outcome. By distinguishing between genetic confounders contributing to observed genetic correlations and actual causation, LHC-MR enhances the accuracy of causal effect estimates compared to traditional MR methods. We adjusted the *P*-value for multiple testing for statistical significance, setting a threshold of 6.94×10^-4^ (*P* = 0.05/number of trait pairs/number of tests = 0.05/36/2). To validate the stability of the causal relationship, we employed five MR methods: simple mode, weighted median, MR-Egger, weighted mode, and inverse variance weighting (IVW) to assess the causality between ADs and CVDs, with significance set at *P* < 0.05.

## Data Availability

All data produced in the present study are available upon reasonable request to the authors.

## Data availability

The study used only openly available GWAS summary statistics on six autoimmune diseases and six CVDs that have originally been conducted using human data. GWAS summary statistics on RA, SLE, T1D, CD, UC, and PSC are available at the GWAS Catalog (GCST90132223, GCST003156, GCST90014023, GCST004132, GCST004133, and GCST004030). GWAS summary statistics on AF, HF, and Stroke are available at the GWAS Catalog (GCST90104539, GCST009541, and GCST90104539). GWAS summary statistics on CAD and PAD are publicly available for download at the Cardiovascular Disease Knowledge Portal (CVDKP) website: https://cvd.hugeamp.org/datasets.html. GWAS summary statistics on VTE are obtained from the deCODE genetics website: https://www.decode.com/summarydata/.

## Code availability

All software used to conduct the analyses in this paper are freely available online. Software (version, where applicable) and sources are listed below: LDSC (v1.0.1; https://github.com/bulik/ldsc), MiXeR (v1.3; https://github.com/precimed/mixer), LAVA (v0.1.0; https://github.com/josefin-werme/LAVA), LCV (https://github.com/lukejoconnor/ LCV); LHC-MR (v0.0.0.9000; https://github.com/LizaDarrous/lhcMR), PLACO (v0.1.1; https://github.com/RayDebashree/PLACO), FUMA (v1.5.4; http://fuma.ctglab.nl/), HyPrColoc(v1.0; https://github.com/jrs95/hyprcoloc), MAGMA (v.1.08; https://ctg.cncr.nl/software/magma), e-MAGMA (https://github.com/eskederks/eMAGMA-tutorial), TWAS (http://gusevlab.org/projects/fusion/), SMR (v1.31; https://yanglab.westlake.edu.cn/software/smr/), COLOC (v5.2.1; https://github.com/chr1swallace/coloc), and R (v.4.1.3; https://www.r-project.org/).

## Acknowledgements

This study was supported by the Natural Science Foundation of China Excellent Young Scientists Fund (Overseas) (Grant no. K241141101), National Natural Science Foundation (Grant no. 82470452), Guangdong Basic and Applied Basic Research Foundation for Distinguished Young Scholars (Grant no. 24050000763), Shenzhen Pengcheng Peacock Plan, Shenzhen Basic Research General Projects of Shenzhen Science and Technology Innovation Commission (Grant no. JCYJ20230807093514029) (To Y.F.), Natural Science Foundation of China Excellent Young Scientists Fund (Overseas) (Grant no. K241001101) (To Z.L.), National Natural Science Foundation of China (Grant no. 82300315; 82374240), Guangdong Province Basic and Applied Basic Research Fund Project (Grant no. 2024A1515012174; 2024A1515013184), National Administration of Traditional Chinese Medicine Research Project (Grant no. 0102023703), Project of the State Key Laboratory of Dampness Syndrome of Traditional Chinese Medicine jointly established by the province and the ministry (Grant no. SZ2022KF10), Scientific Research Initiation Project of Guangdong Provincial Hospital of Traditional Chinese Medicine (Grant no. 2021KT1709), Research Project of Guangdong Provincial Bureau of Traditional Chinese Medicine (Grant no. 20241120), Guangdong Provincial Key Laboratory of Research on Emergency in TCM (Grant no. 2023B1212060062; 2023KT15450) (To R.Z.), and Center for Computational Science and Engineering at Southern University of Science and Technology. The funder had no role in the design, implementation, analysis, interpretation of the data, approval of the manuscript, and decision to submit the manuscript for publication.

## Author contributions

J.Q., Y.F., Z.L., R.Z., and S.P. conceptualized and supervised this project and wrote the manuscript. J.Q., M.C., M.C., and Y.Z. performed the main analyses and wrote the manuscript. J.Q., J.H., P.Z., and R.Z. performed the statistical analysis and assisted with interpreting the results. L.C., F.L., and X.F. provided expertise in GWAS summary statistics. All authors discussed the results and commented on the paper.

## Reference

1 Roth GA, Mensah GA, Johnson CO et al. Global Burden of Cardiovascular Diseases and Risk Factors, 1990-2019: Update From the GBD 2019 Study. J Am Coll Cardiol 2020; 76: 2982–3021.

2 Parkin J & Cohen B. An overview of the immune system. Lancet 2001; 357: 1777–1789.

3 Edsfeldt A, Grufman H, Asciutto G et al. Circulating cytokines reflect the expression of pro-inflammatory cytokines in atherosclerotic plaques. Atherosclerosis 2015; 241: 443–449.

4 Tsioufis P, Theofilis P, Tsioufis K et al. The Impact of Cytokines in Coronary Atherosclerotic Plaque: Current Therapeutic Approaches. Int J Mol Sci 2022; 23:

5 Frangogiannis NG. The inflammatory response in myocardial injury, repair, and remodelling. Nat Rev Cardiol 2014; 11: 255–265.

6 Conrad N, Verbeke G, Molenberghs G et al. Autoimmune diseases and cardiovascular risk: a population-based study on 19 autoimmune diseases and 12 cardiovascular diseases in 22 million individuals in the UK. Lancet 2022; 400: 733–743.

7 Leonard D, Svenungsson E, Dahlqvist J et al. Novel gene variants associated with cardiovascular disease in systemic lupus erythematosus and rheumatoid arthritis. Ann Rheum Dis 2018; 77: 1063–1069.

8 Svenungsson E, Gustafsson J, Leonard D et al. A STAT4 risk allele is associated with ischaemic cerebrovascular events and anti-phospholipid antibodies in systemic lupus erythematosus. Ann Rheum Dis 2010; 69: 834–840.

9 van der Harst P & Verweij N. Identification of 64 Novel Genetic Loci Provides an Expanded View on the Genetic Architecture of Coronary Artery Disease. Circ Res 2018; 122: 433–443.

10 Sivakumaran S, Agakov F, Theodoratou E et al. Abundant pleiotropy in human complex diseases and traits. Am J Hum Genet 2011; 89: 607–618.

11 Gao N, Kong M, Li X et al. Systemic Lupus Erythematosus and Cardiovascular Disease: A Mendelian Randomization Study. Front Immunol 2022; 13: 908831.

12 Kain J, Owen KA, Marion MC et al. Mendelian randomization and pathway analysis demonstrate shared genetic associations between lupus and coronary artery disease. Cell Rep Med 2022; 3: 100805.

13 Rong JC, Chen XD, Jin NK et al. Exploring the causal association of rheumatoid arthritis with atrial fibrillation: a Mendelian randomization study. Clin Rheumatol 2024; 43: 29–40.

14 Wang M, Chao C, Mei K et al. Relationship between rheumatoid arthritis and cardiovascular comorbidity, causation or co-occurrence: A Mendelian randomization study. Front Cardiovasc Med 2023; 10: 1099861.

15 Gong W, Guo P, Li Y et al. Role of the Gut-Brain Axis in the Shared Genetic Etiology Between Gastrointestinal Tract Diseases and Psychiatric Disorders: A Genome-Wide Pleiotropic Analysis. JAMA Psychiatry 2023; 80: 360–370.

16 Shirai Y, Nakanishi Y, Suzuki A et al. Multi-trait and cross-population genome-wide association studies across autoimmune and allergic diseases identify shared and distinct genetic component. Ann Rheum Dis 2022; 81: 1301–1312.

17 Rana N, Privitera G, Kondolf HC et al. GSDMB is increased in IBD and regulates epithelial restitution/repair independent of pyroptosis. Cell 2022; 185: 283–298.e217.

18 Das S, Miller M & Broide DH. Chromosome 17q21 Genes ORMDL3 and GSDMB in Asthma and Immune Diseases. Adv Immunol 2017; 135: 1–52.

19 Vasudevan SO, Behl B & Rathinam VA. Pyroptosis-induced inflammation and tissue damage. Semin Immunol 2023; 69: 101781.

20 Huang S, Cui M, Huang J et al. RNF123 Mediates Ubiquitination and Degradation of SOCS1 To Regulate Type I Interferon Production during Duck Tembusu Virus Infection. J Virol 2023; 97: e0009523.

21 Kirchler C, Husar-Memmer E, Rappersberger K et al. Type I Interferon as cardiovascular risk factor in systemic and cutaneous lupus erythematosus: A systematic review. Autoimmun Rev 2021; 20: 102794.

22 Liao YC, Wang YS, Guo YC et al. BRAP Activates Inflammatory Cascades and Increases the Risk for Carotid Atherosclerosis. Mol Med 2011; 17: 1065–1074.

23 Barrett JC, Clayton DG, Concannon P et al. Genome-wide association study and meta-analysis find that over 40 loci affect risk of type 1 diabetes. Nat Genet 2009; 41: 703–707.

24 Liu JZ, van Sommeren S, Huang H et al. Association analyses identify 38 susceptibility loci for inflammatory bowel disease and highlight shared genetic risk across populations. Nat Genet 2015; 47: 979–986.

25 Ehret GB, Munroe PB, Rice KM et al. Genetic variants in novel pathways influence blood pressure and cardiovascular disease risk. Nature 2011; 478: 103–109.

26 Ajoolabady A, Chiong M, Lavandero S et al. Mitophagy in cardiovascular diseases: molecular mechanisms, pathogenesis, and treatment. Trends Mol Med 2022; 28: 836–849.

27 Budas GR, Disatnik MH & Mochly-Rosen D. Aldehyde dehydrogenase 2 in cardiac protection: a new therapeutic target? Trends Cardiovasc Med 2009; 19: 158–164.

28 Pang J, Wang J, Zhang Y et al. Targeting acetaldehyde dehydrogenase 2 (ALDH2) in heart failure-Recent insights and perspectives. Biochim Biophys Acta Mol Basis Dis 2017; 1863: 1933–1941.

29 Yokoyama A, Brooks PJ, Yokoyama T et al. Blood Leukocyte Counts and Genetic Polymorphisms of Alcohol Dehydrogenase-1B and Aldehyde Dehydrogenase-2 in Japanese Alcoholic Men. Alcohol Clin Exp Res 2016; 40: 507–517.

30 Zhang H, Li Z & Zheng Y. Identifying the Therapeutic and Prognostic Role of the CD8+ T Cell-Related Gene ALDH2 in Head and Neck Squamous Cell Carcinoma. Cancer Inform 2022; 21: 11769351221139252.

31 Gudbjartsson DF, Bjornsdottir US, Halapi E et al. Sequence variants affecting eosinophil numbers associate with asthma and myocardial infarction. Nat Genet 2009; 41: 342–347.

32 Zhernakova A, Elbers CC, Ferwerda B et al. Evolutionary and functional analysis of celiac risk loci reveals SH2B3 as a protective factor against bacterial infection. Am J Hum Genet 2010; 86: 970–977.

33 Lv X, Gao X, Liu J et al. Immune-mediated inflammatory diseases and risk of venous thromboembolism: A Mendelian randomization study. Front Immunol 2022; 13: 1042751.

34 Ishigaki K, Sakaue S, Terao C et al. Multi-ancestry genome-wide association analyses identify novel genetic mechanisms in rheumatoid arthritis. Nat Genet 2022; 54: 1640–1651.

35 Bentham J, Morris DL, Graham DSC et al. Genetic association analyses implicate aberrant regulation of innate and adaptive immunity genes in the pathogenesis of systemic lupus erythematosus. Nat Genet 2015; 47: 1457–1464.

36 Chiou J, Geusz RJ, Okino ML et al. Interpreting type 1 diabetes risk with genetics and single-cell epigenomics. Nature 2021; 594: 398–402.

37 de Lange KM, Moutsianas L, Lee JC et al. Genome-wide association study implicates immune activation of multiple integrin genes in inflammatory bowel disease. Nat Genet 2017; 49: 256–261.

38 Ji SG, Juran BD, Mucha S et al. Genome-wide association study of primary sclerosing cholangitis identifies new risk loci and quantifies the genetic relationship with inflammatory bowel disease. Nat Genet 2017; 49: 269–273.

39 Nielsen JB, Thorolfsdottir RB, Fritsche LG et al. Biobank-driven genomic discovery yields new insight into atrial fibrillation biology. Nat Genet 2018; 50: 1234–1239.

40 Aragam KG, Jiang T, Goel A et al. Discovery and systematic characterization of risk variants and genes for coronary artery disease in over a million participants. Nat Genet 2022; 54: 1803–1815.

41 Ghouse J, Tragante V, Ahlberg G et al. Genome-wide meta-analysis identifies 93 risk loci and enables risk prediction equivalent to monogenic forms of venous thromboembolism. Nat Genet 2023; 55: 399–409.

42 Shah S, Henry A, Roselli C et al. Genome-wide association and Mendelian randomisation analysis provide insights into the pathogenesis of heart failure. Nat Commun 2020; 11: 163.

43 van Zuydam NR, Stiby A, Abdalla M et al. Genome-Wide Association Study of Peripheral Artery Disease. Circ Genom Precis Med 2021; 14: e002862.

44 Mishra A, Malik R, Hachiya T et al. Stroke genetics informs drug discovery and risk prediction across ancestries. Nature 2022; 611: 115–123.

45 Bulik-Sullivan BK, Loh PR, Finucane HK et al. LD Score regression distinguishes confounding from polygenicity in genome-wide association studies. Nat Genet 2015; 47: 291–295.

46 Werme J, van der Sluis S, Posthuma D et al. An integrated framework for local genetic correlation analysis. Nat Genet 2022; 54: 274–282.

47 Frei O, Holland D, Smeland OB et al. Bivariate causal mixture model quantifies polygenic overlap between complex traits beyond genetic correlation. Nat Commun 2019; 10: 2417.

48 Ray D & Chatterjee N. A powerful method for pleiotropic analysis under composite null hypothesis identifies novel shared loci between Type 2 Diabetes and Prostate Cancer. PLoS Genet 2020; 16: e1009218.

49 Watanabe K, Taskesen E, van Bochoven A et al. Functional mapping and annotation of genetic associations with FUMA. Nat Commun 2017; 8: 1826.

50 Gerring ZF, Mina-Vargas A, Gamazon ER et al. E-MAGMA: an eQTL-informed method to identify risk genes using genome-wide association study summary statistics. Bioinformatics 2021; 37: 2245–2249.

