## Supplementary Information for "Genetic analyses identify shared genetic components related to autoimmune and cardiovascular diseases"

Associate Professor

School of Public Health and Emergency Management, School of Medicine, Southern University of Science and Technology, Shenzhen, Guangdong, 518055, China.

Rongjun Zou MD, PhD

Professor

Department of Cardiovascular Surgery, Guangdong Provincial Hospital of Chinese Medicine, the Second Affiliated Hospital of Guangzhou University of Chinese Medicine, the Second Clinical College of Guangzhou University of Chinese Medicine, Guangzhou, China;

State Key Laboratory of Dampness Syndrome of Chinese Medicine, Guangzhou, China;

Guangdong Provincial Key Laboratory of TCM Emergency Research, Guangzhou, China.

Siim Pauklin PhD

Group leader and CRUK Career Development Fellow

Botnar Research Centre, Nuffield Department of Orthopaedics, Rheumatology and Musculoskeletal Sciences, University of Oxford, Headington, Oxford OX3 7LD, UK.

**Supplementary Figures**

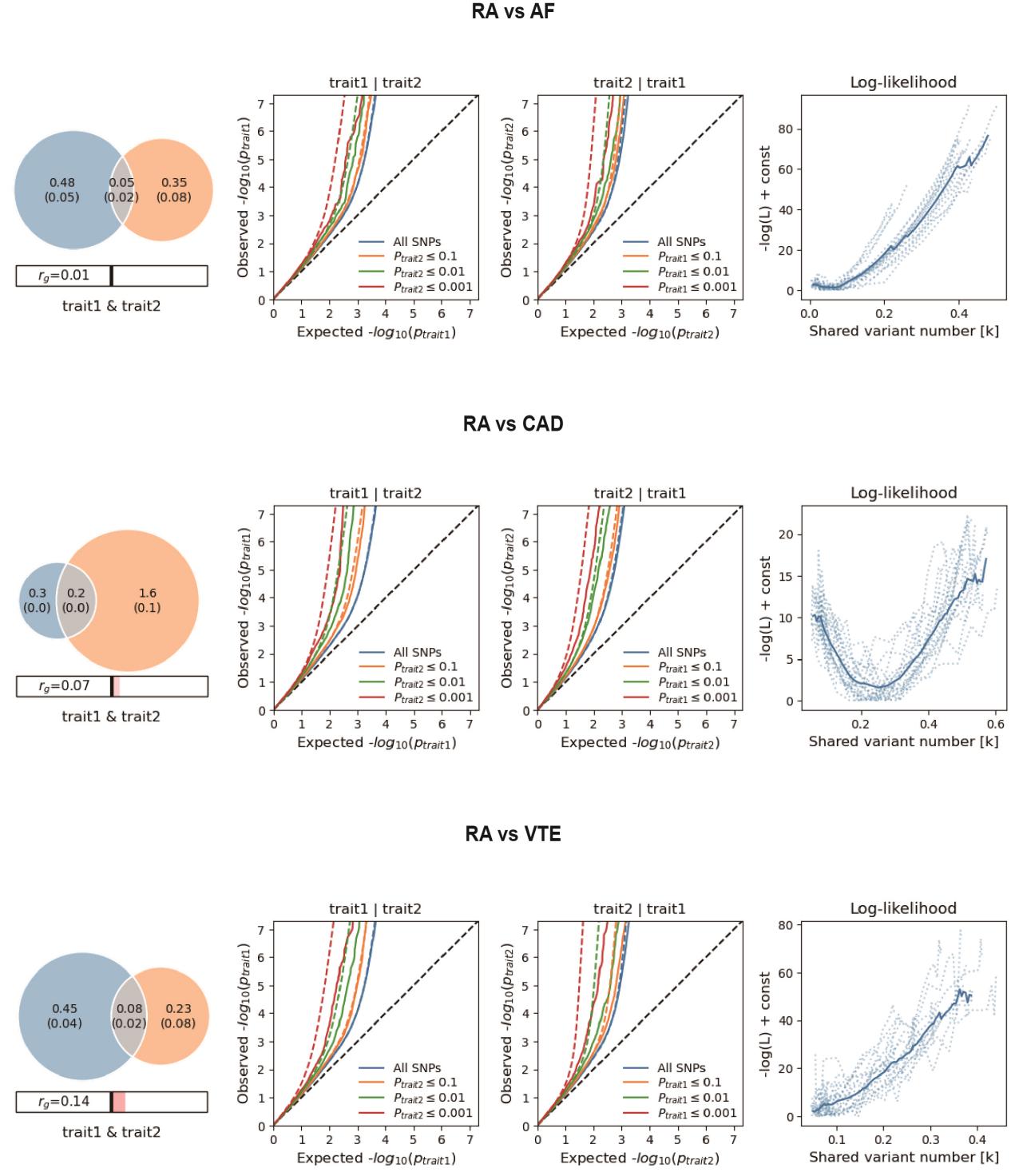

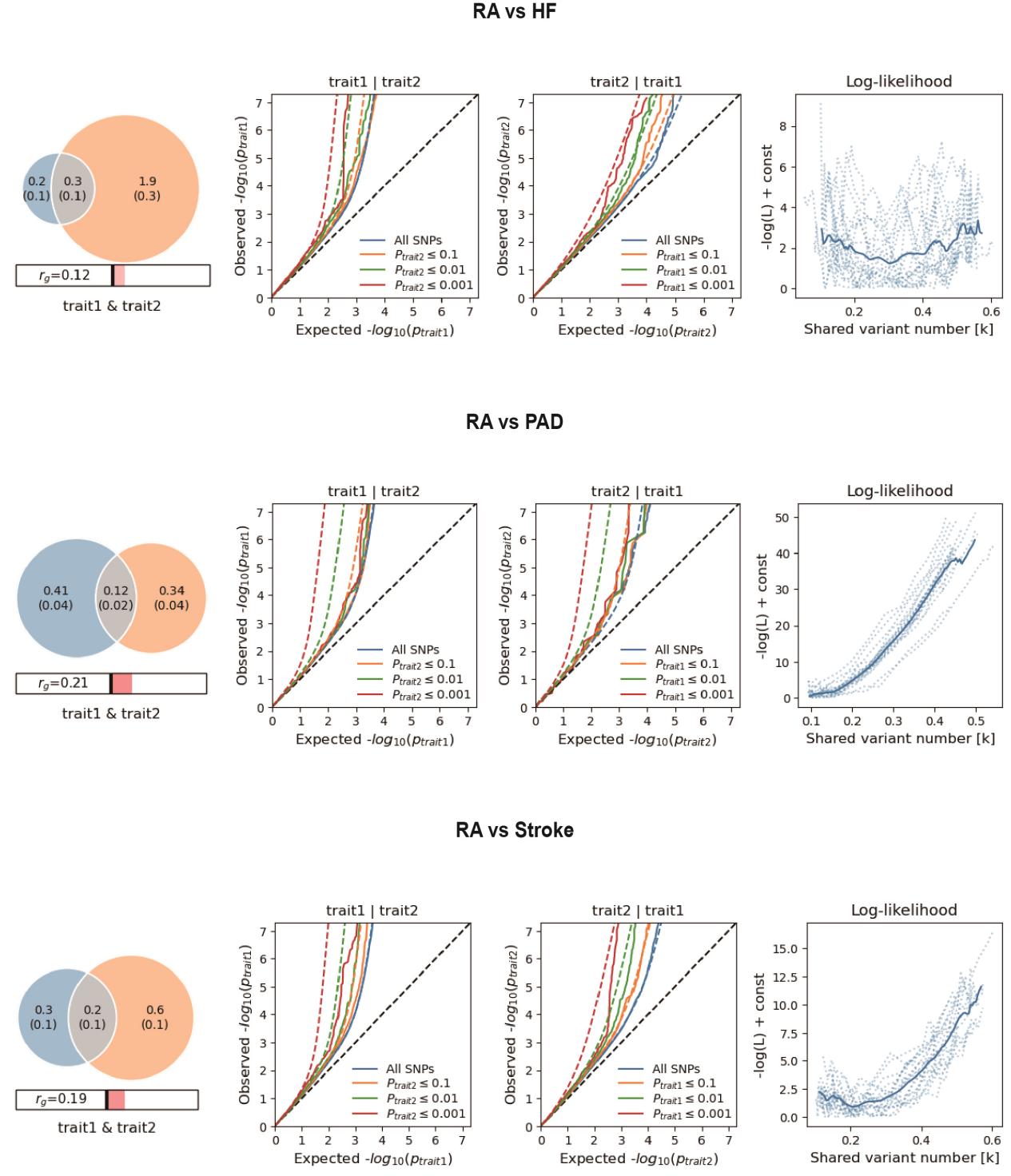

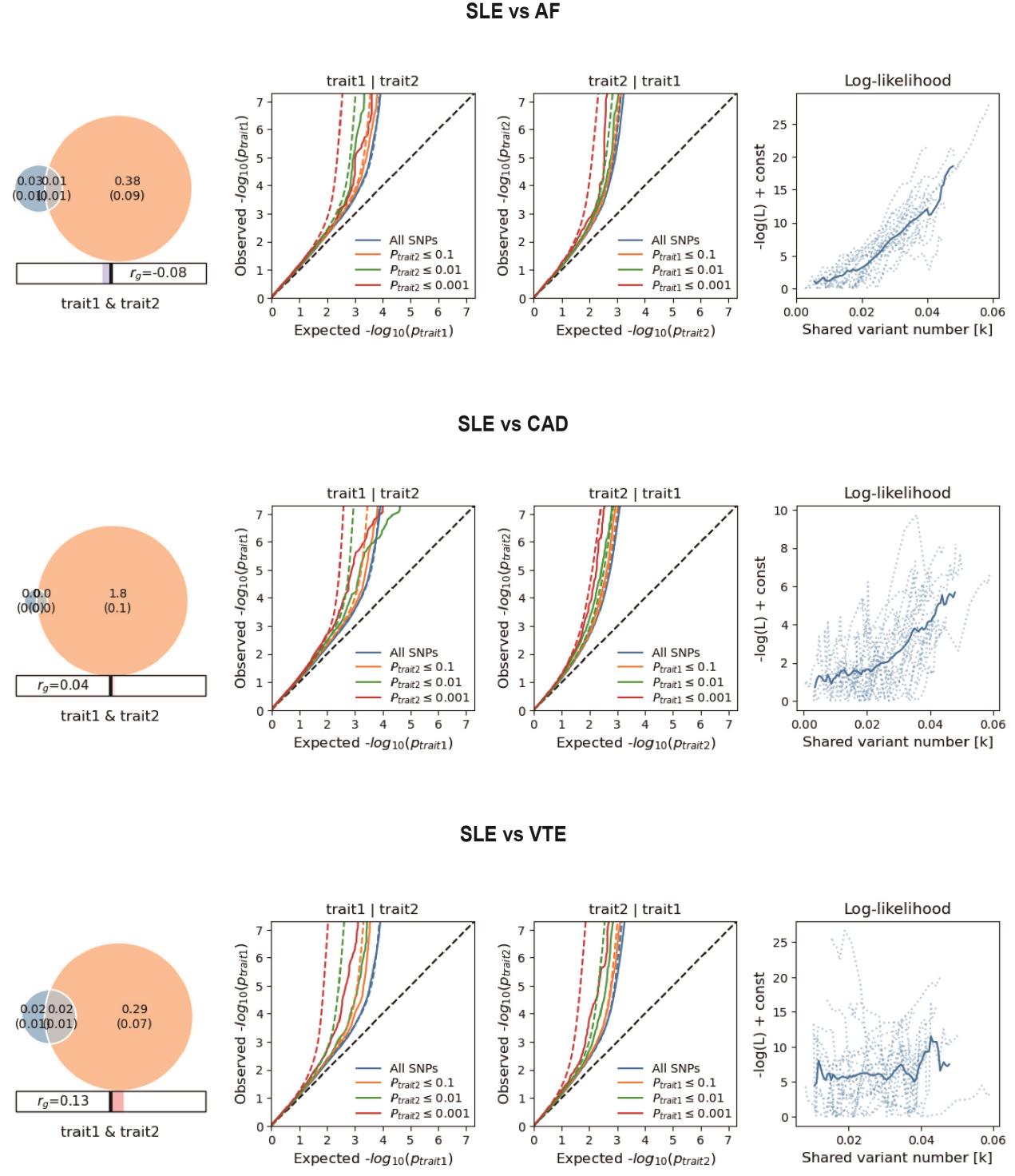

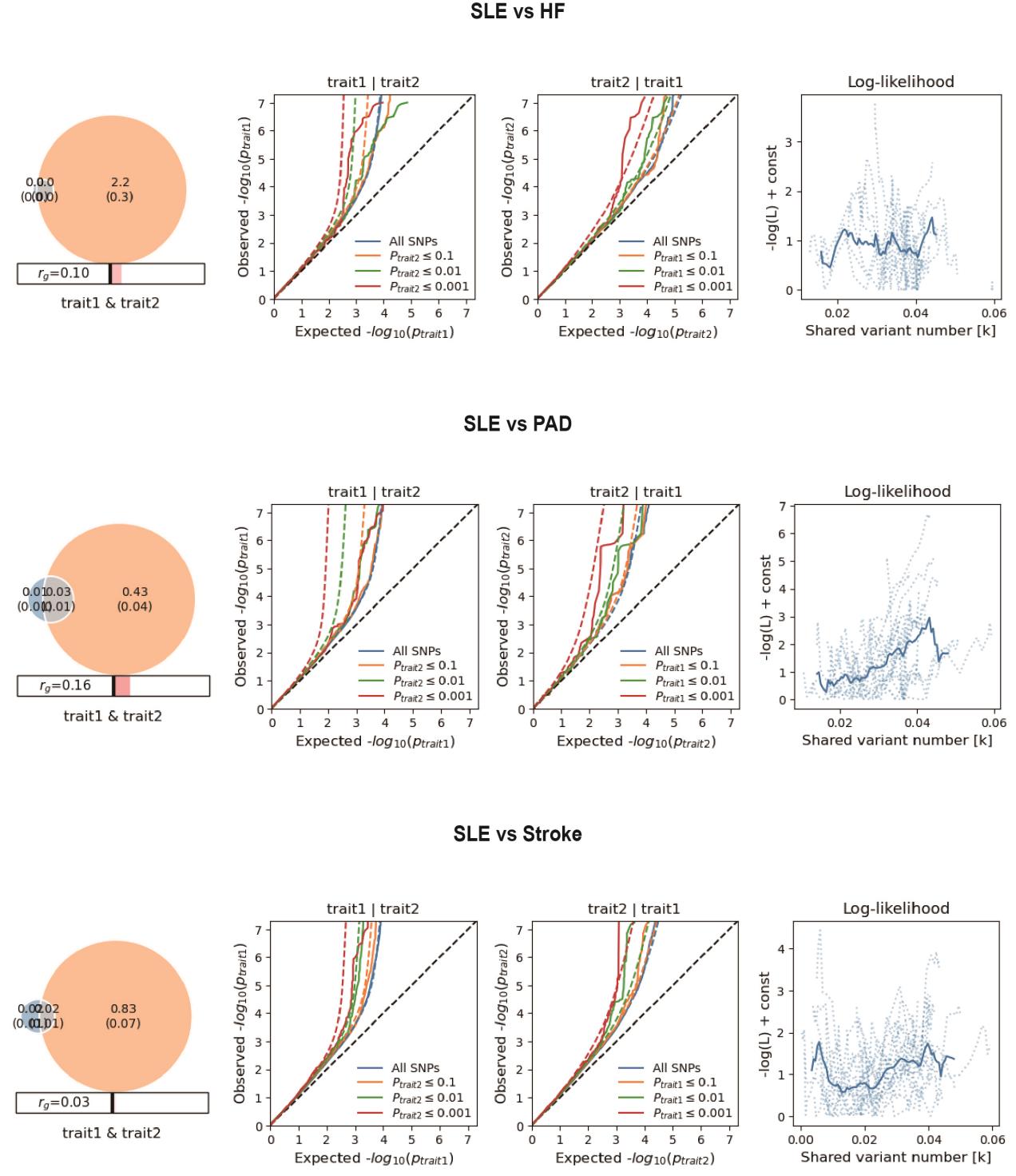

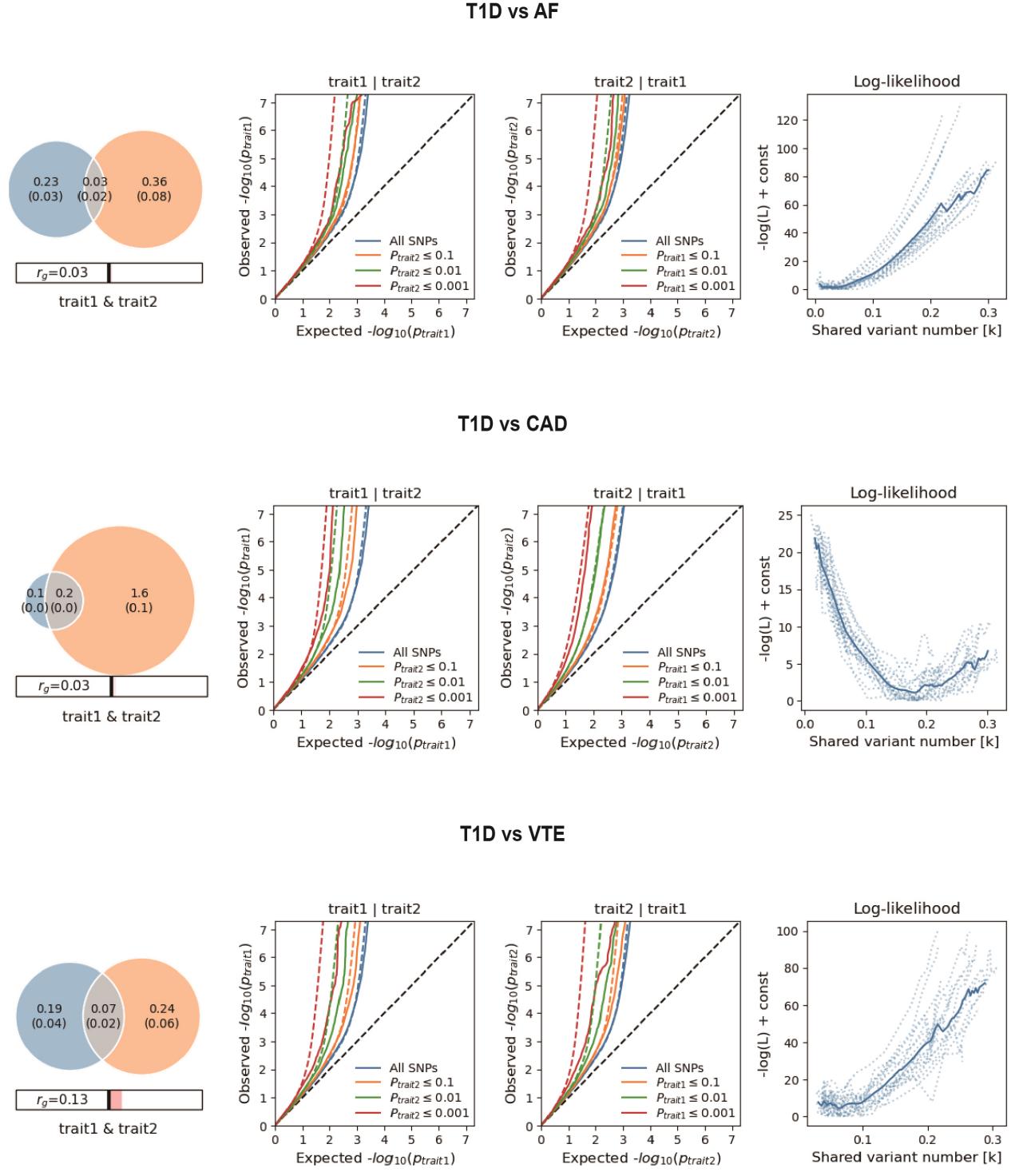

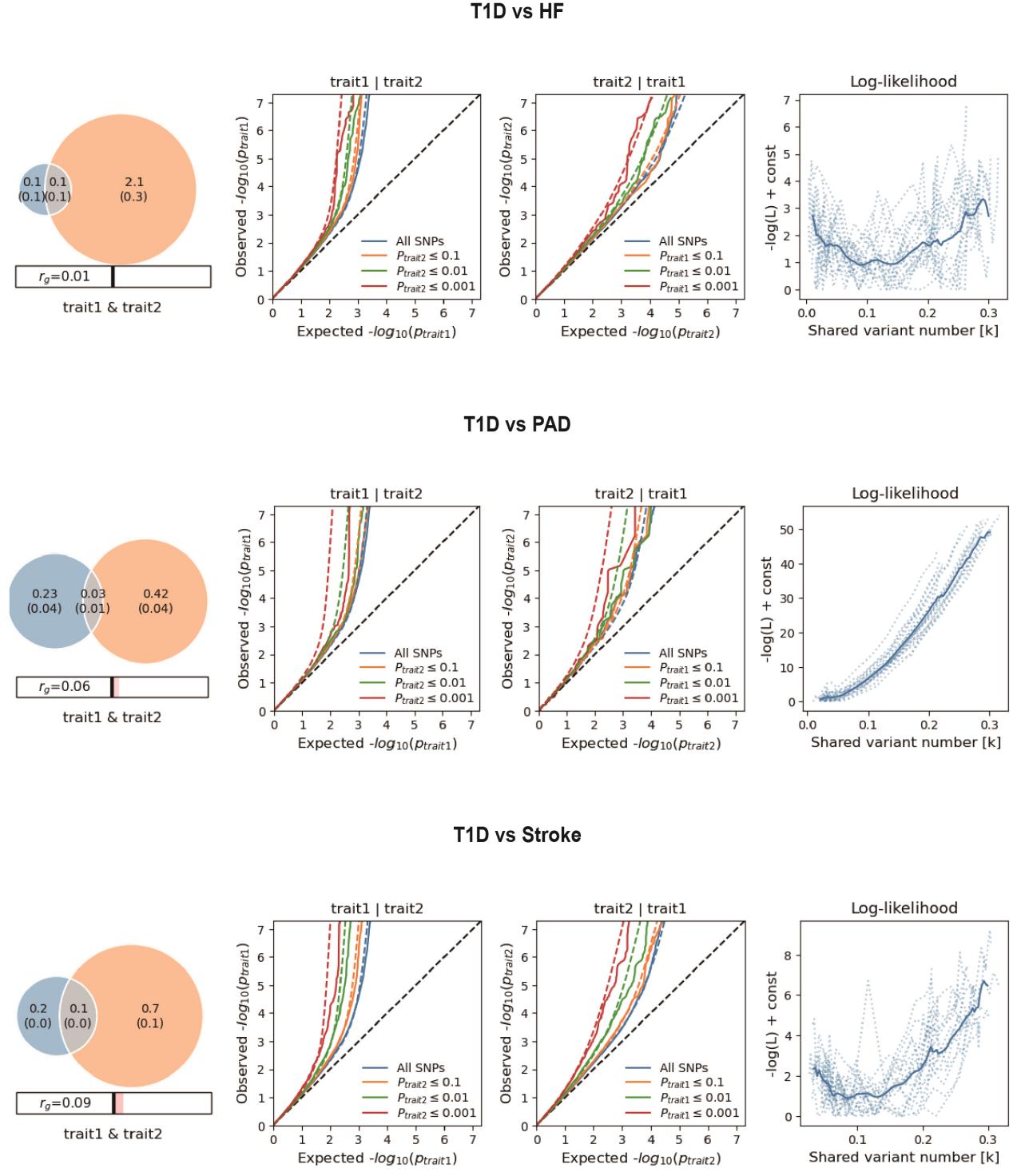

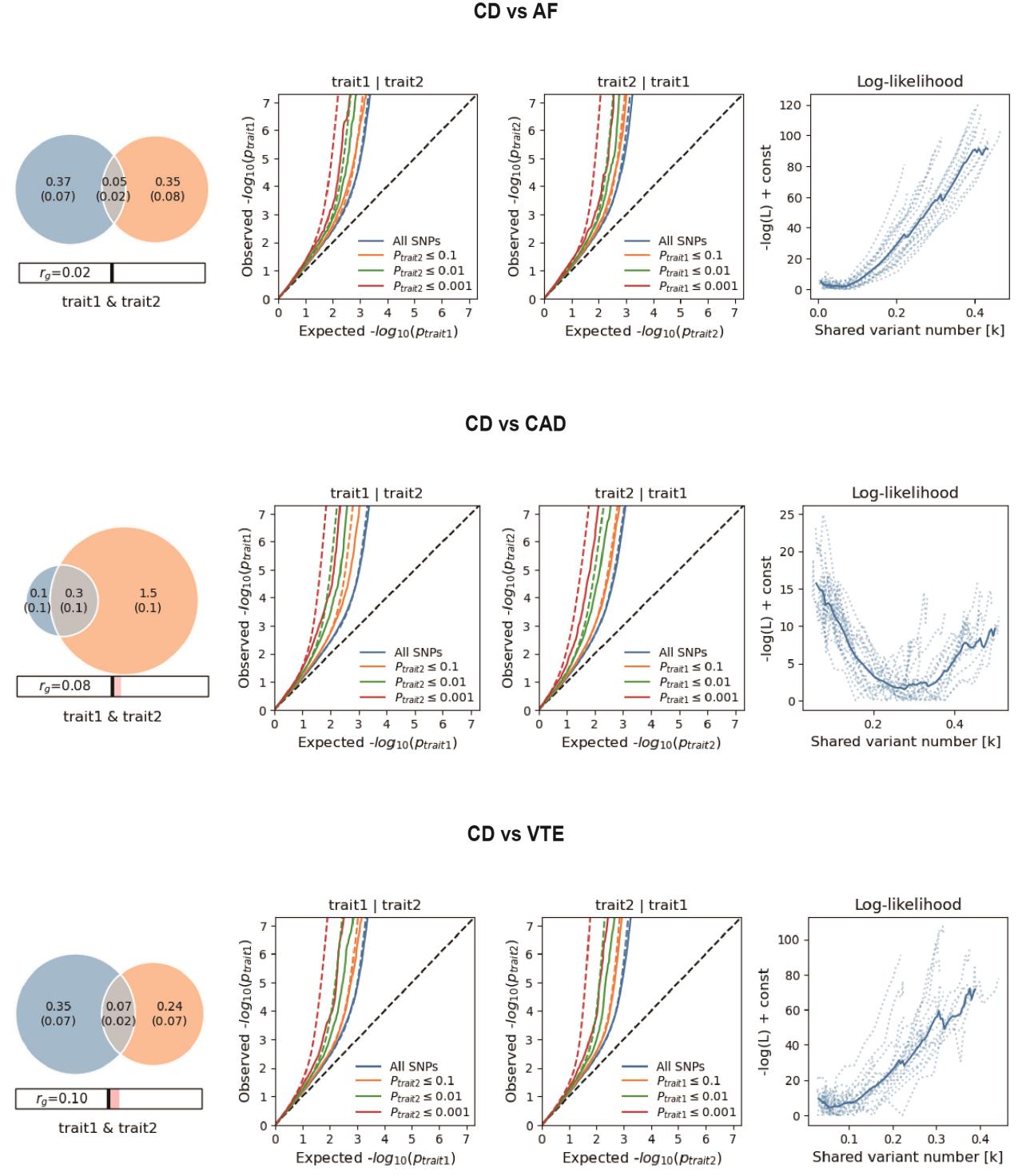

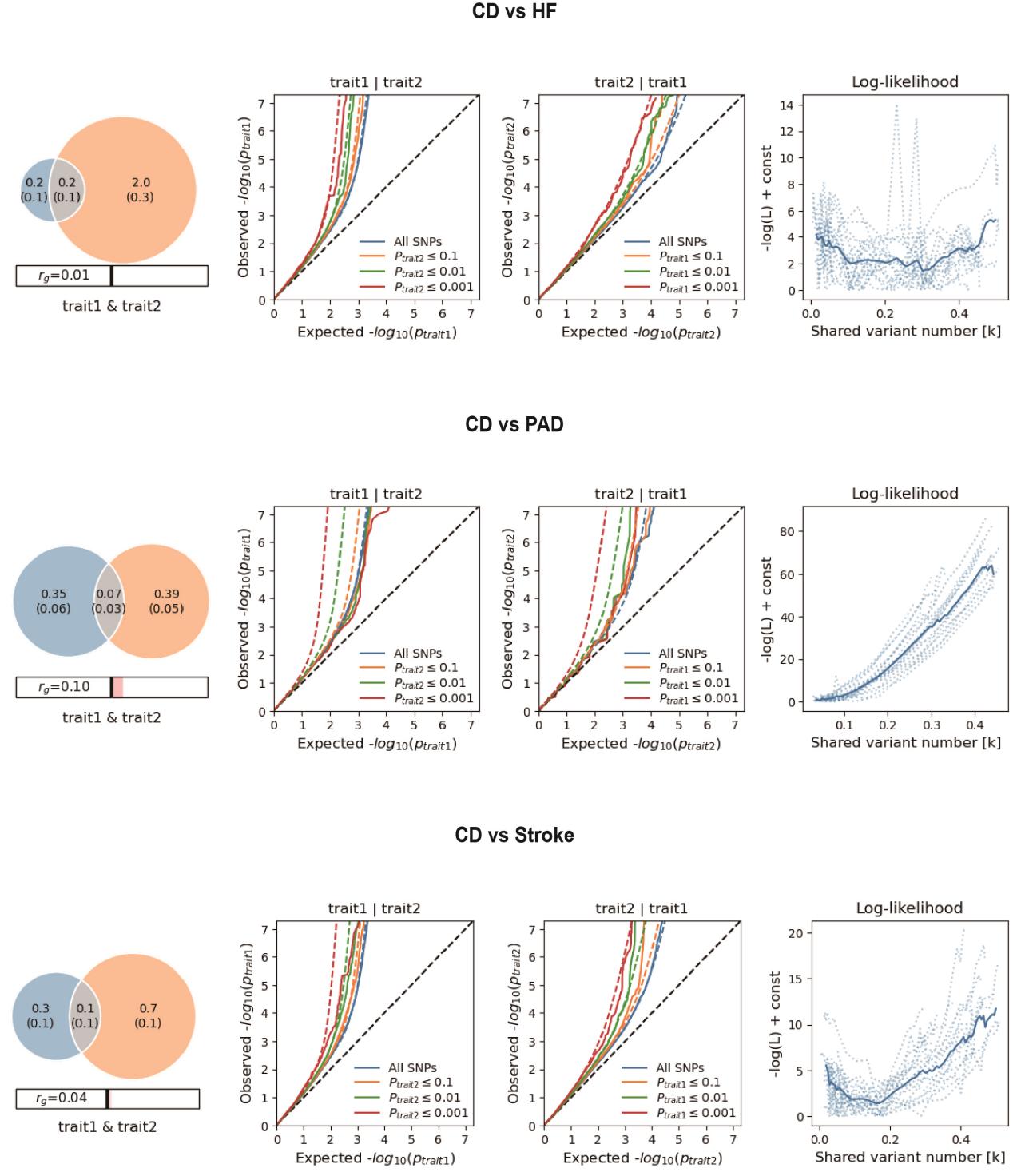

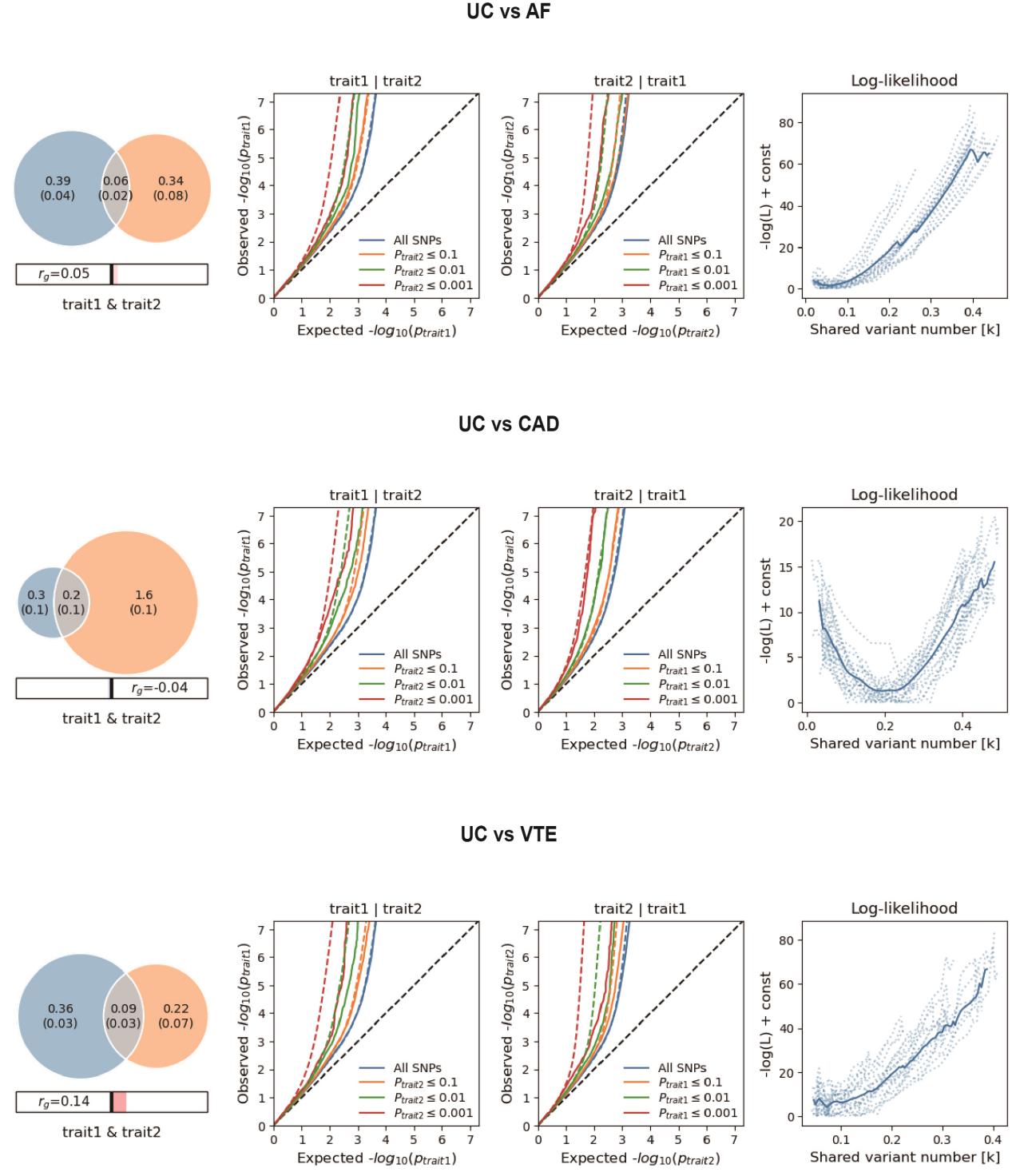

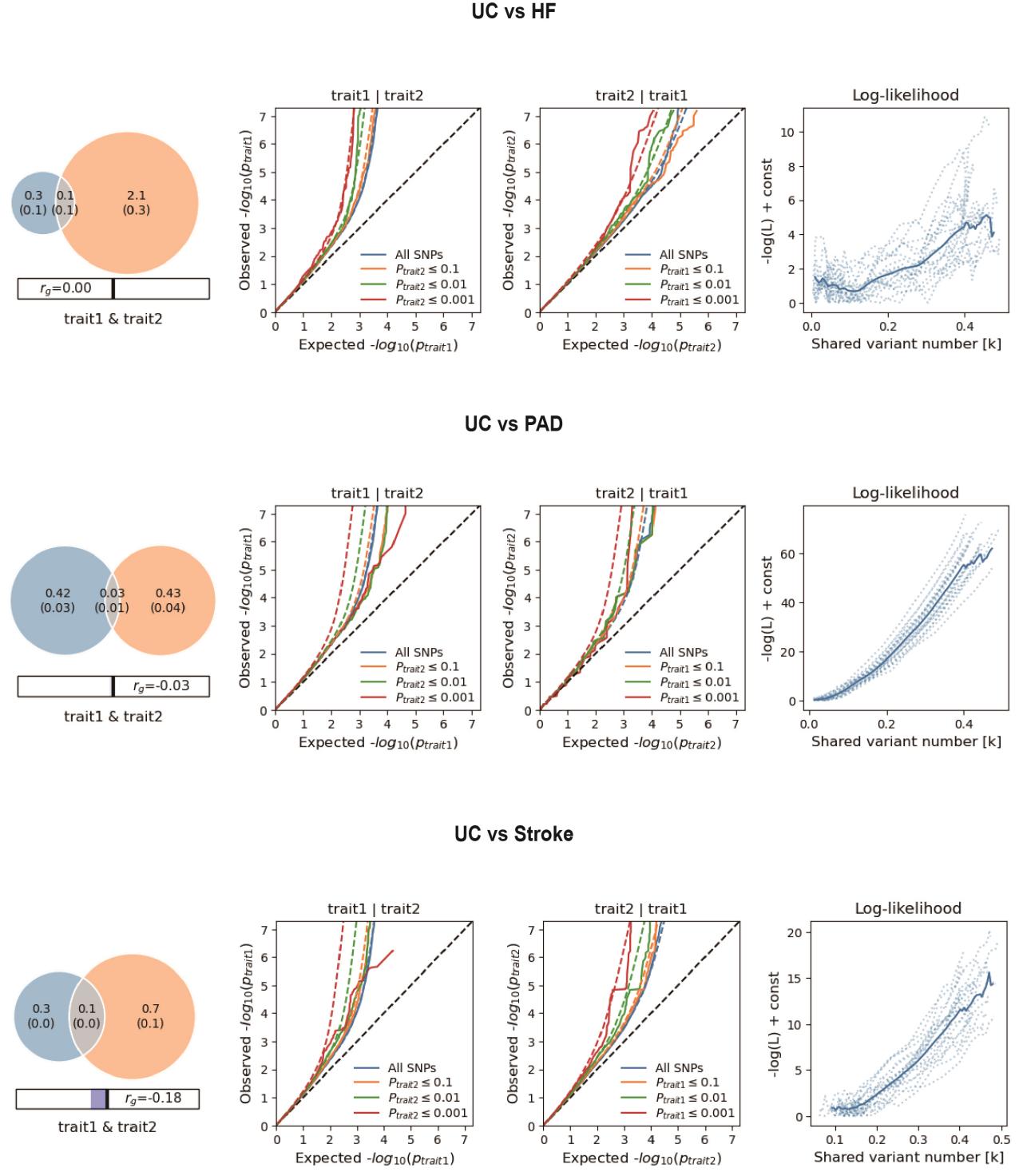

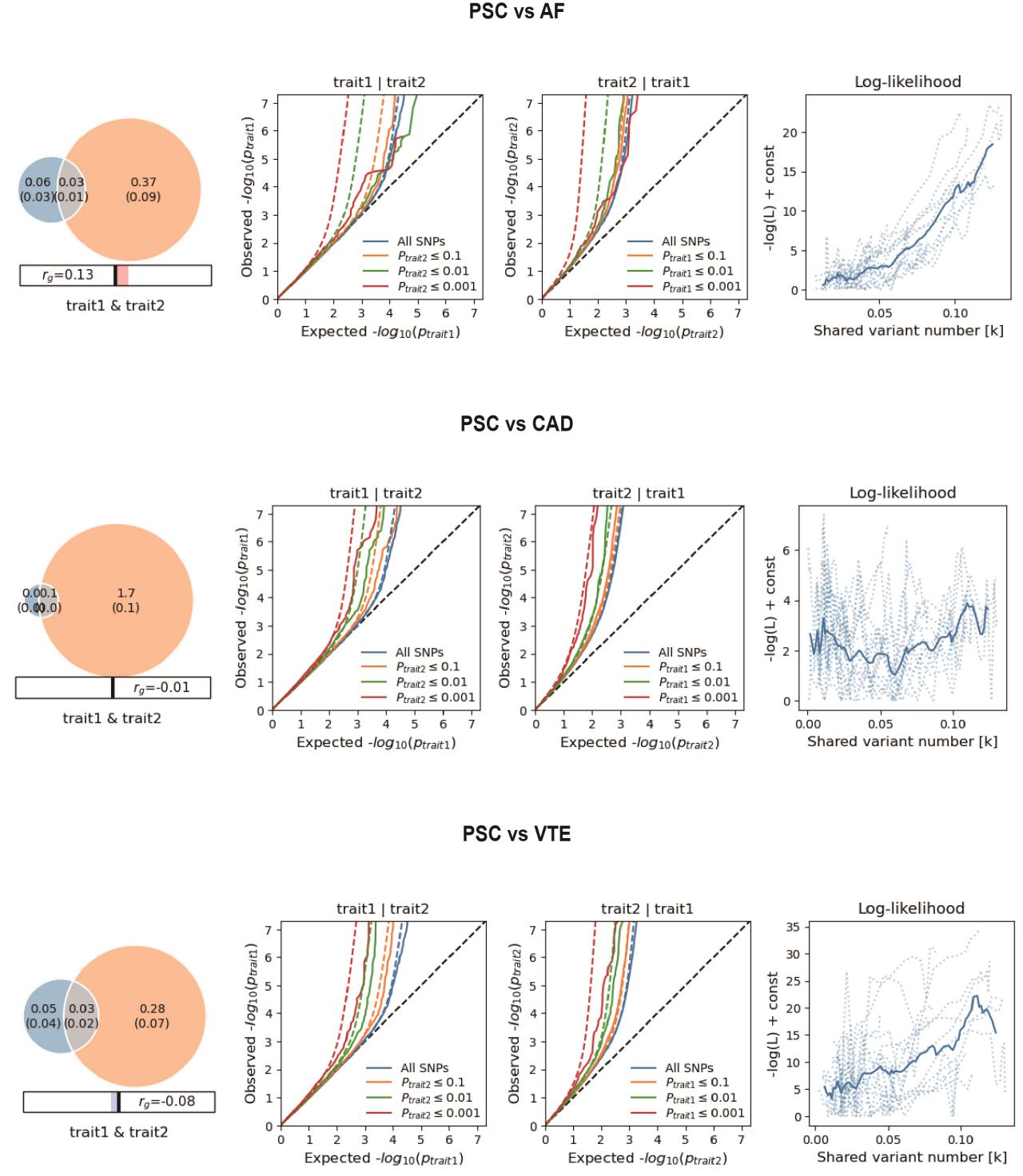

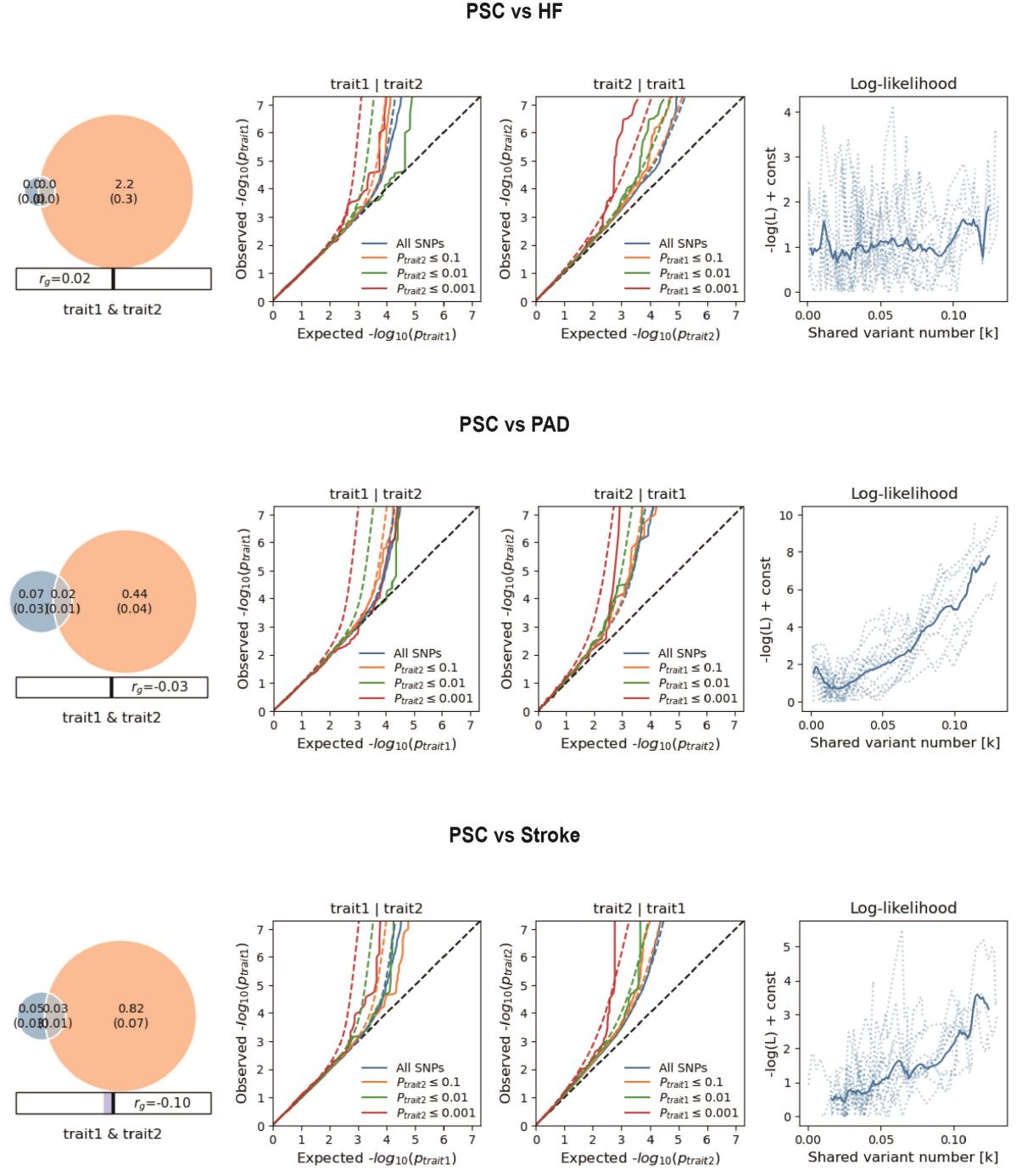

**Supplementary Fig. 1. Supplemental MiXeR figures for each of six autoimmune diseases and six cardiovascular diseases.**

On the left, MiXeR Venn diagrams illustrating MiXeR modelled genetic overlap, genome-wide genetic correlation (*r_g_*) and genetic correlation of shared variants (*r_g_s*) between ADs and CVDs. Conditional QQ plots of observed versus expected -log_10_ p-values in the primary trait as a function of the significance of the association with the secondary trait at the level of all SNPs (blue lines), p ≤ 0.1 (orange lines), p ≤ 0.01 (green lines) and p ≤ 0.001 (red lines). Dotted lines indicate model predictions for each stratum. The black dotted line is the expected Q-Q plot under the null hypothesis (no SNPs associated with the phenotype). Points on the Q-Q plot are weighted according to LD structure, using n=64 iterations of random pruning at an LD threshold r2 = 0.1. On the right, log-likelihood curves highlight the goodness of model fit by plotting the negative log-likelihood function (lower values correspond to better model fit) against the π12 parameter (number of influencing variants shared between two traits). The remaining parameters of the model were constrained to their fitted values. The π12 range on the log-likelihood plots goes from the smallest possible value π12 = *r_g_**sqrt (π1u, π2u) that is still compatible with the estimated genetic correlation, up to the largest possible value π12 = min(π1u, π2u) that corresponds to the minimum total polygenicity among the two traits. The minimum point indicates the best-fitting model estimate of the number of influencing variants shared between two traits. Rheumatoid arthritis, RA; Systemic lupus erythematosus, SLE; Type 1 diabetes, T1D; Crohn's disease, CD; Ulcerative colitis, UC; Primary sclerosing cholangitis, PSC; AF, Atrial fibrillation; CAD, Coronary artery disease; VTE, Venous thromboembolism; HF, Heart failure; PAD, Peripheral artery disease.

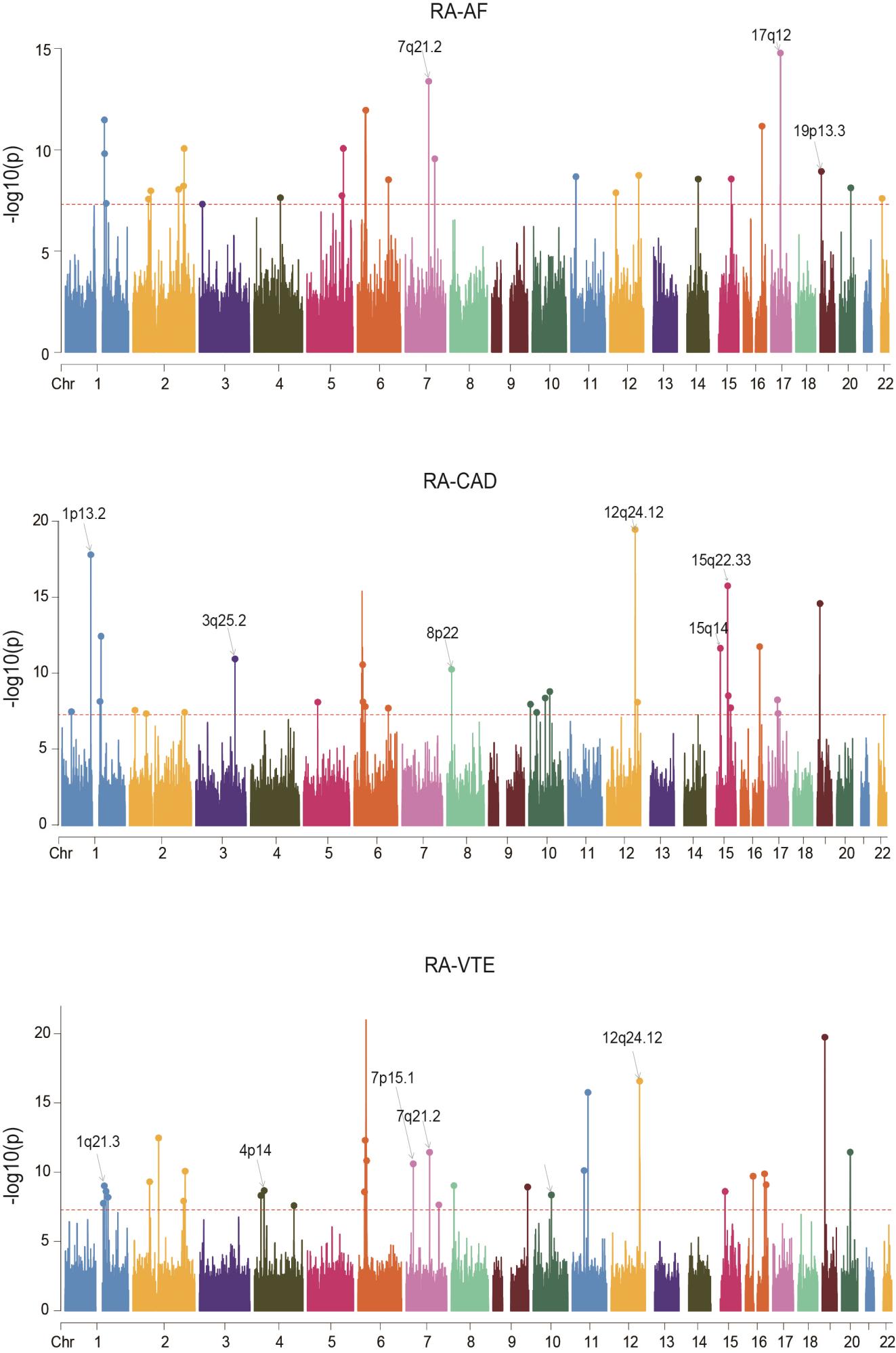

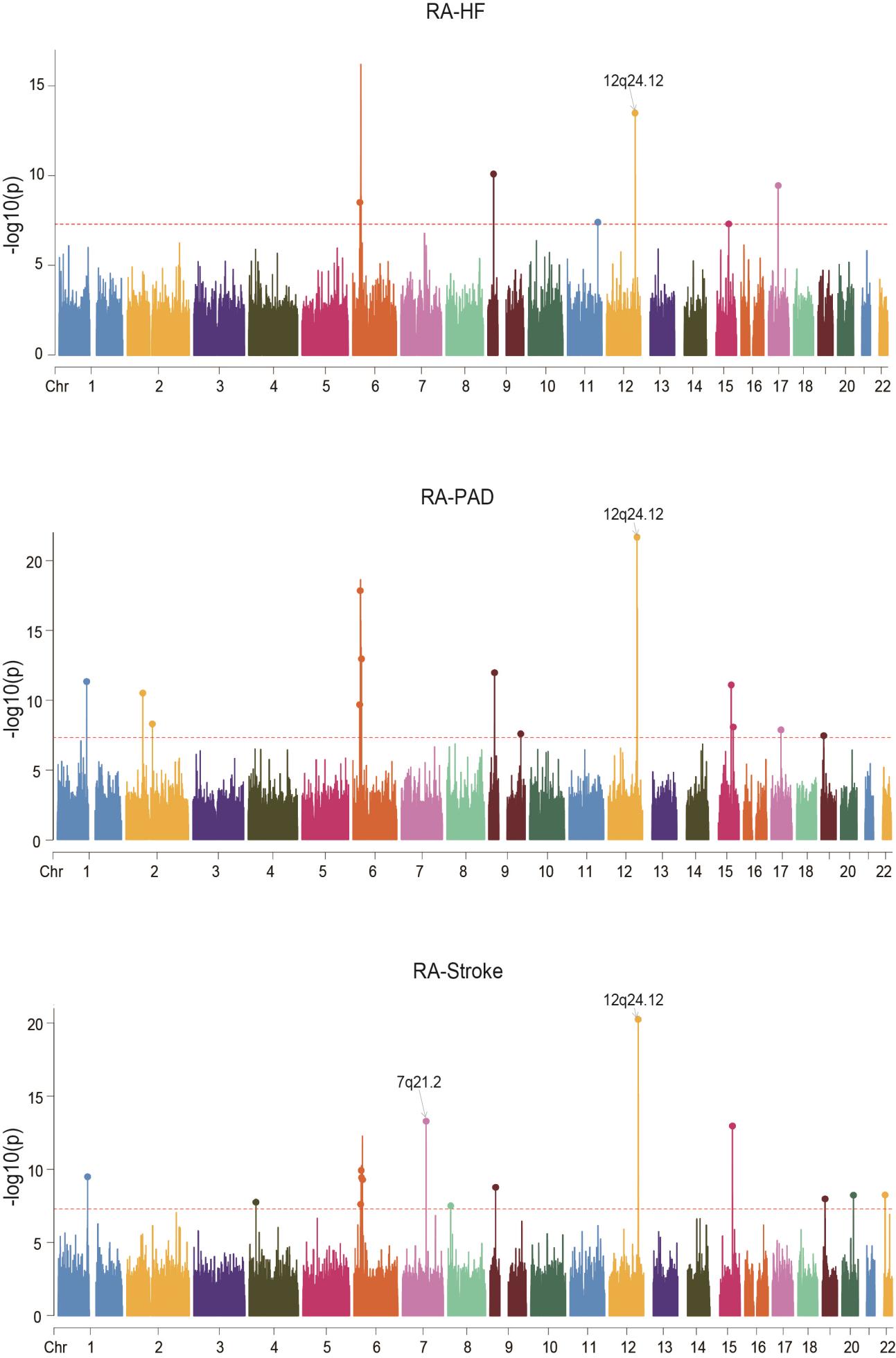

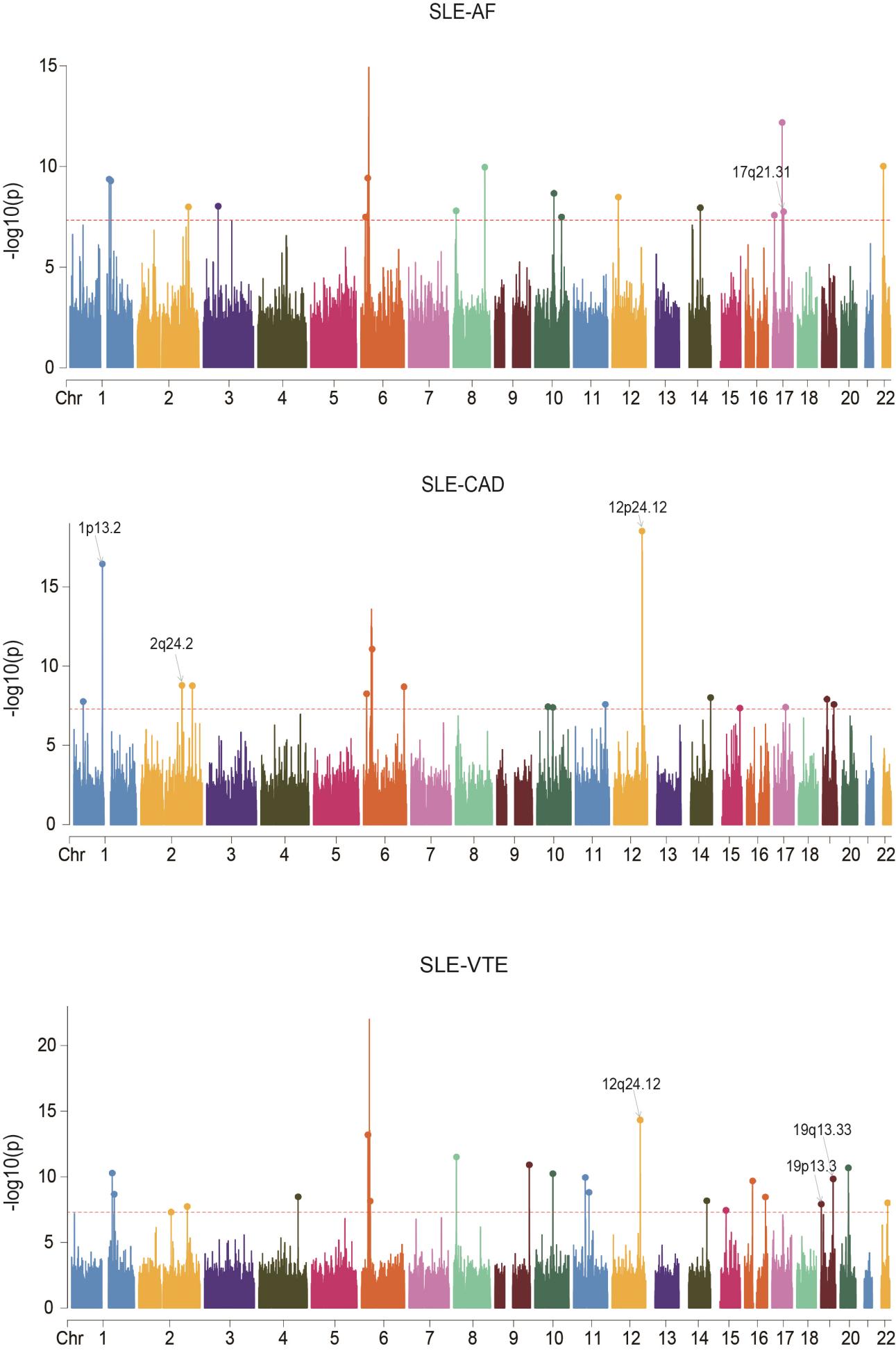

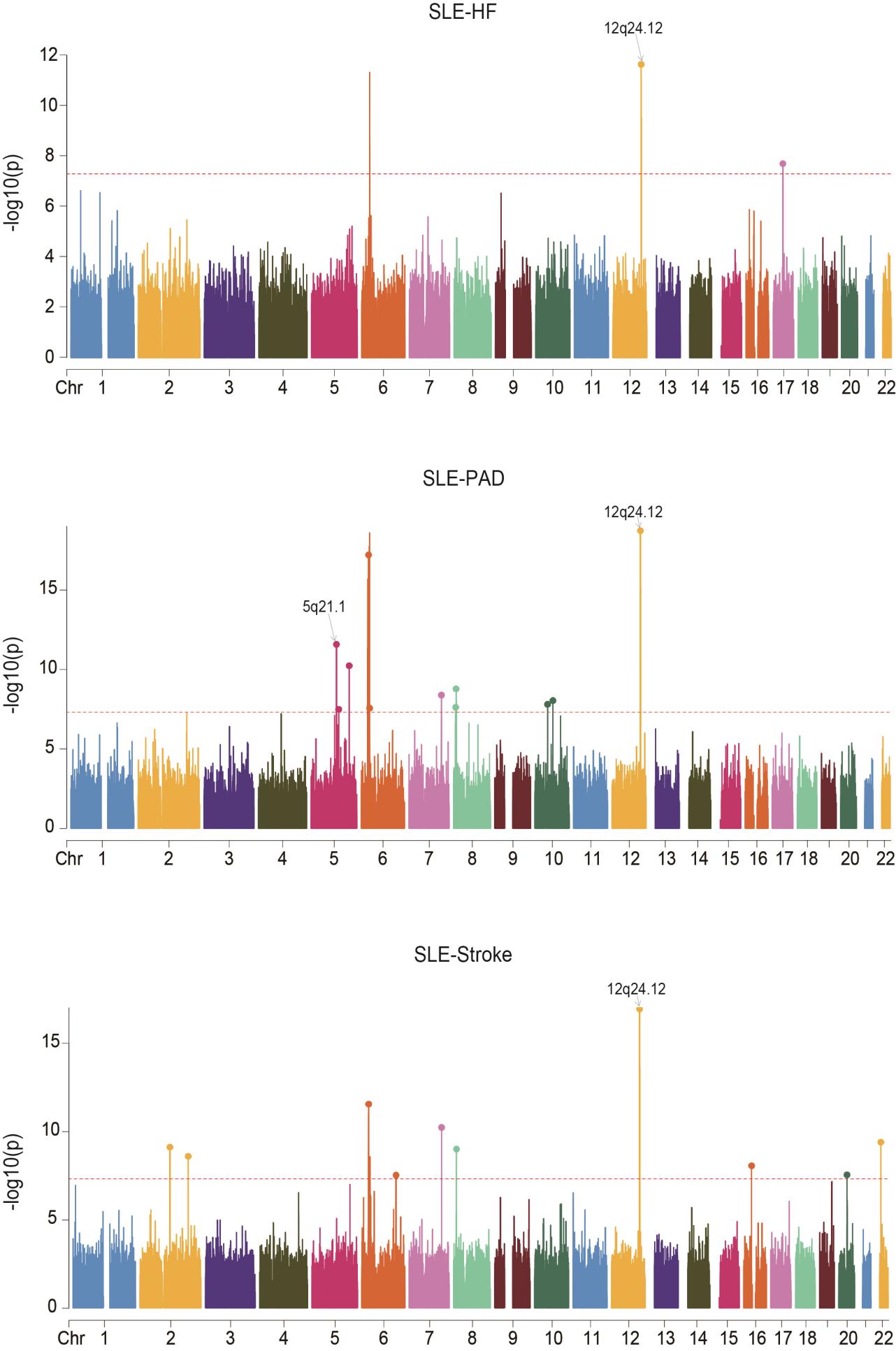

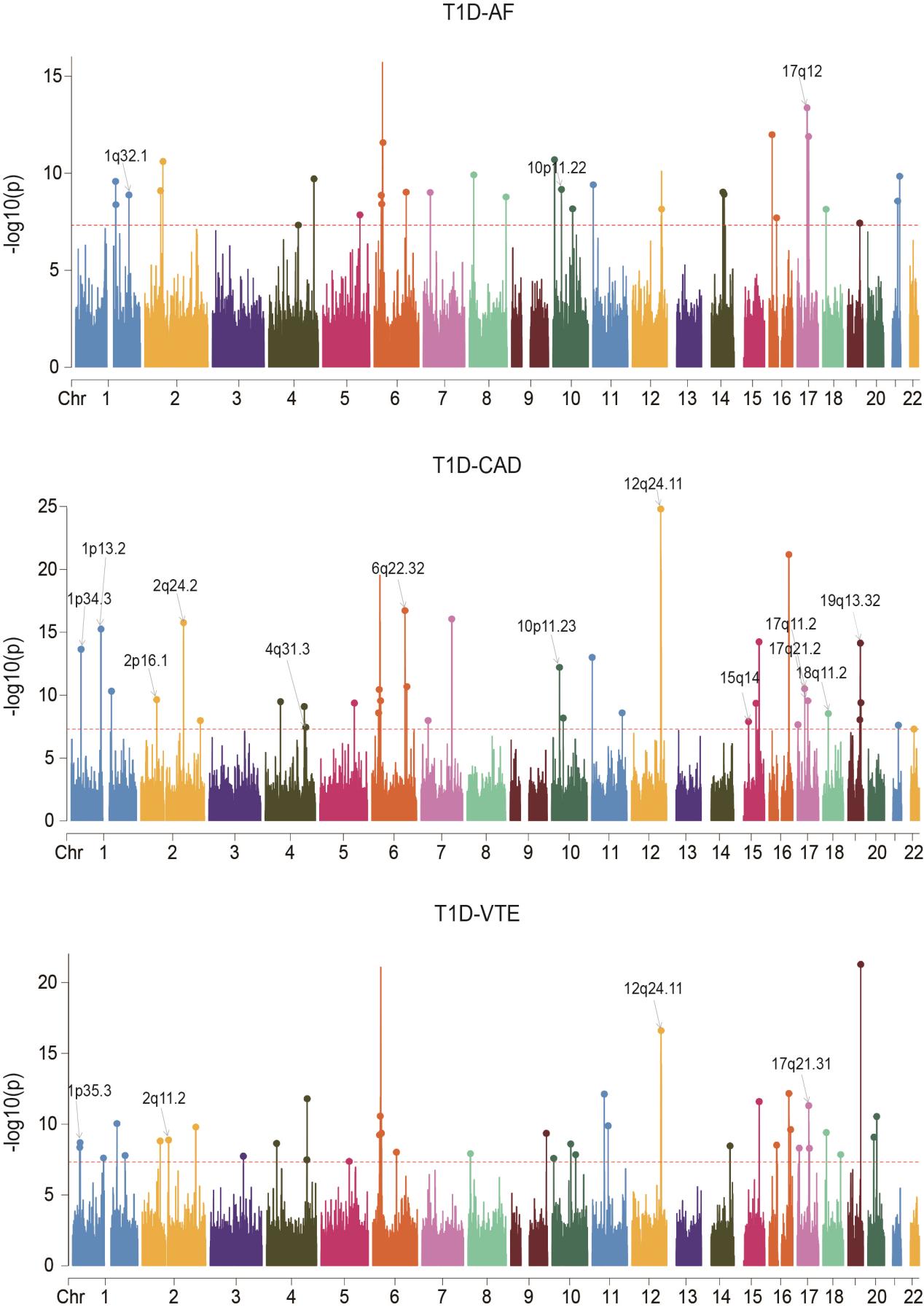

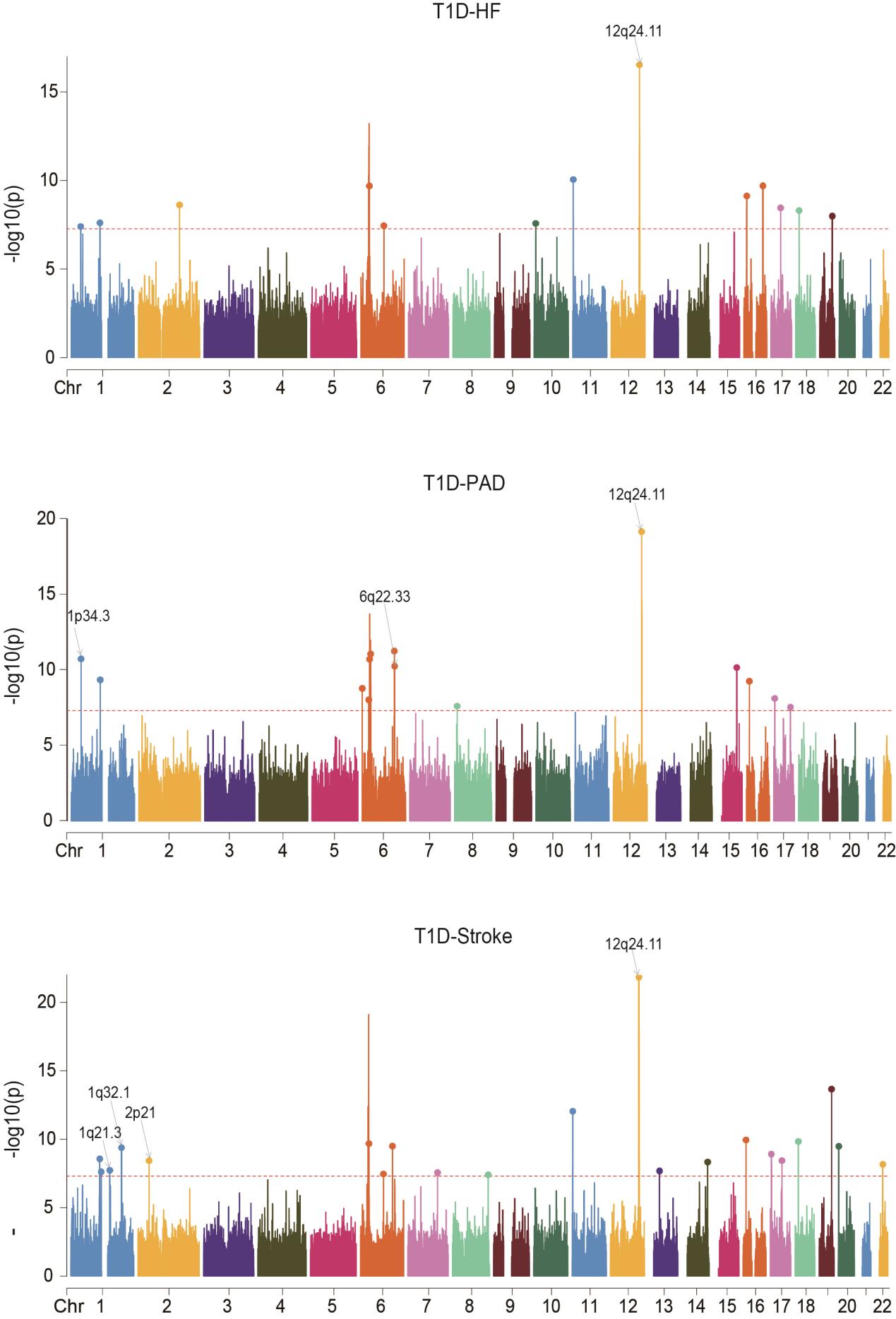

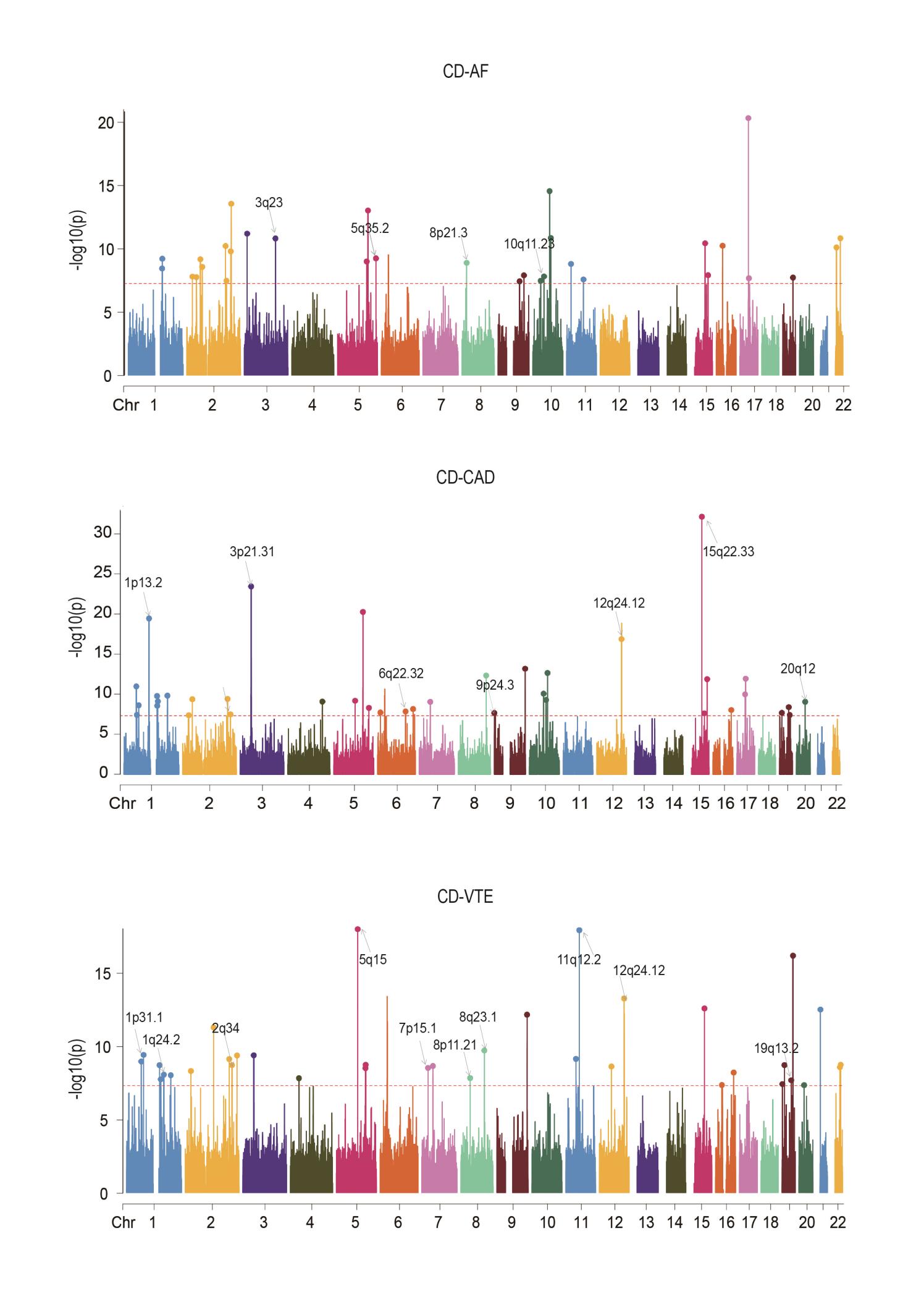

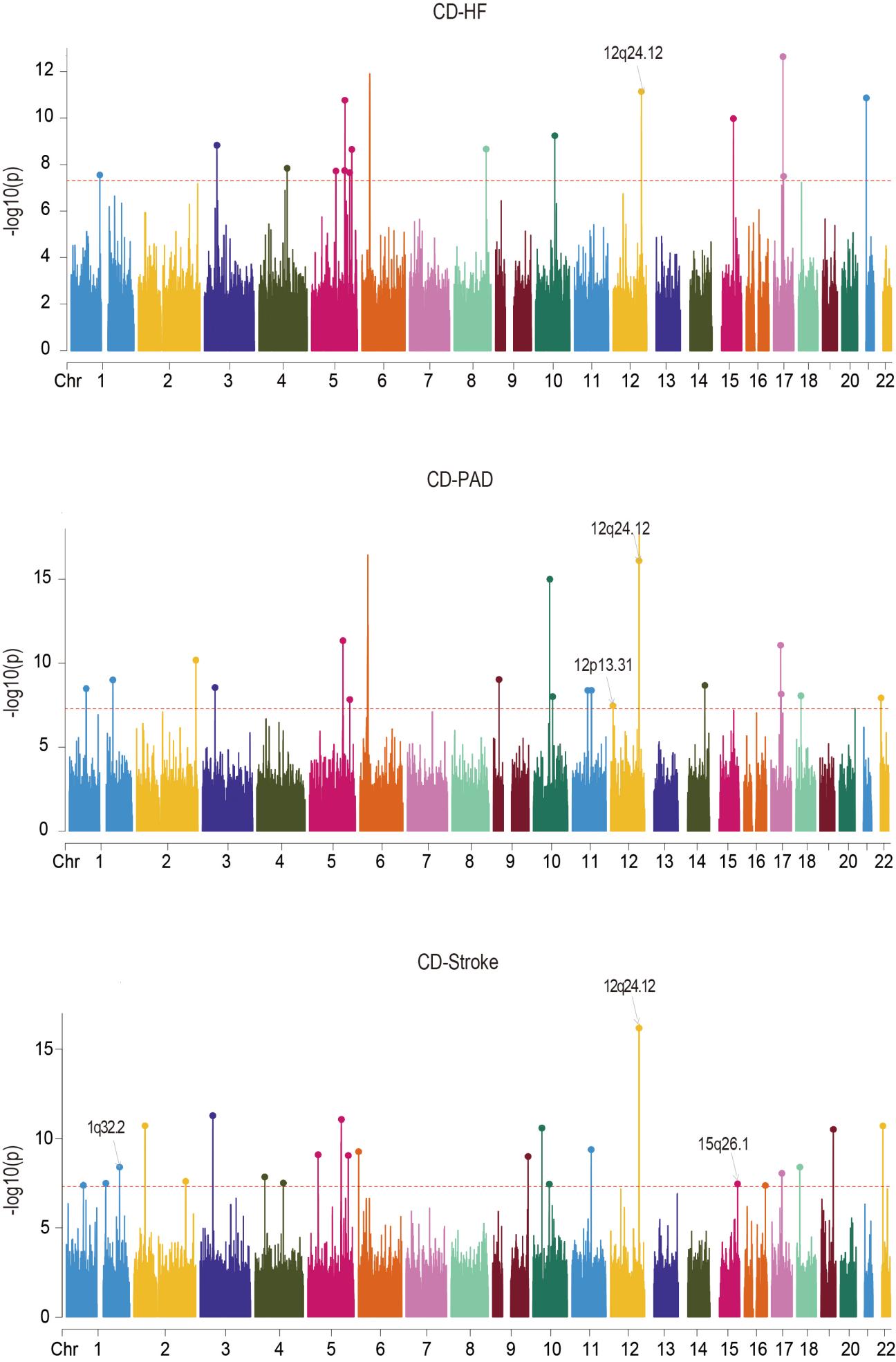

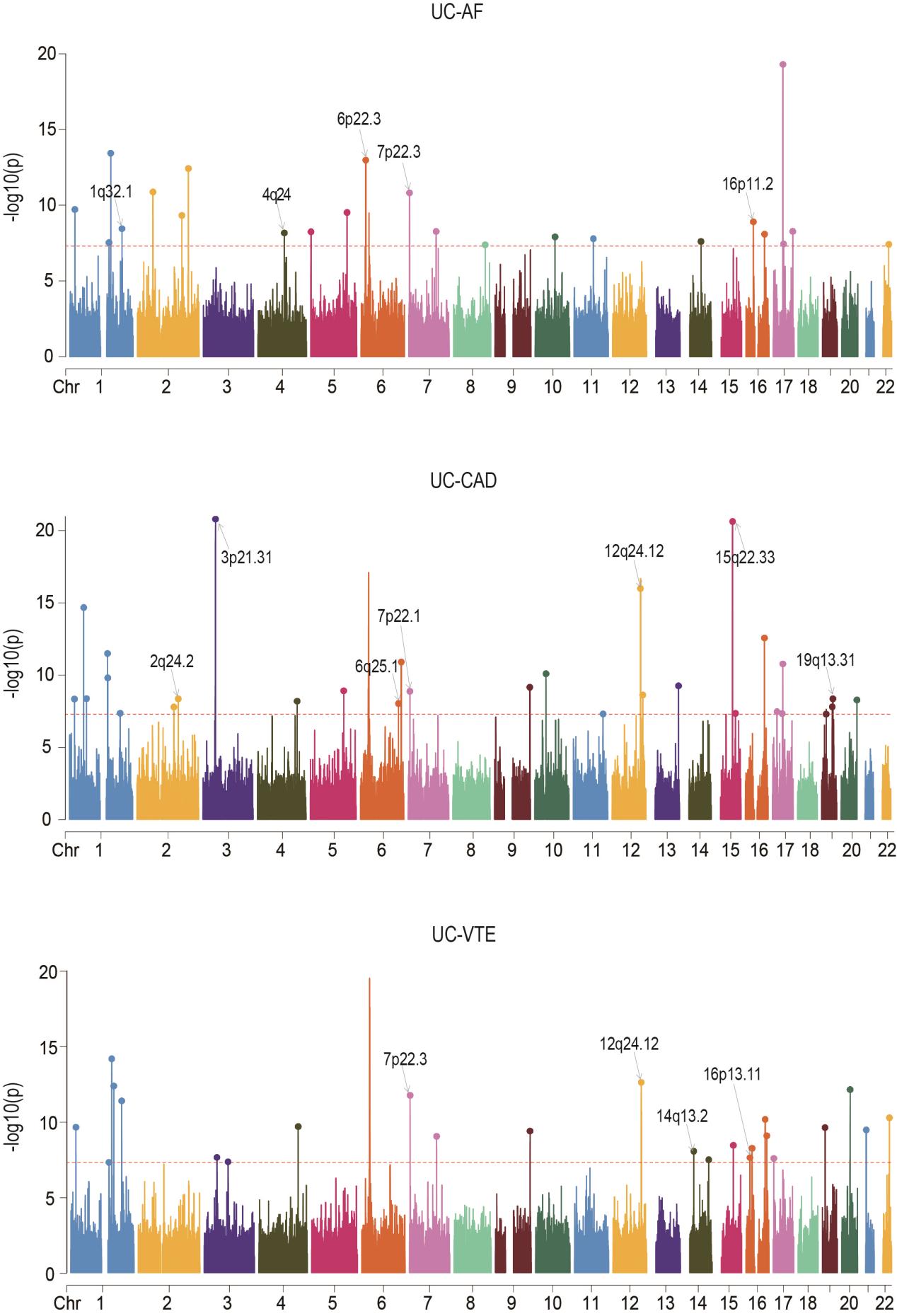

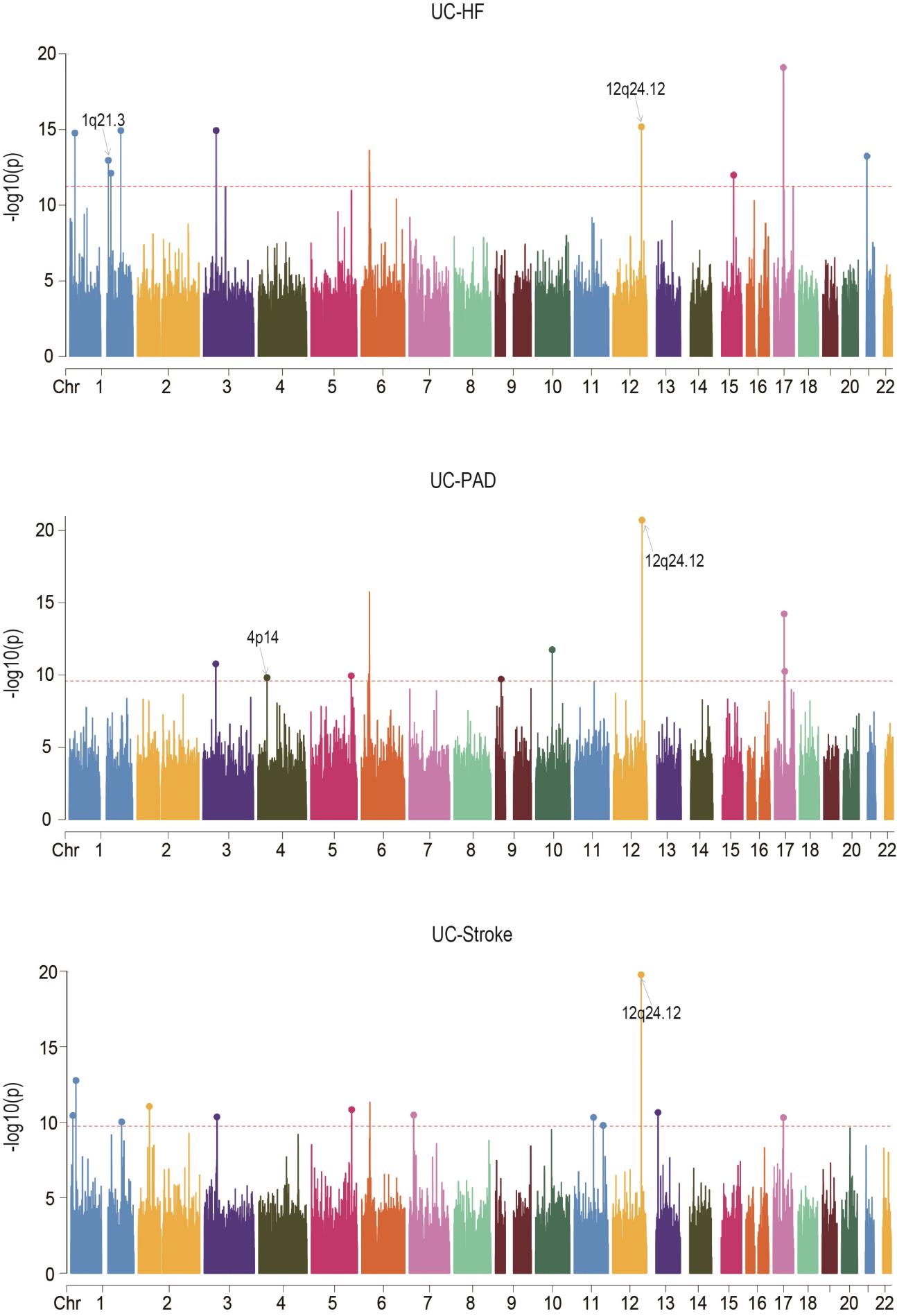

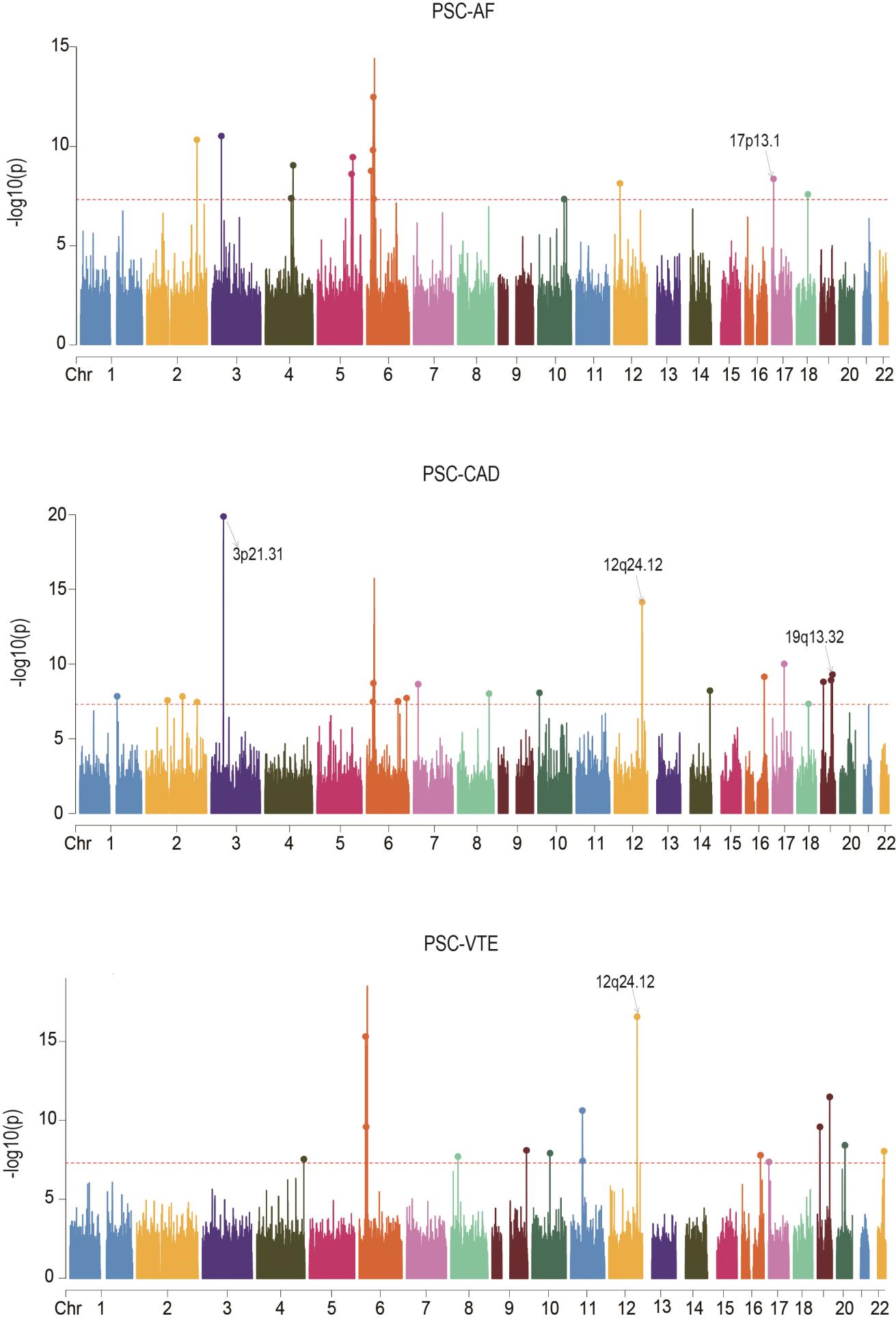

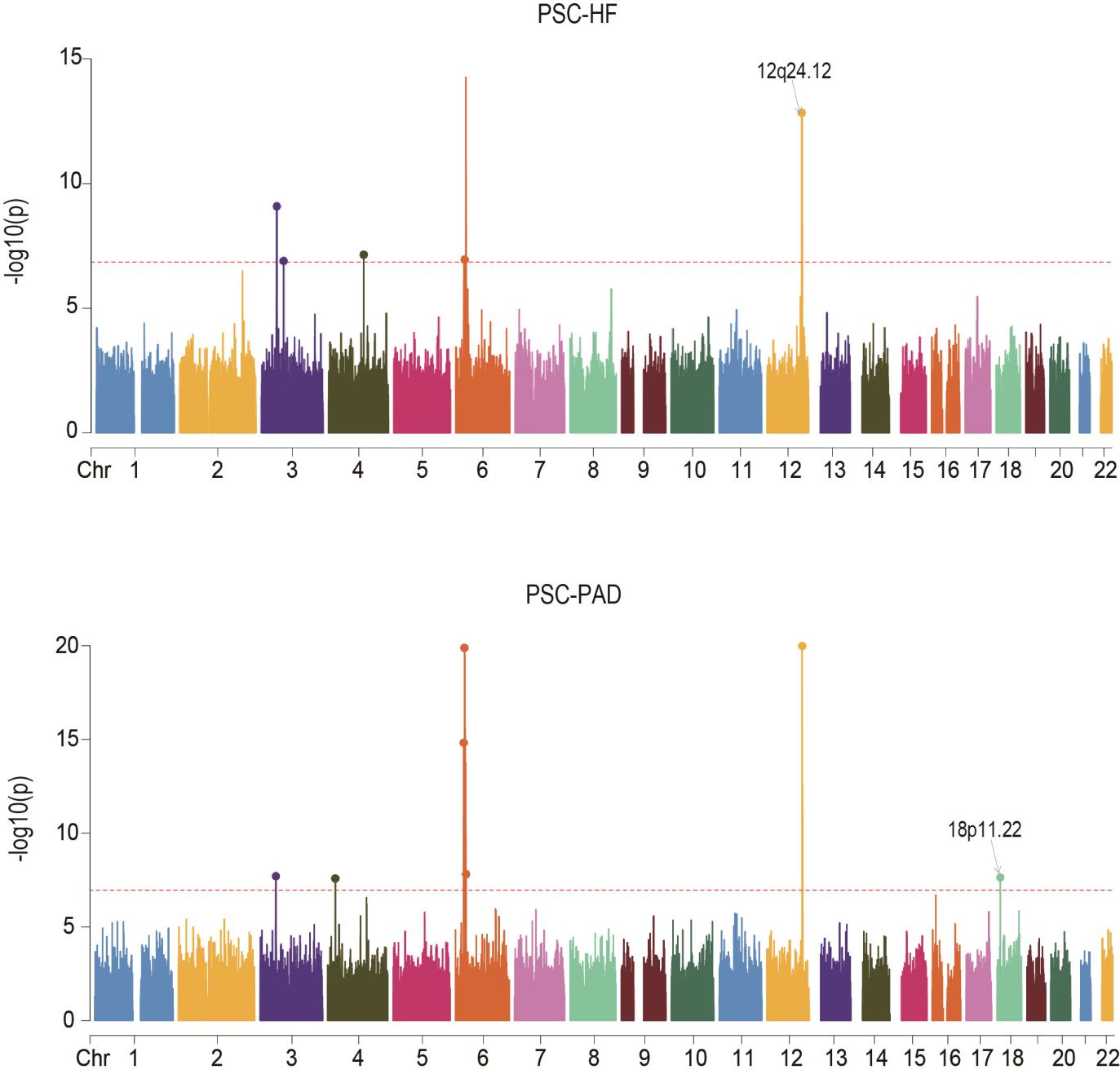

**Supplementary Fig. 2. Manhattan plots for the PLACO results of six autoimmune diseases and six cardiovascular diseases.**

The x-axis reflects the chromosomal position, and the y-axis reflects negative log_10_ transformed *P*-values for each SNP. The horizontal dashed red line indicates the genome-wide significant *P*-value of -log_10_ (5×10^-8^). The independent genome-wide significant associations with the smallest P-value (Top lead SNP) are encircled in a colorful circle. Only SNPs shared across all summary statistics were included. Labels are the chromosome regions where genomic risk loci with strong evidence for colocalization (PP.H_4_ > 0.7) are located. Rheumatoid arthritis, RA; Systemic lupus erythematosus, SLE; Type 1 diabetes, T1D; Crohn's disease, CD; Ulcerative colitis, UC; Primary sclerosing cholangitis, PSC; AF, Atrial fibrillation; CAD, Coronary artery disease; VTE, Venous thromboembolism; HF, Heart failure; PAD, Peripheral artery disease.

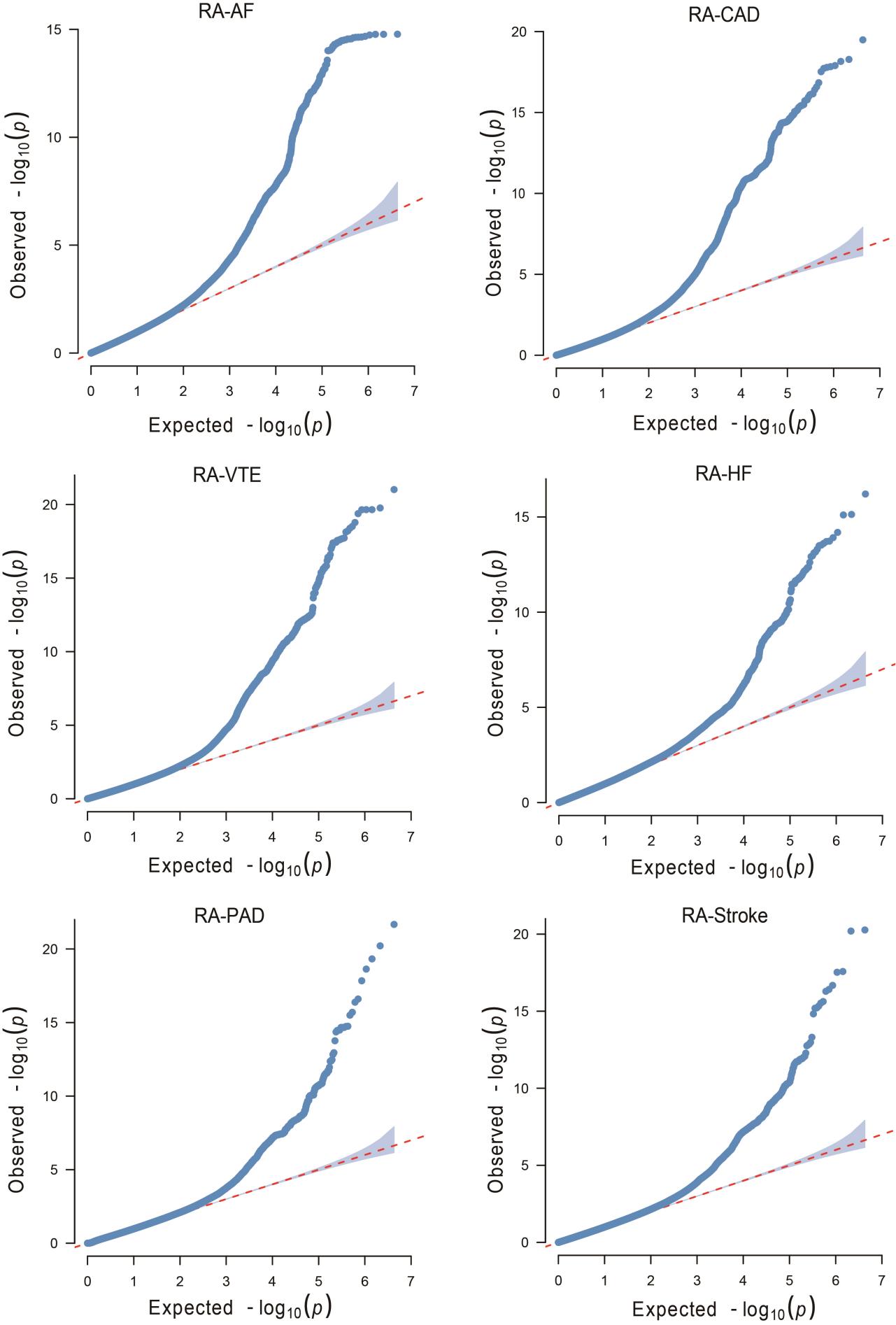

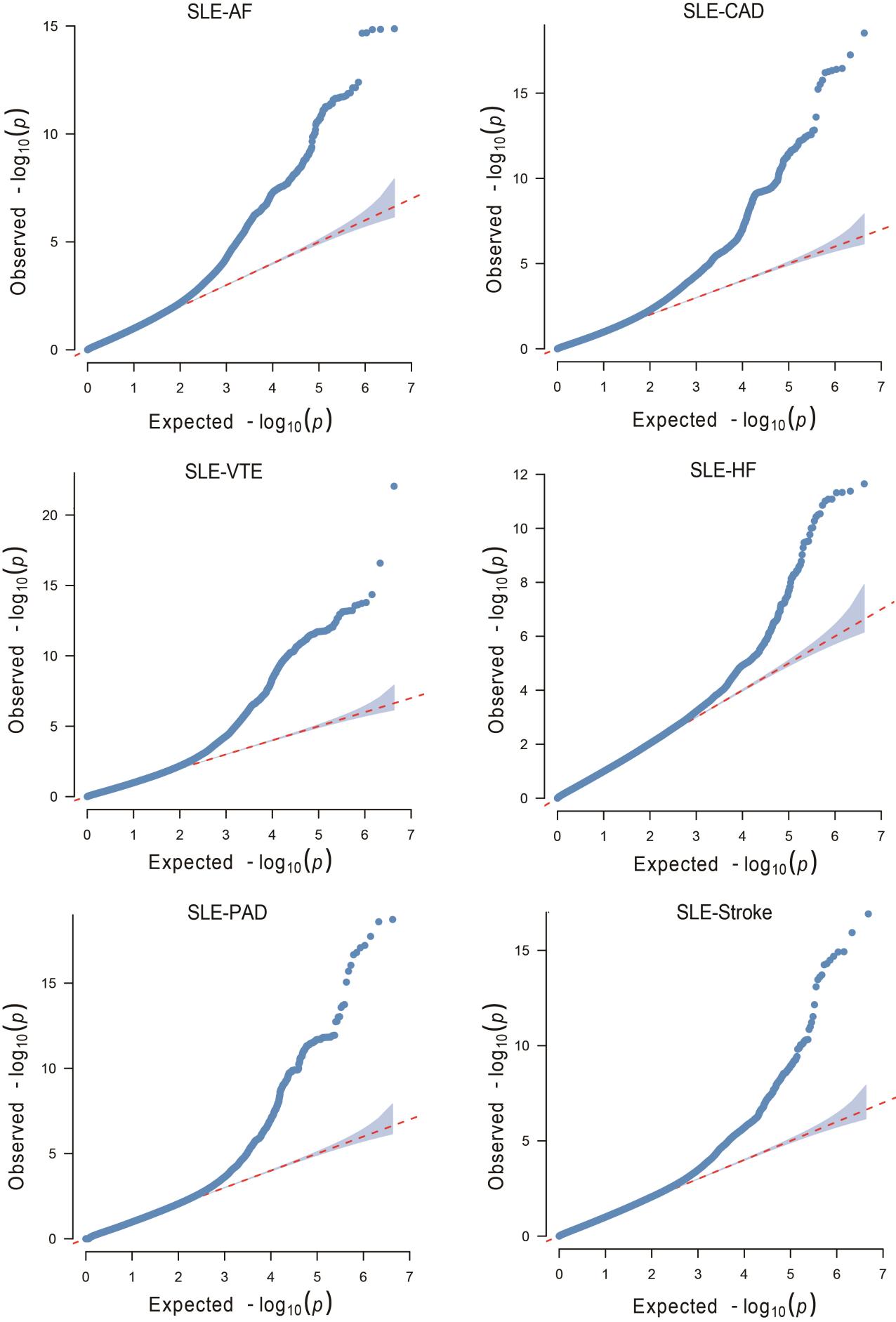

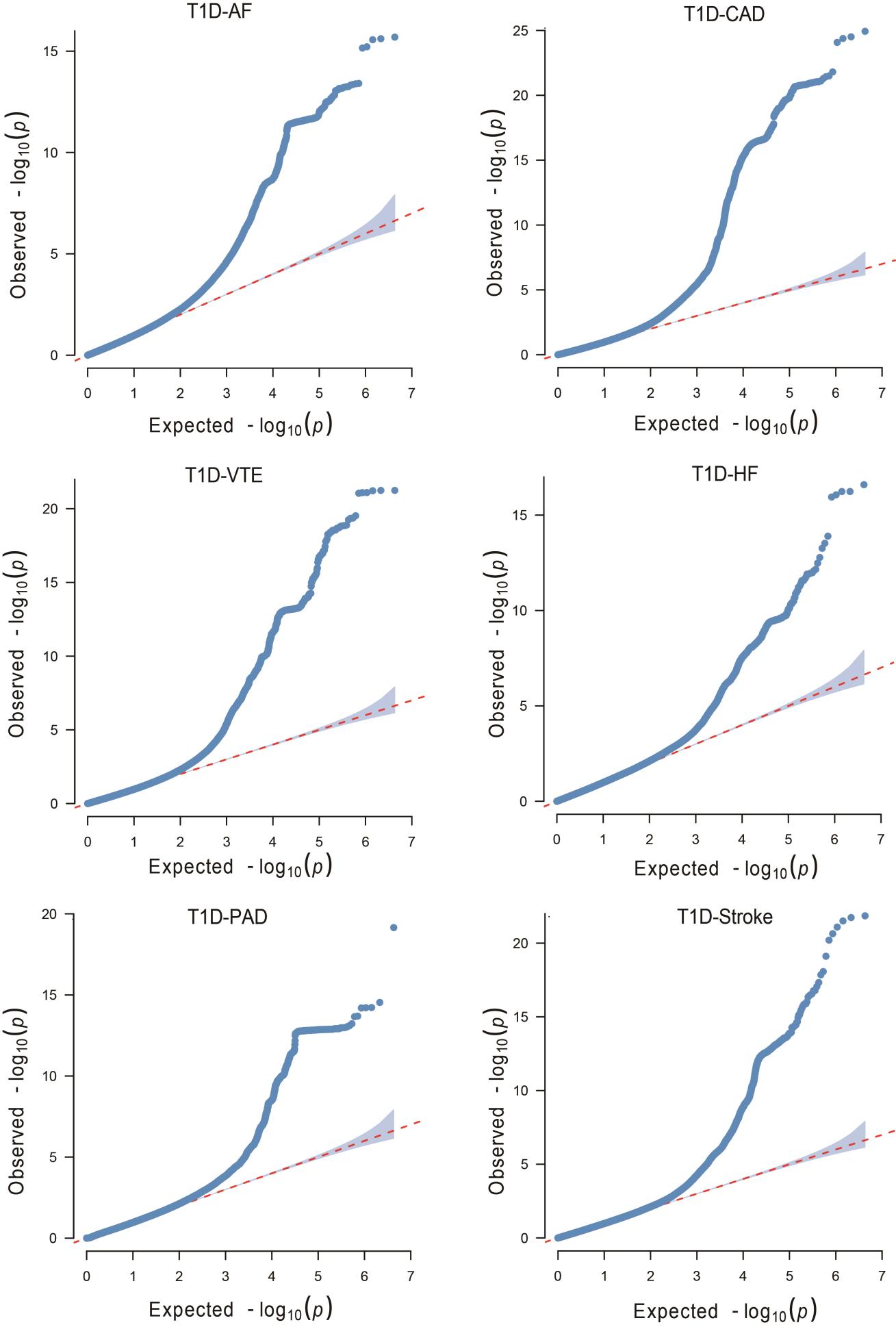

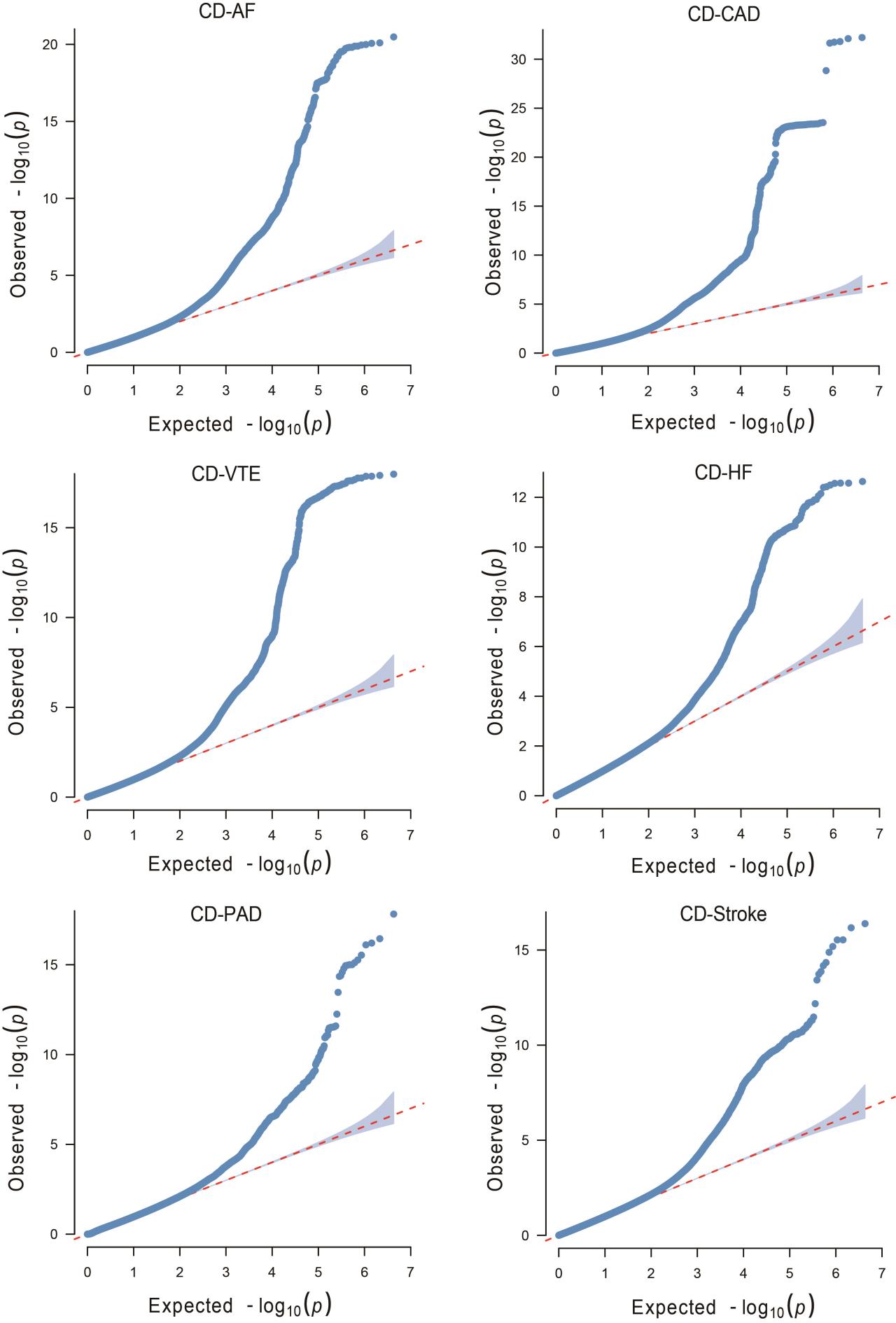

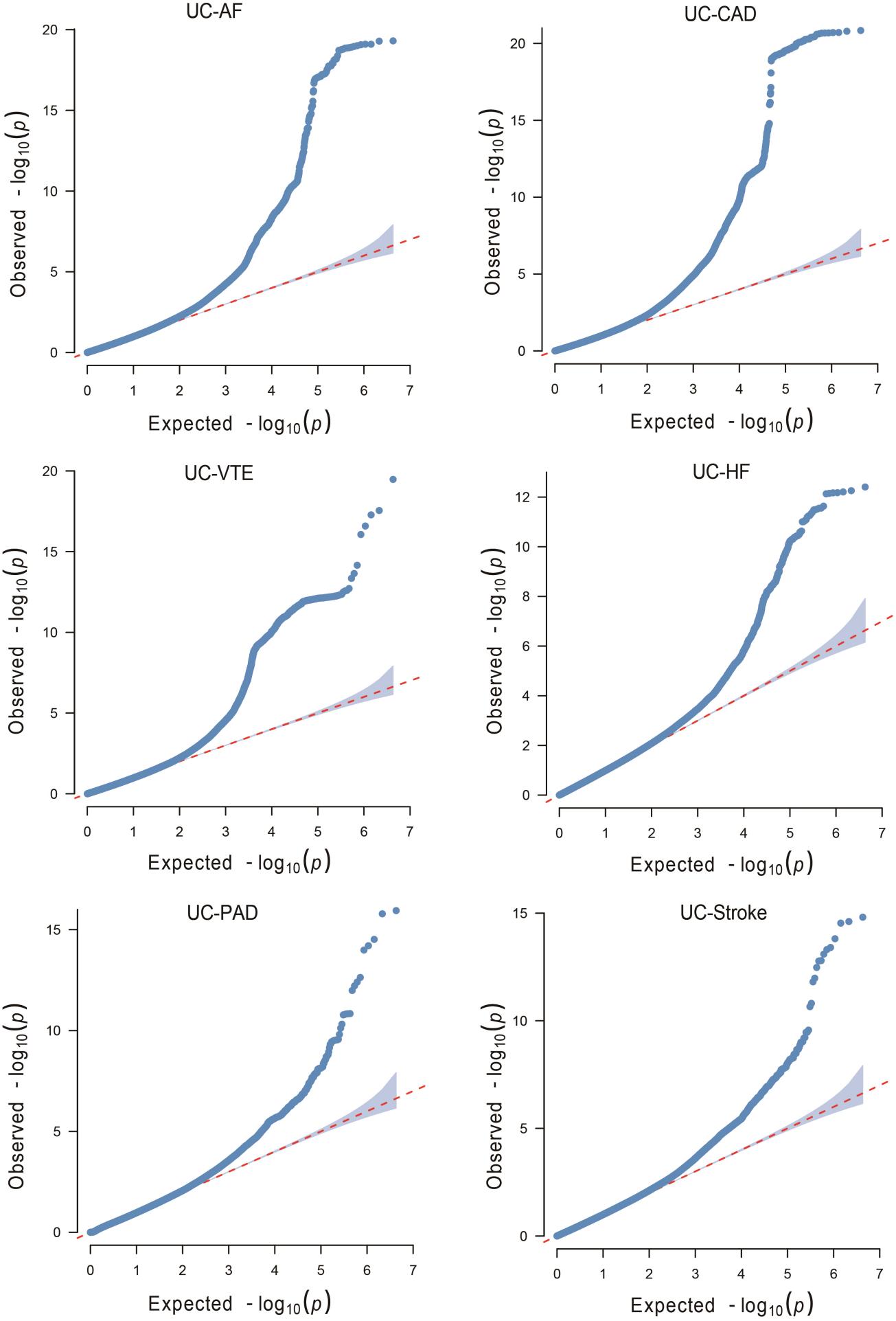

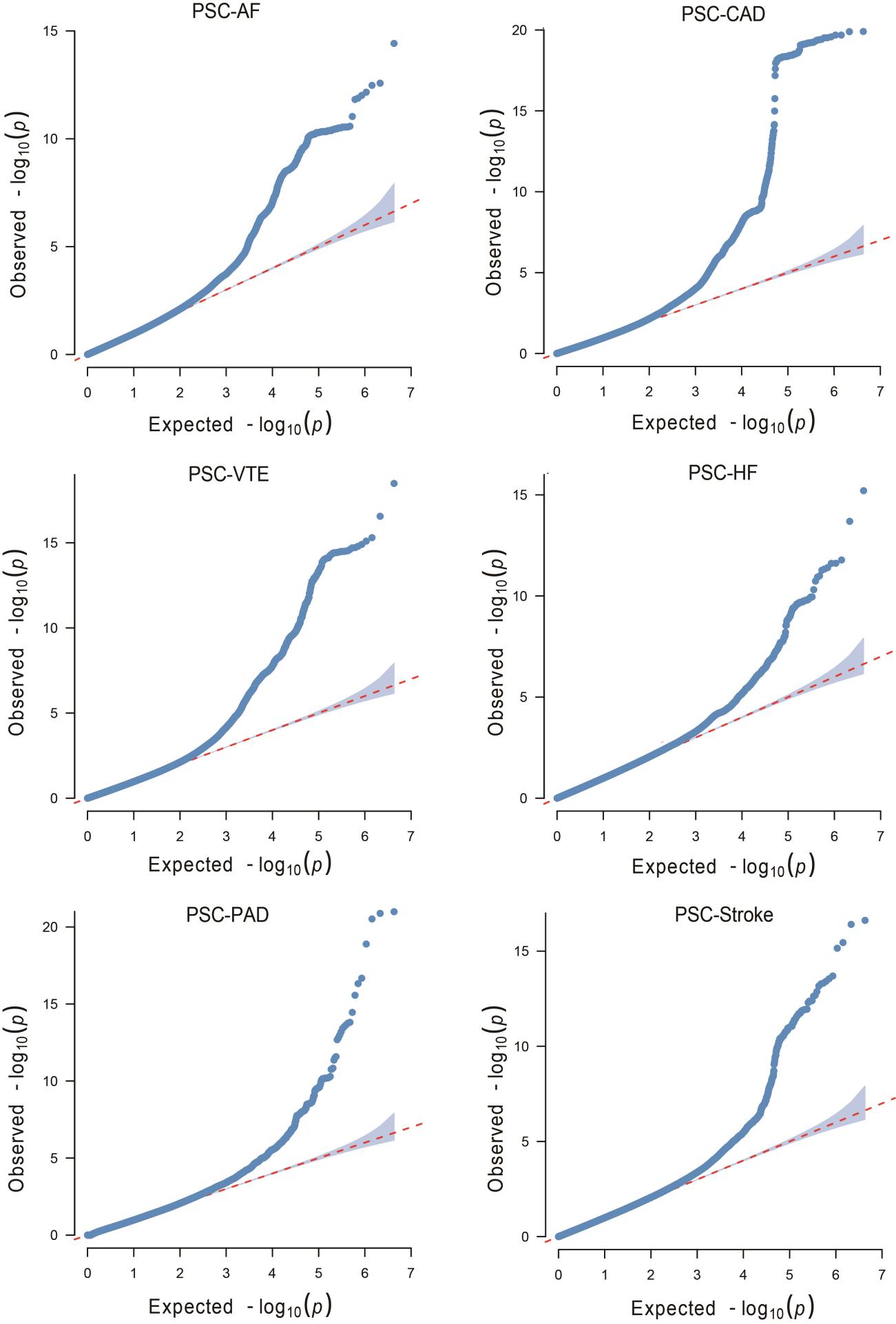

**Supplementary Fig. 3. Quantile-quantile (Q-Q) plots for PLACO results of six autoimmune diseases and six cardiovascular diseases.**

QQ plots depict expected -log10 *P*-values (x-axis) against observed -log10 *P_PLACO_*-values (y-axis). Red dots indicate significant pleiotropic variants (*P_PLACO_* < 5×10^-8^). Rheumatoid arthritis, RA; Systemic lupus erythematosus, SLE; Type 1 diabetes, T1D; Crohn's disease, CD; Ulcerative colitis, UC; Primary sclerosing cholangitis, PSC; AF, Atrial fibrillation; CAD, Coronary artery disease; VTE, Venous thromboembolism; HF, Heart failure; PAD, Peripheral artery disease.

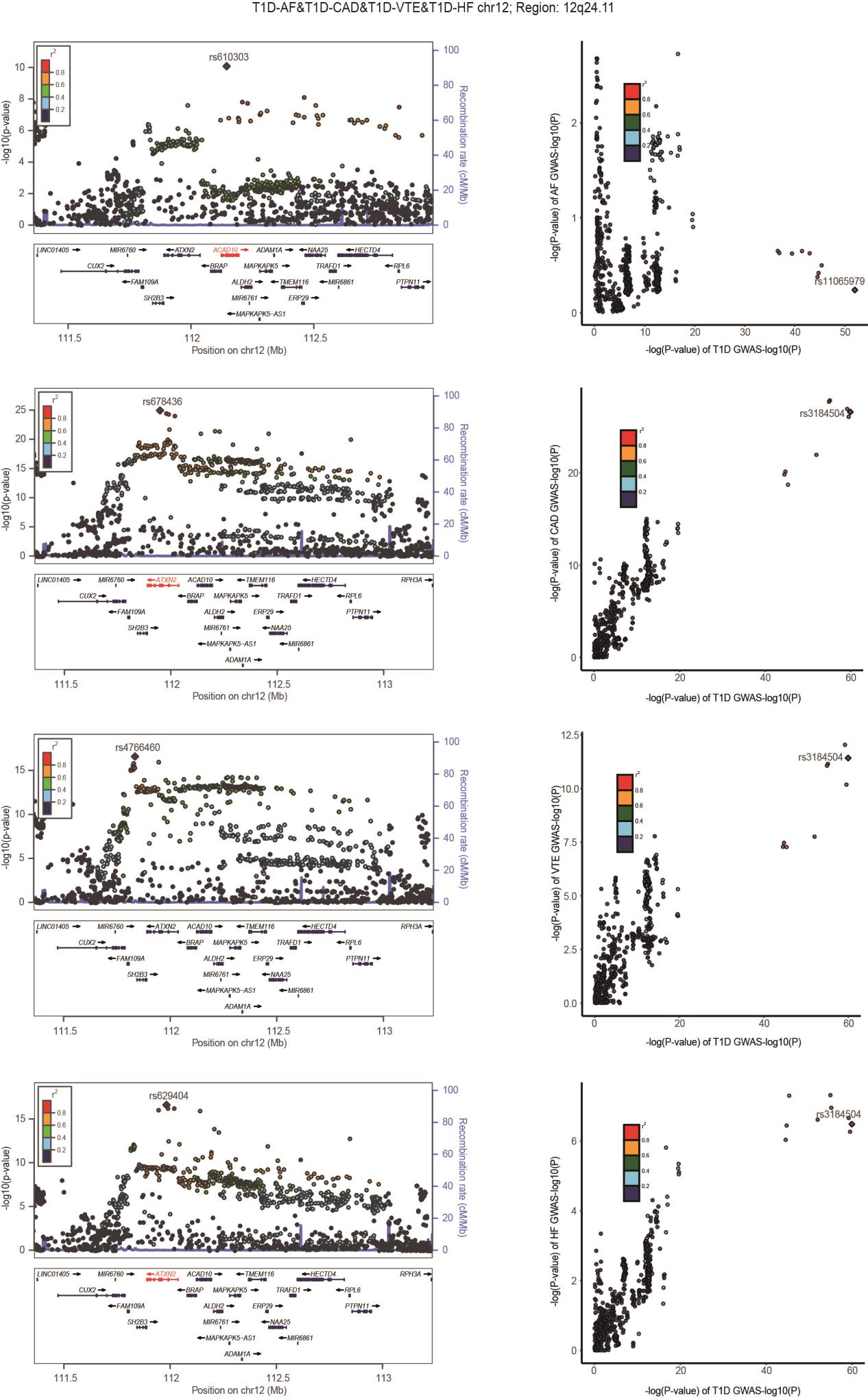

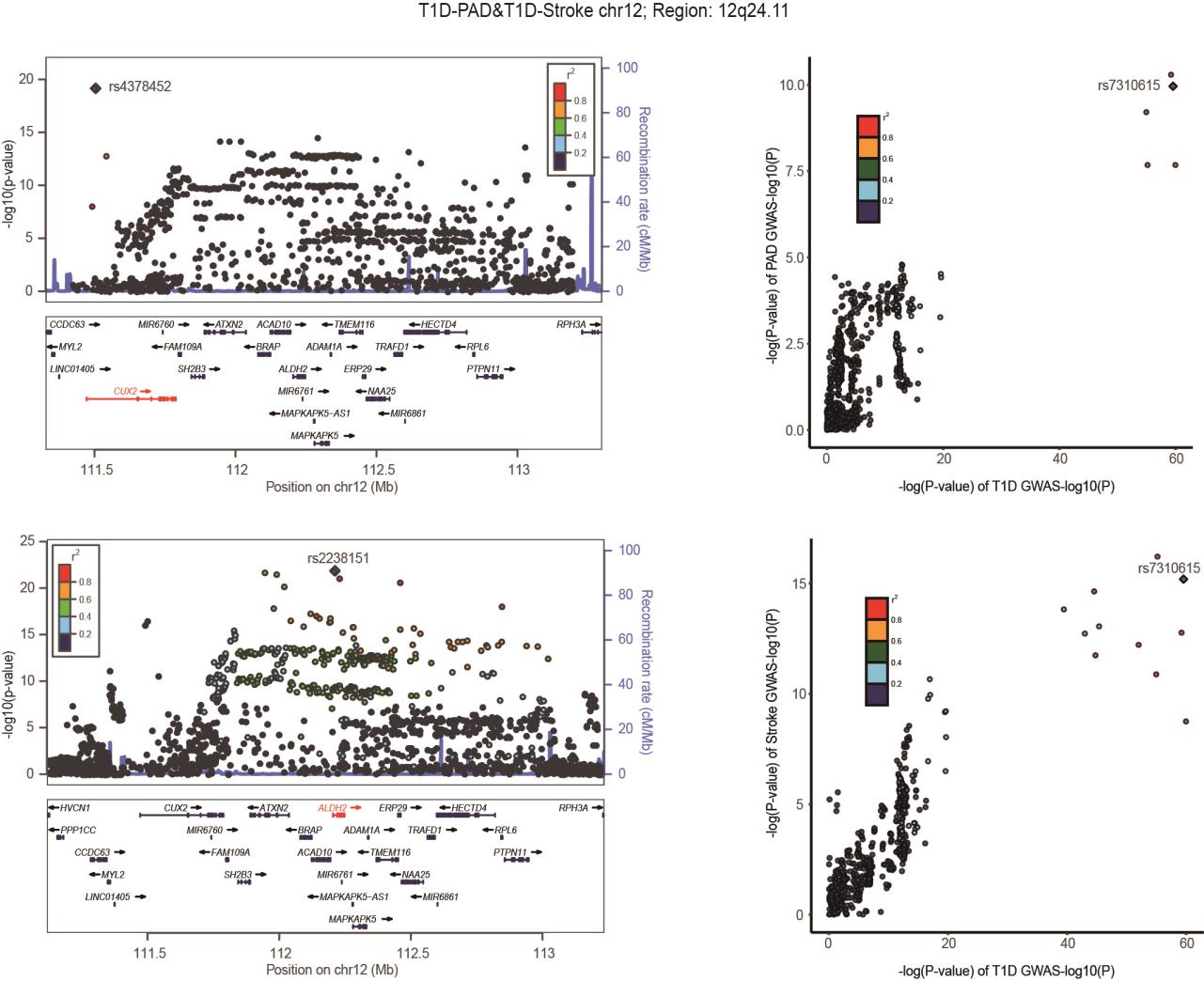

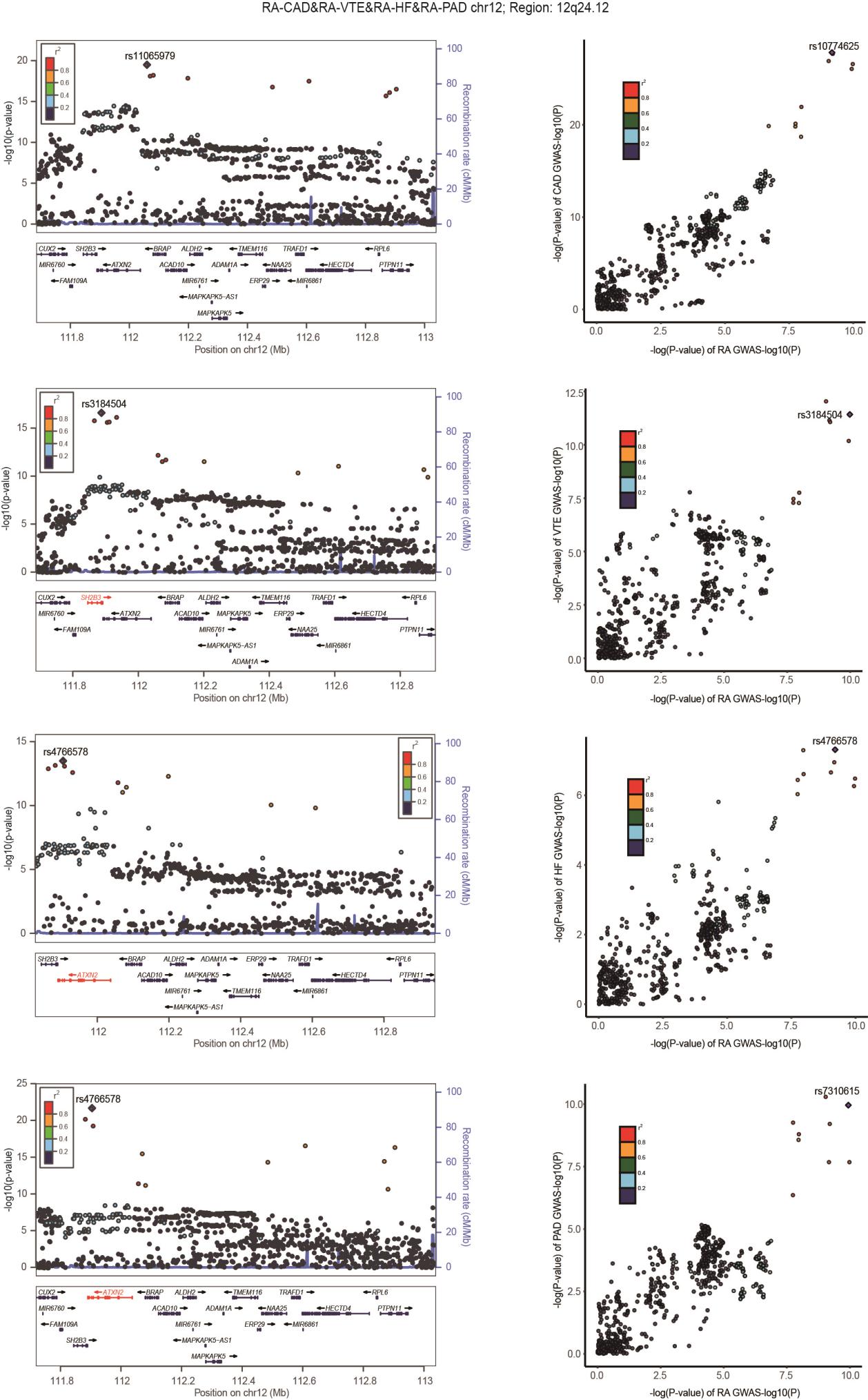

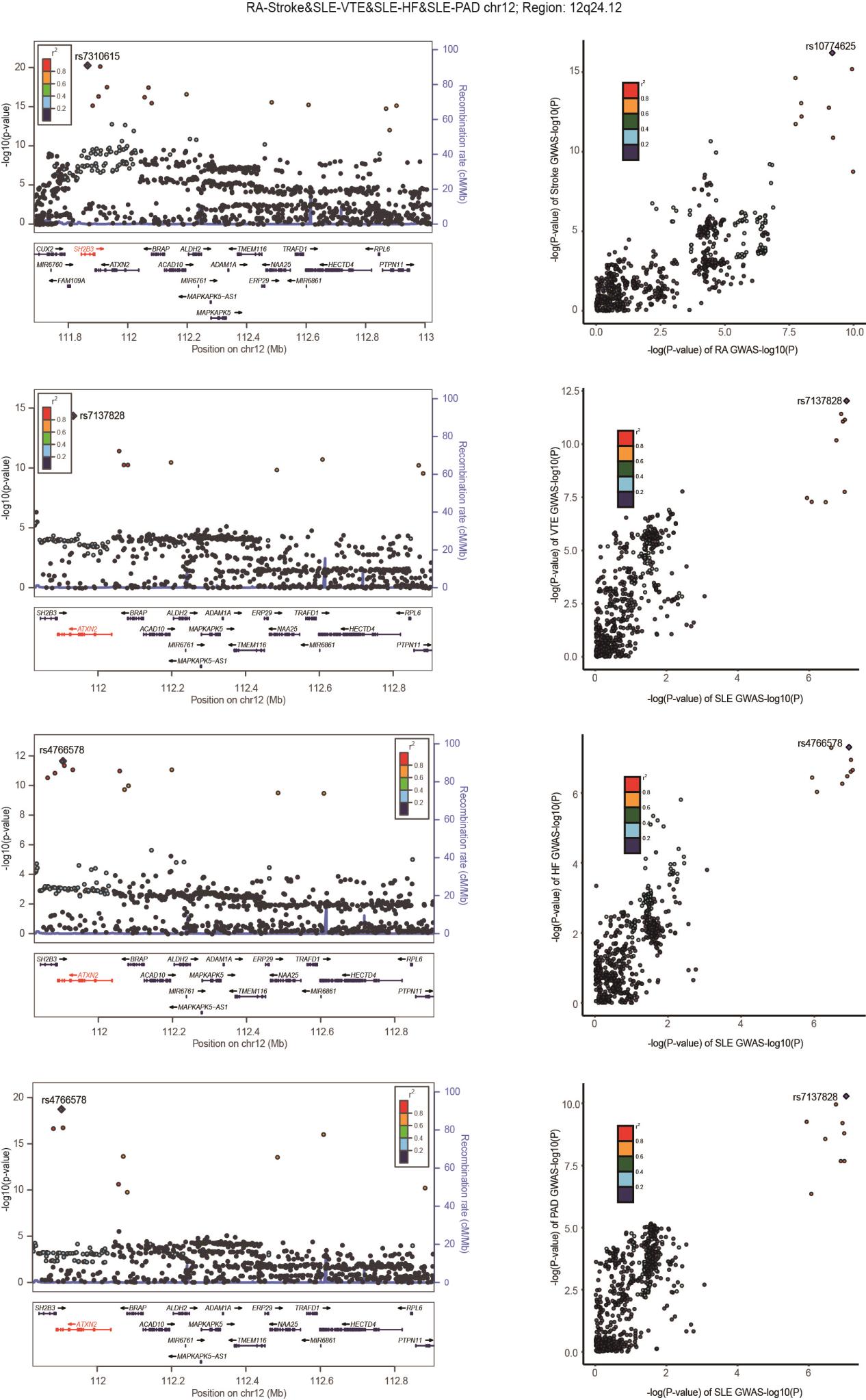

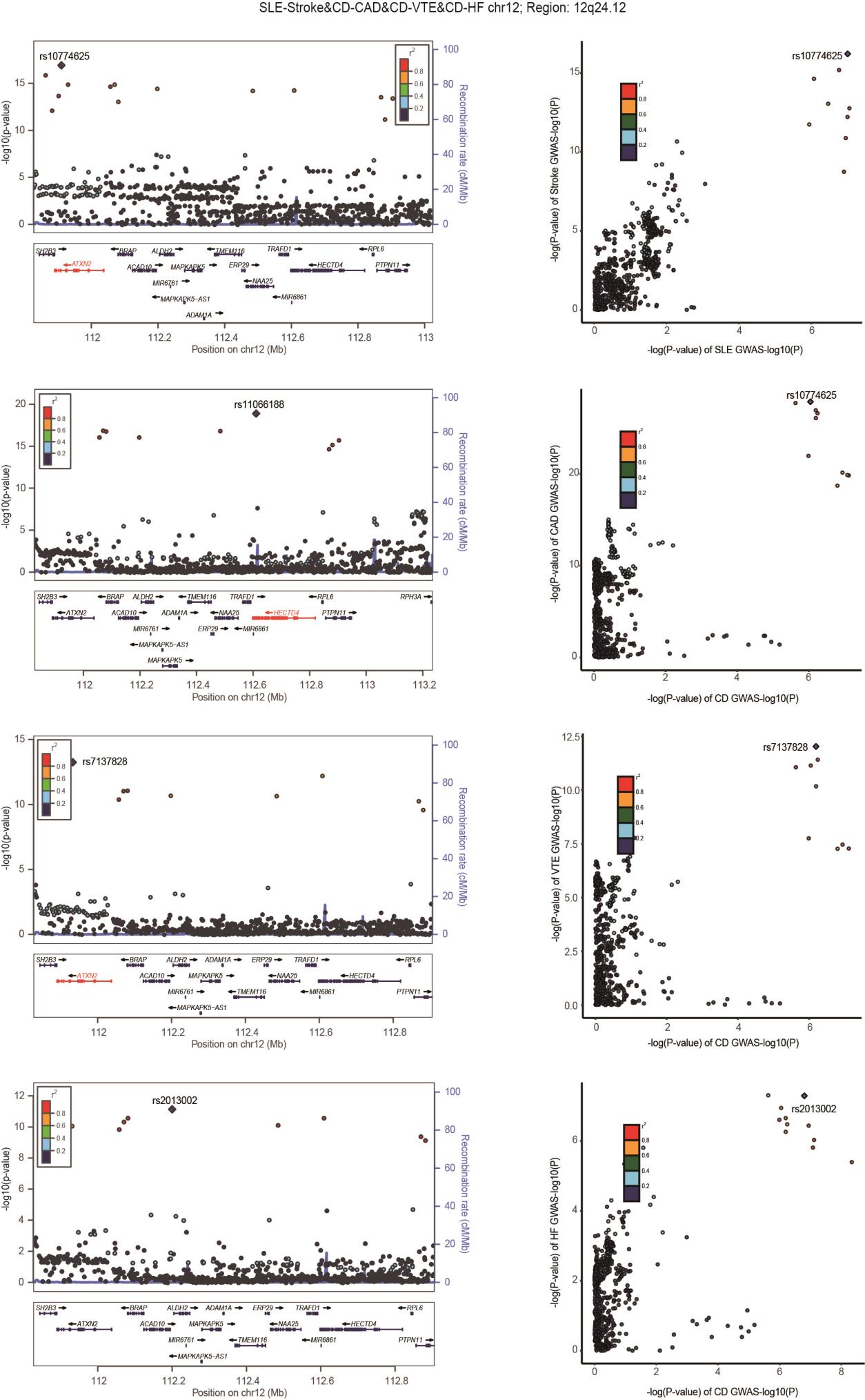

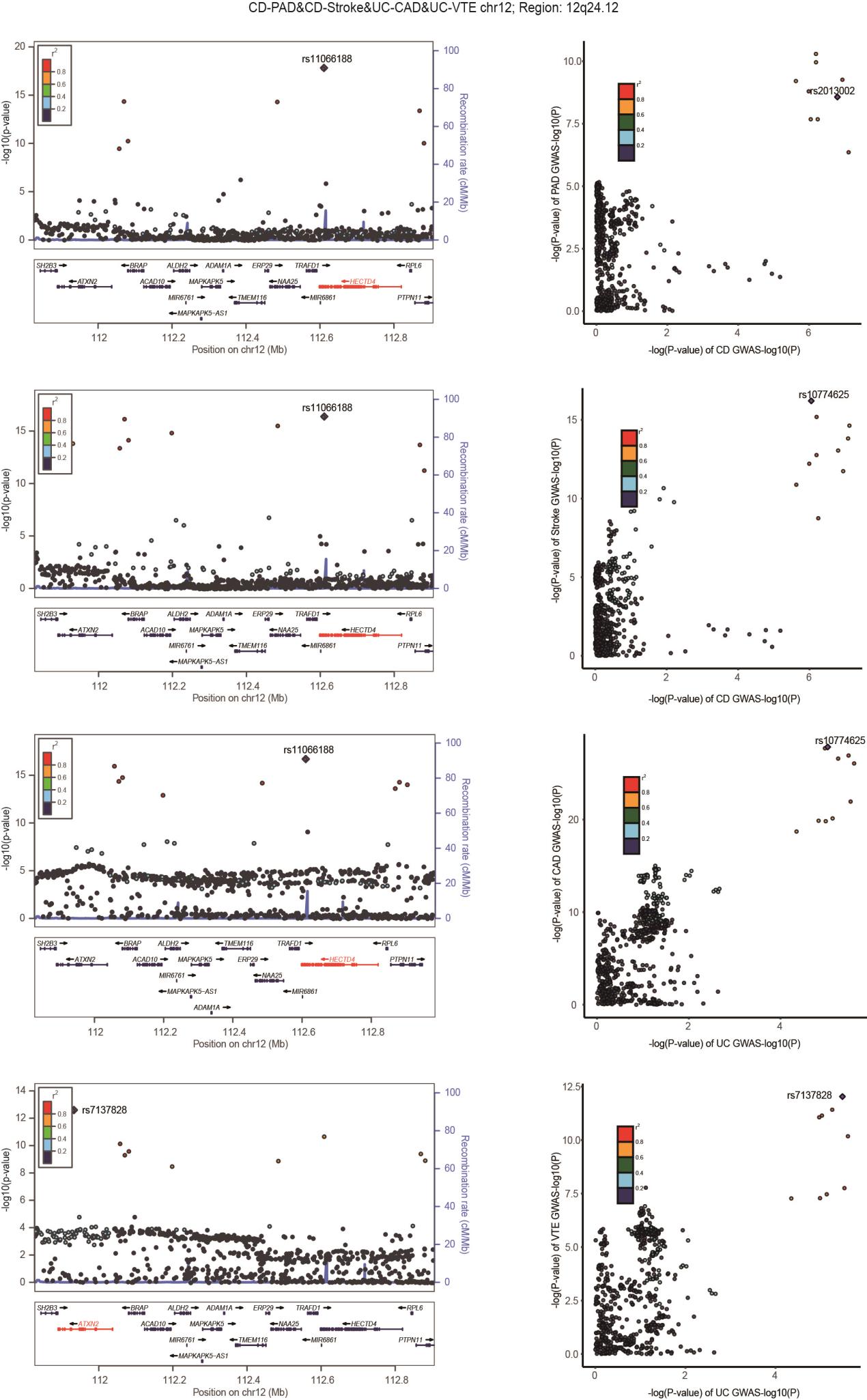

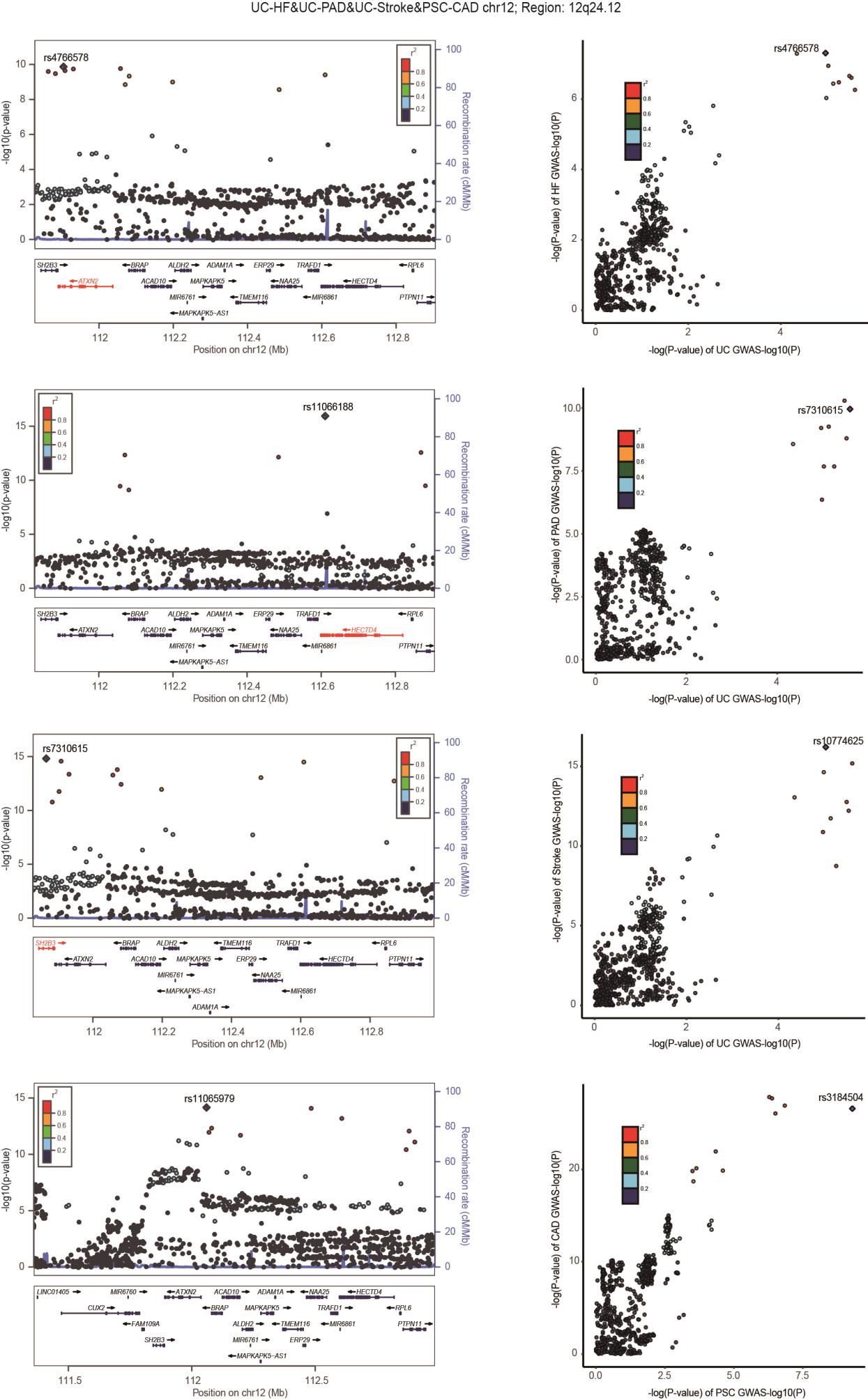

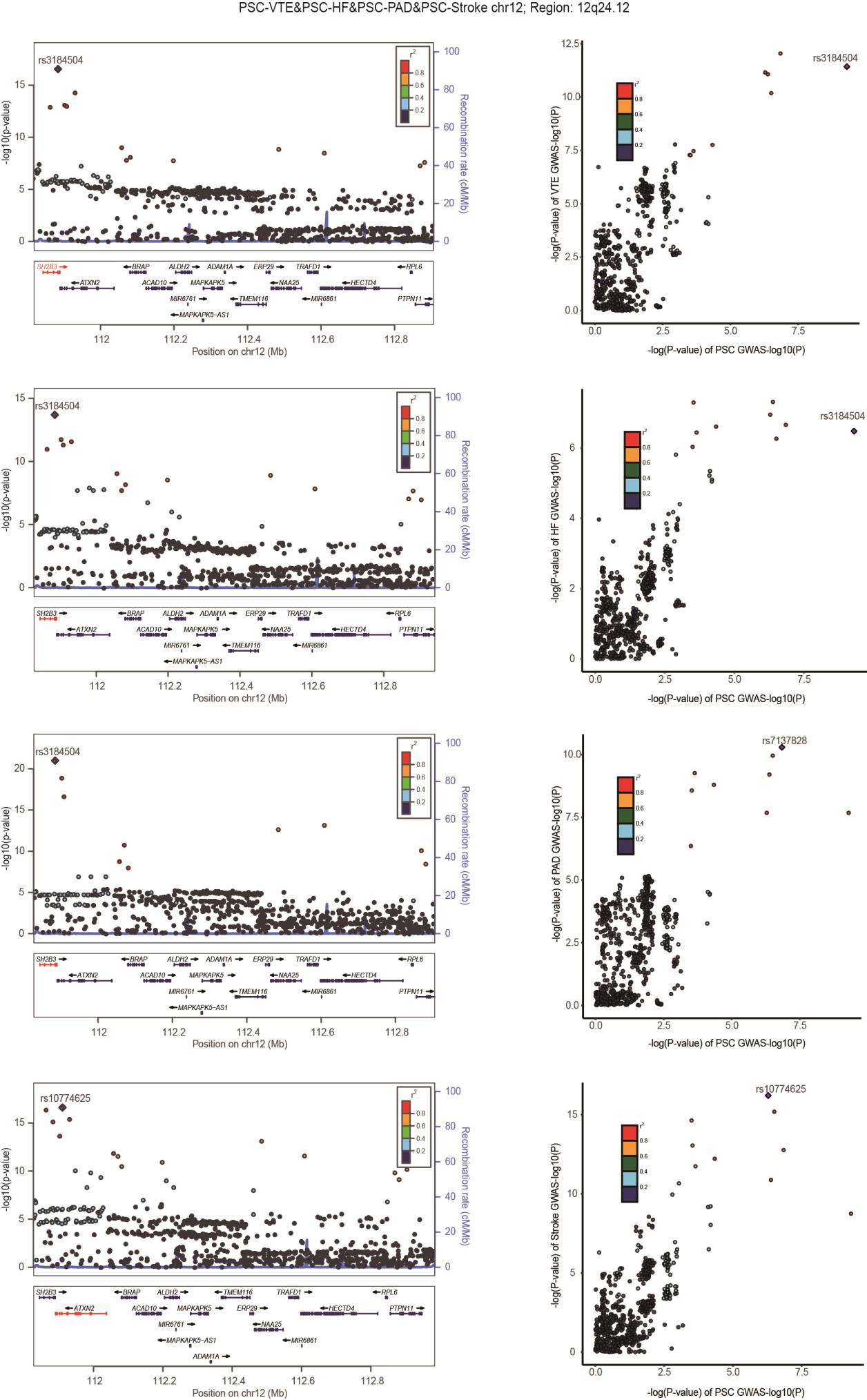

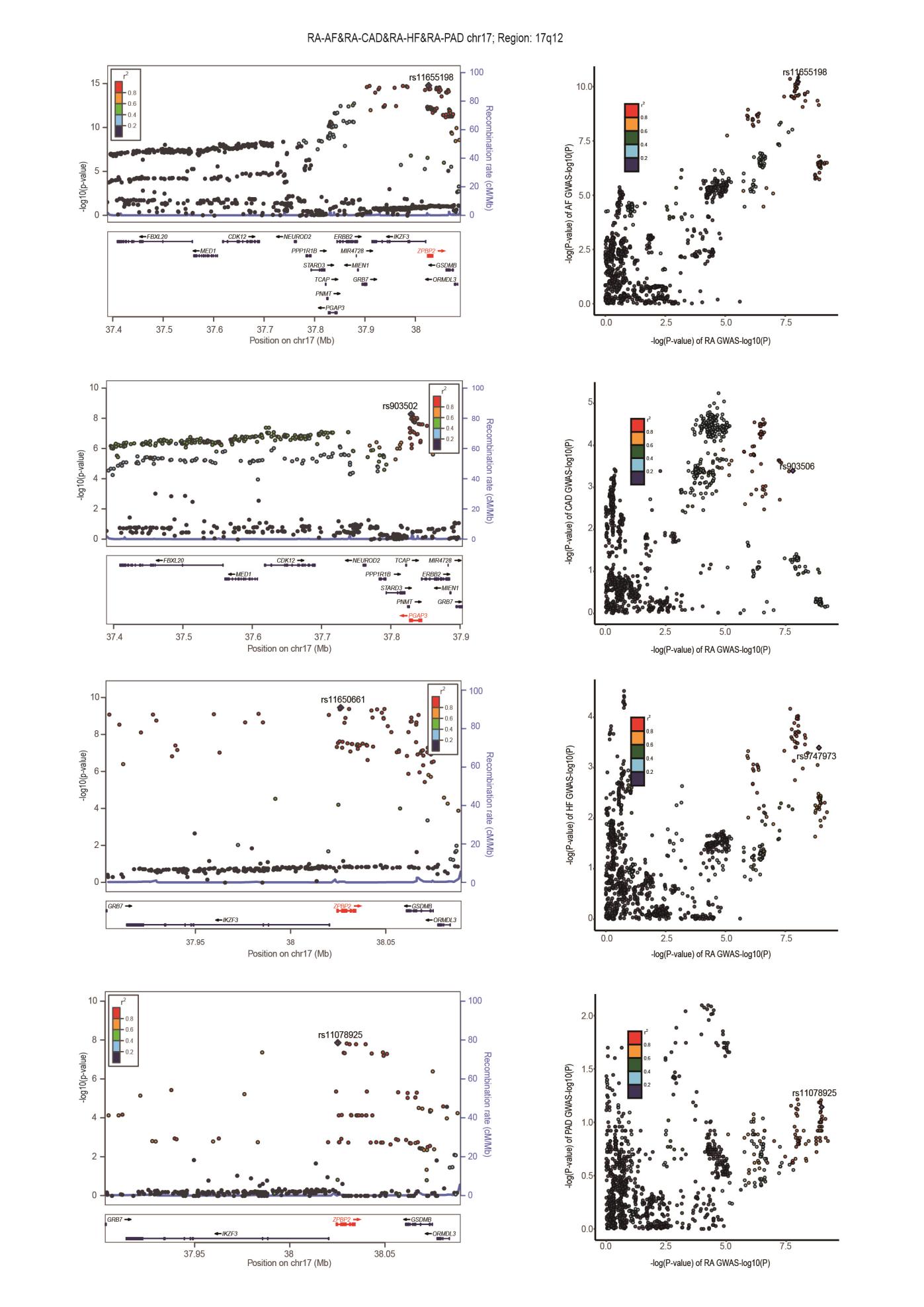

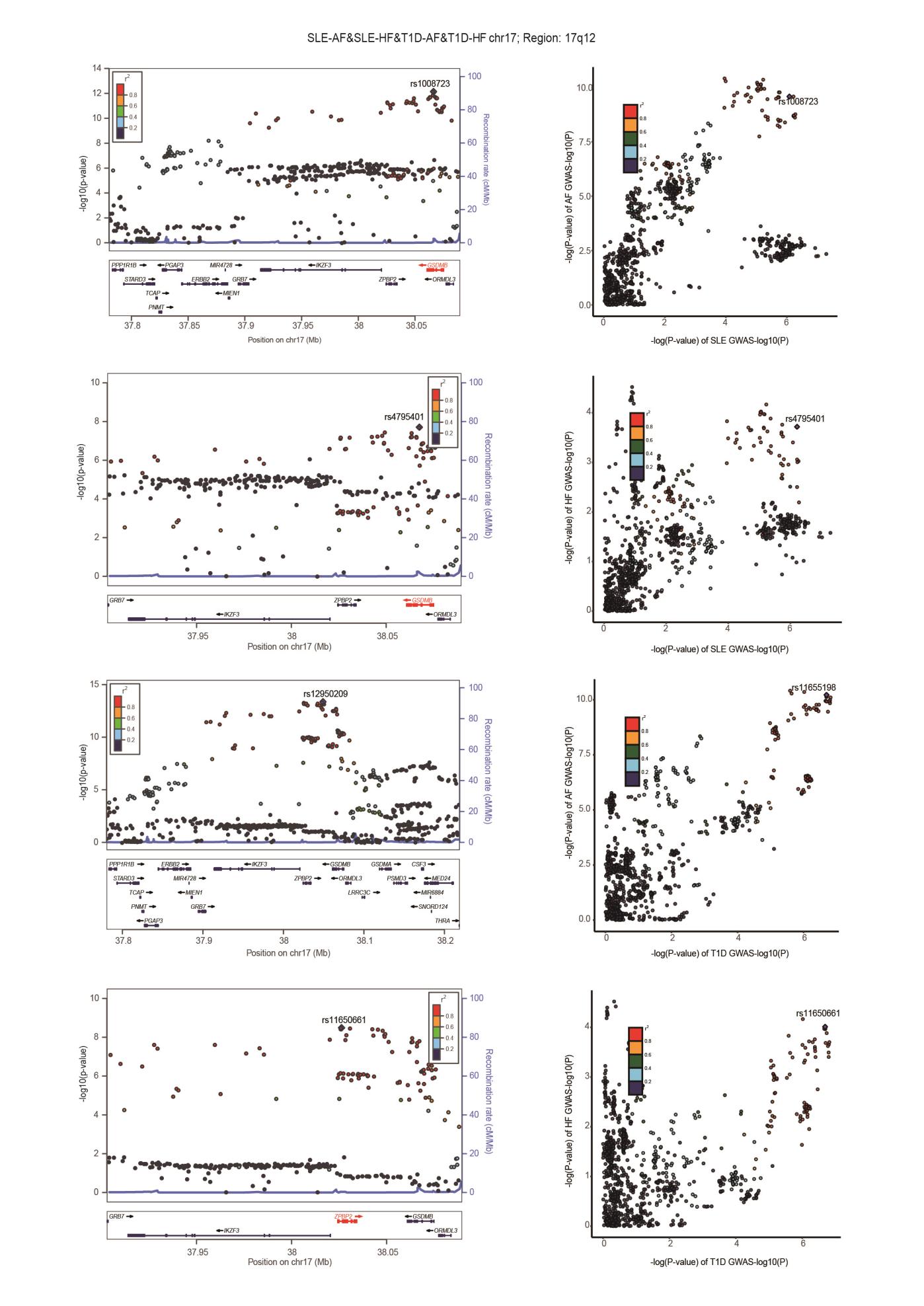

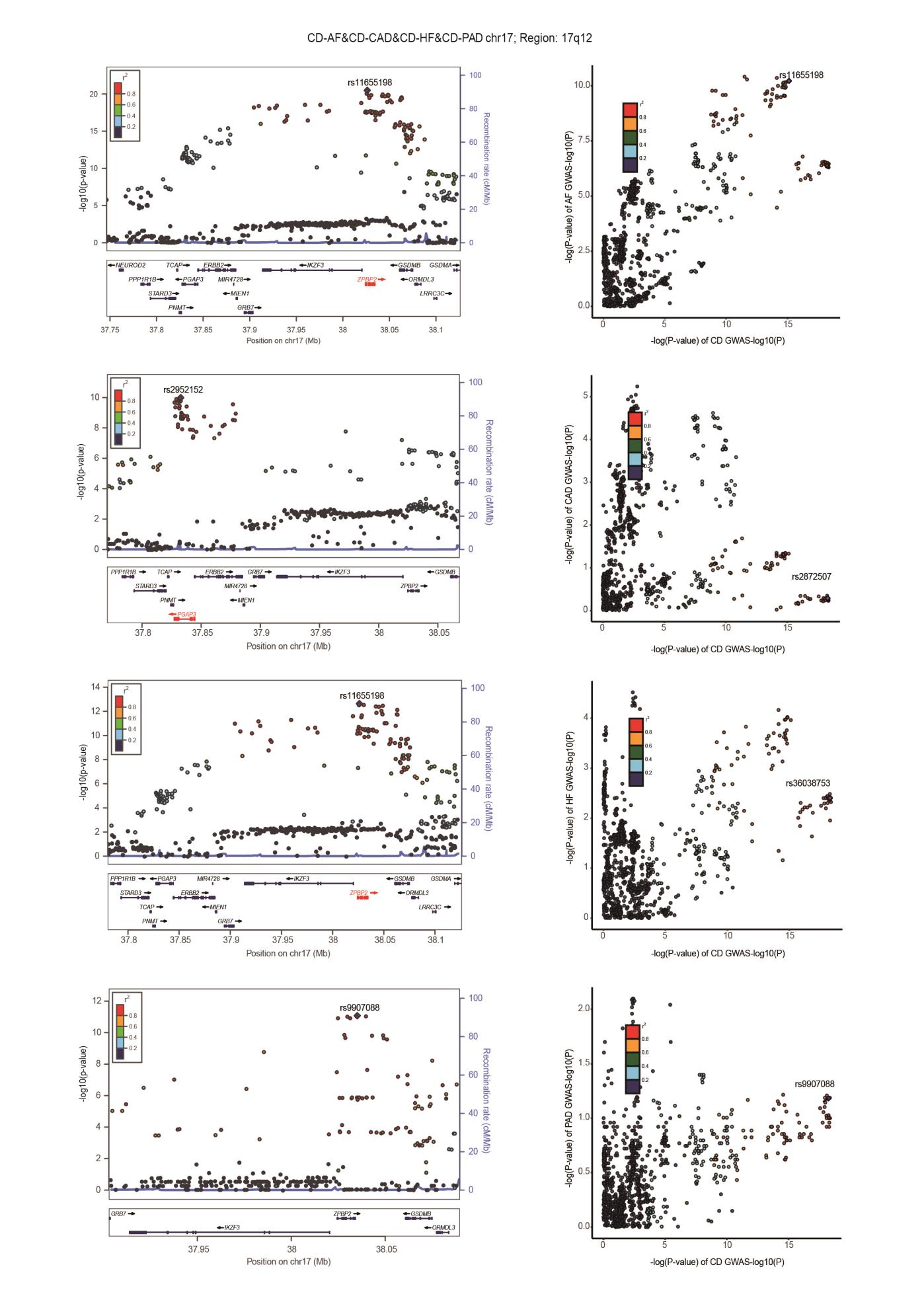

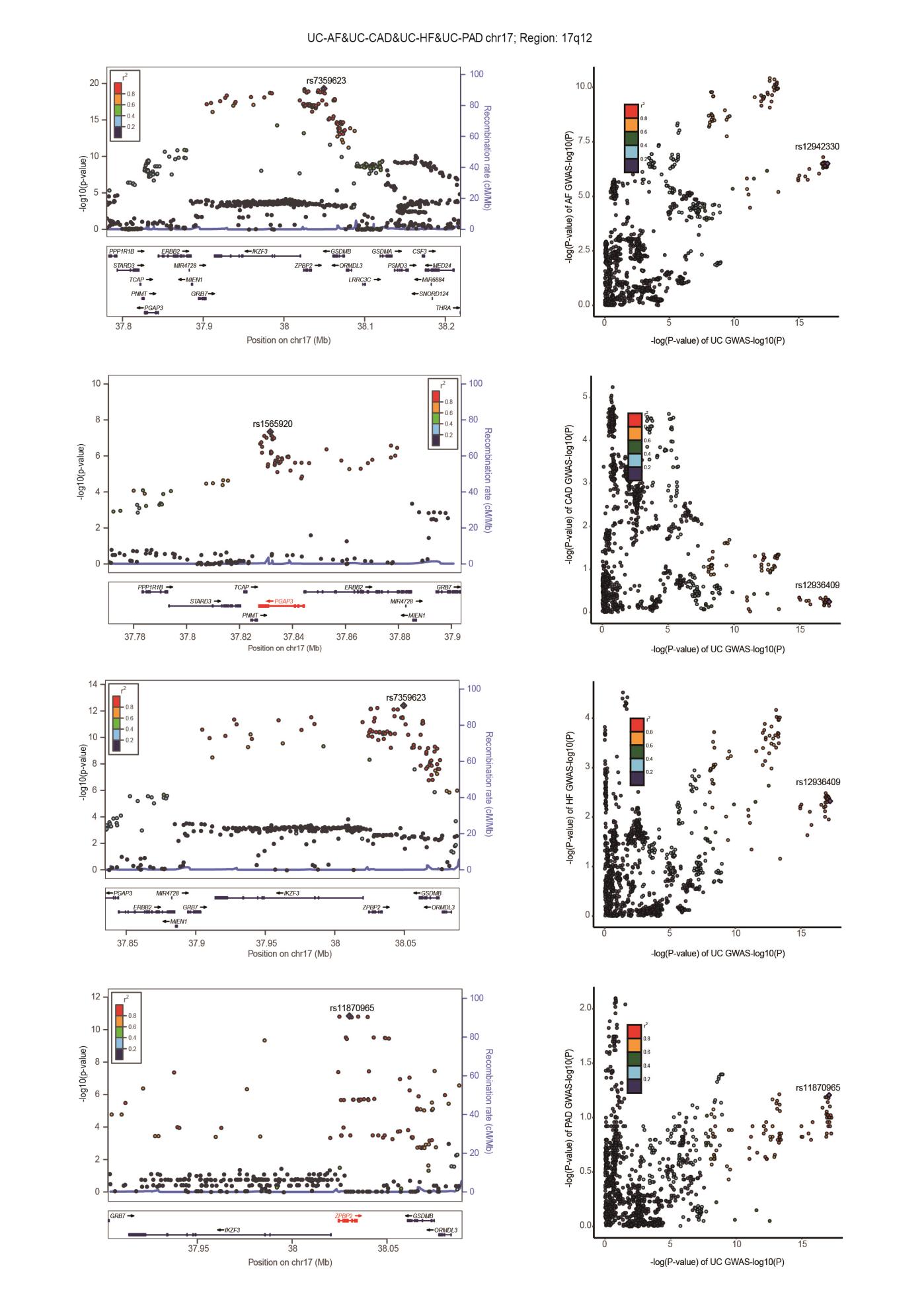

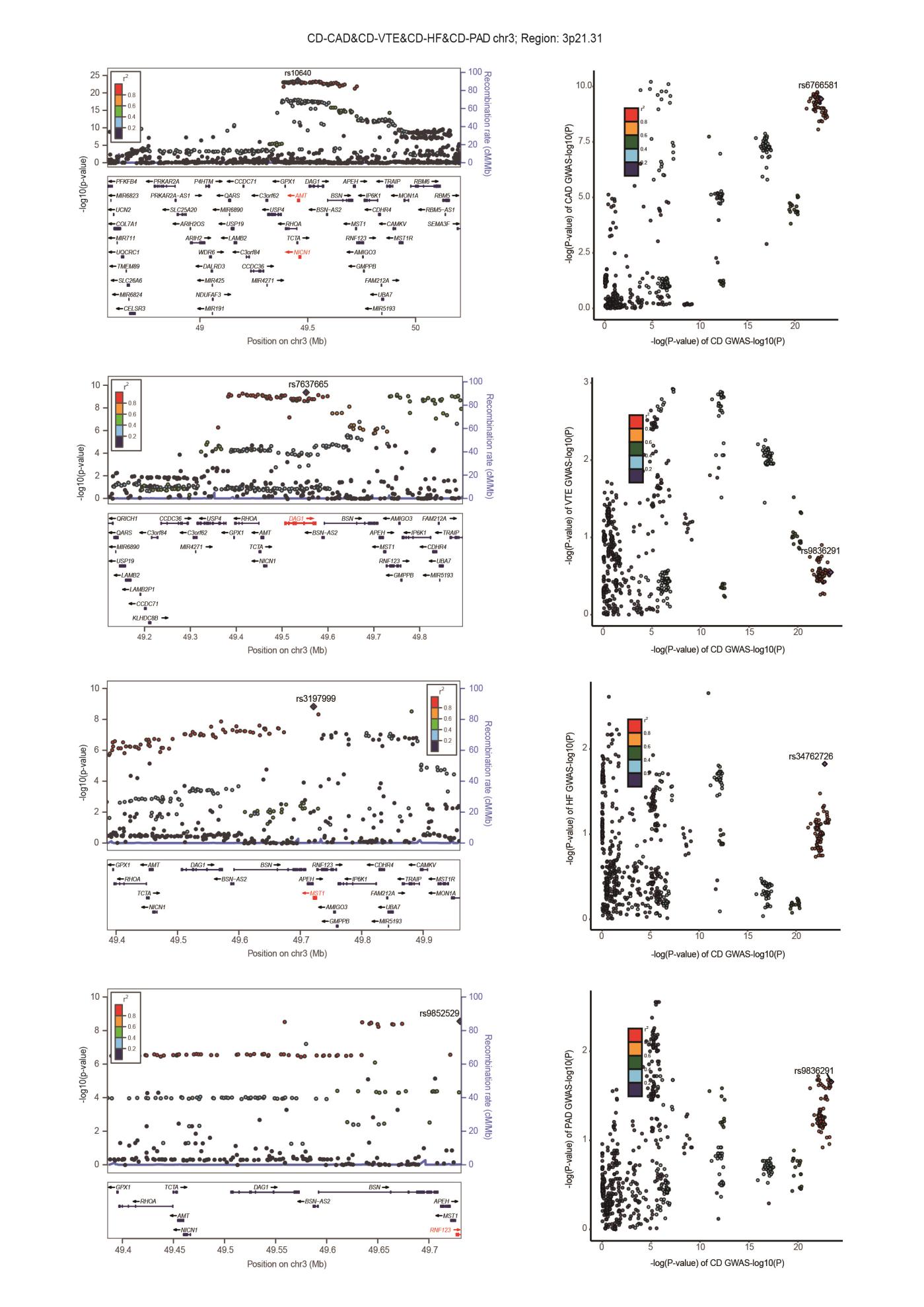

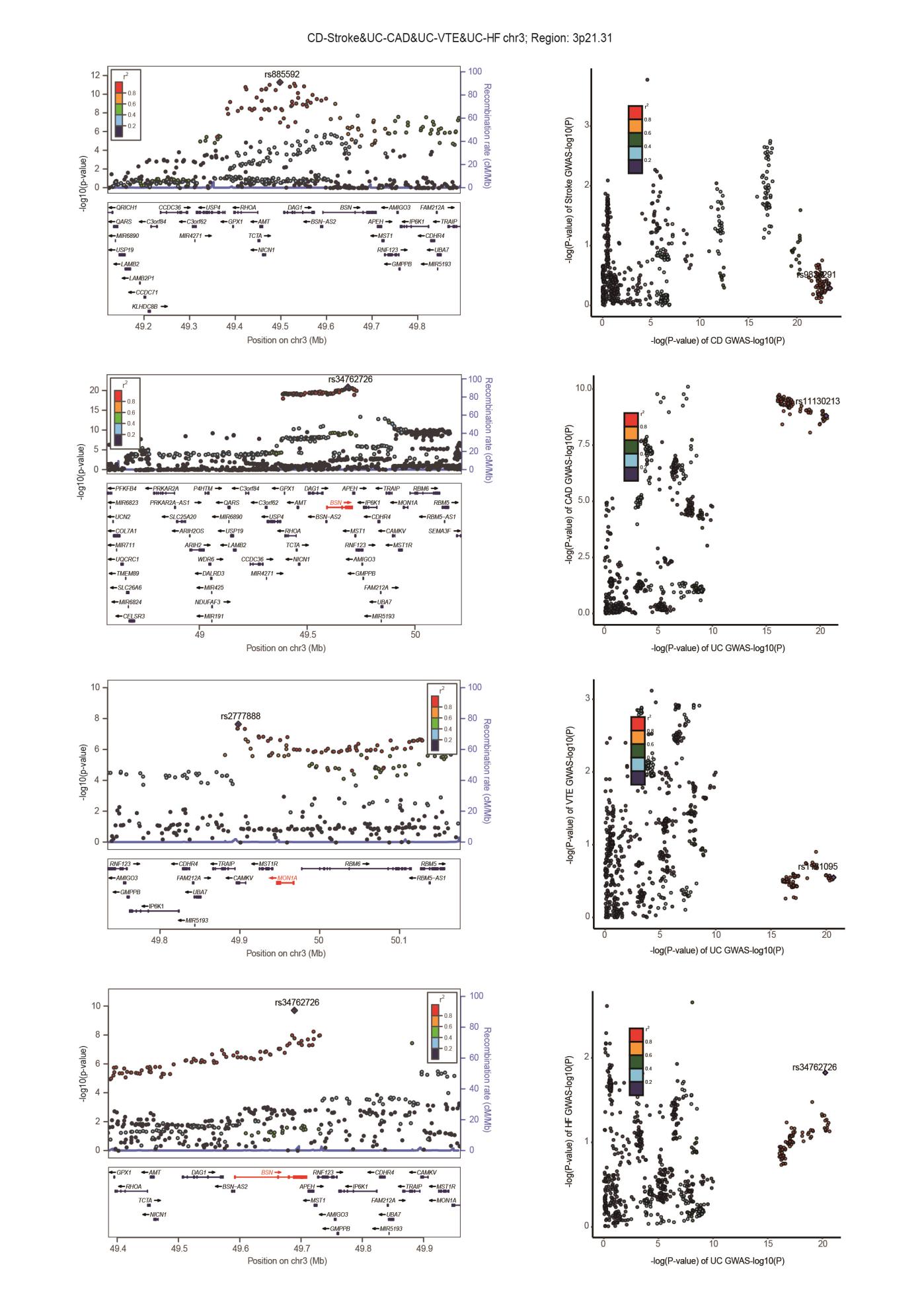

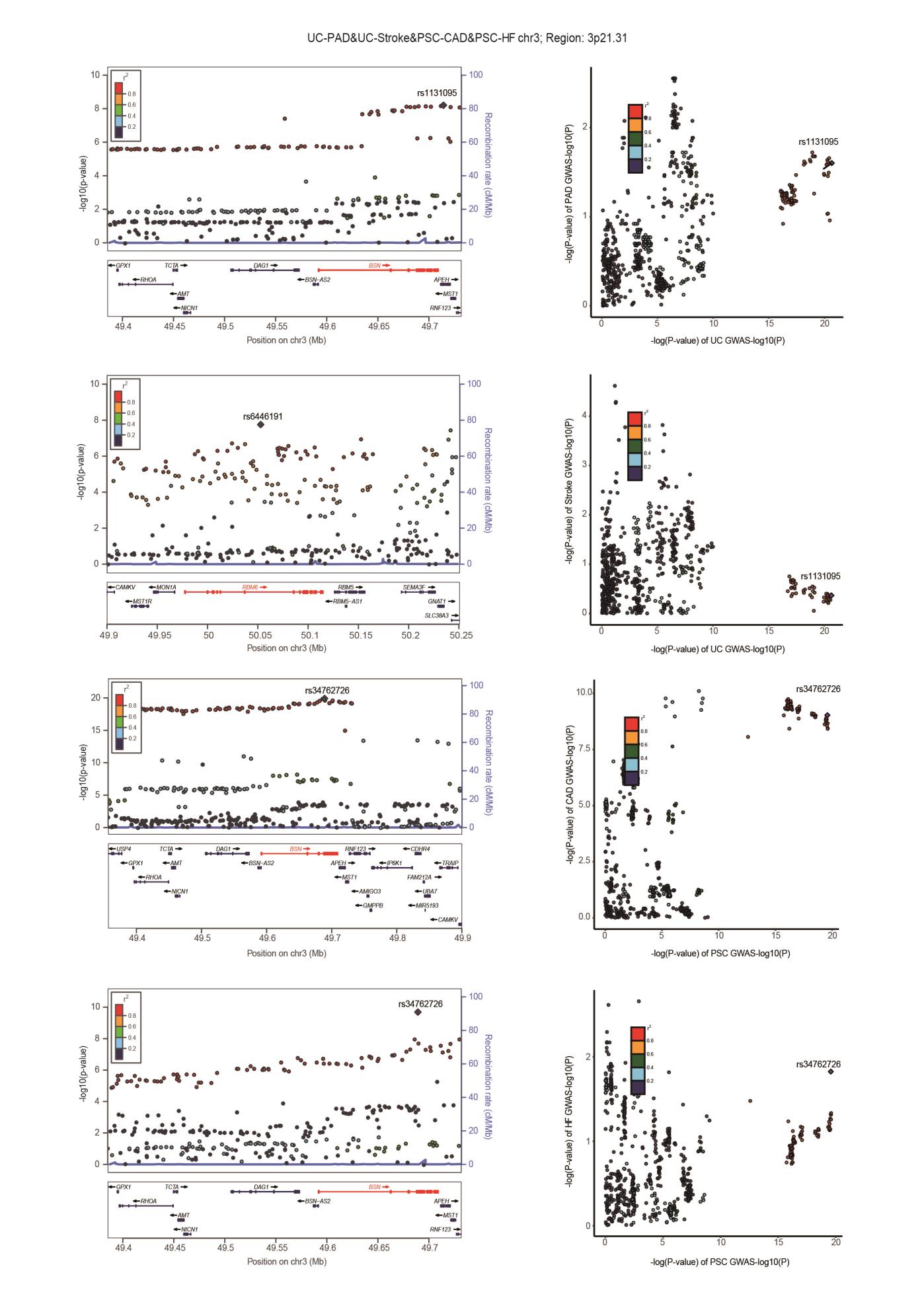

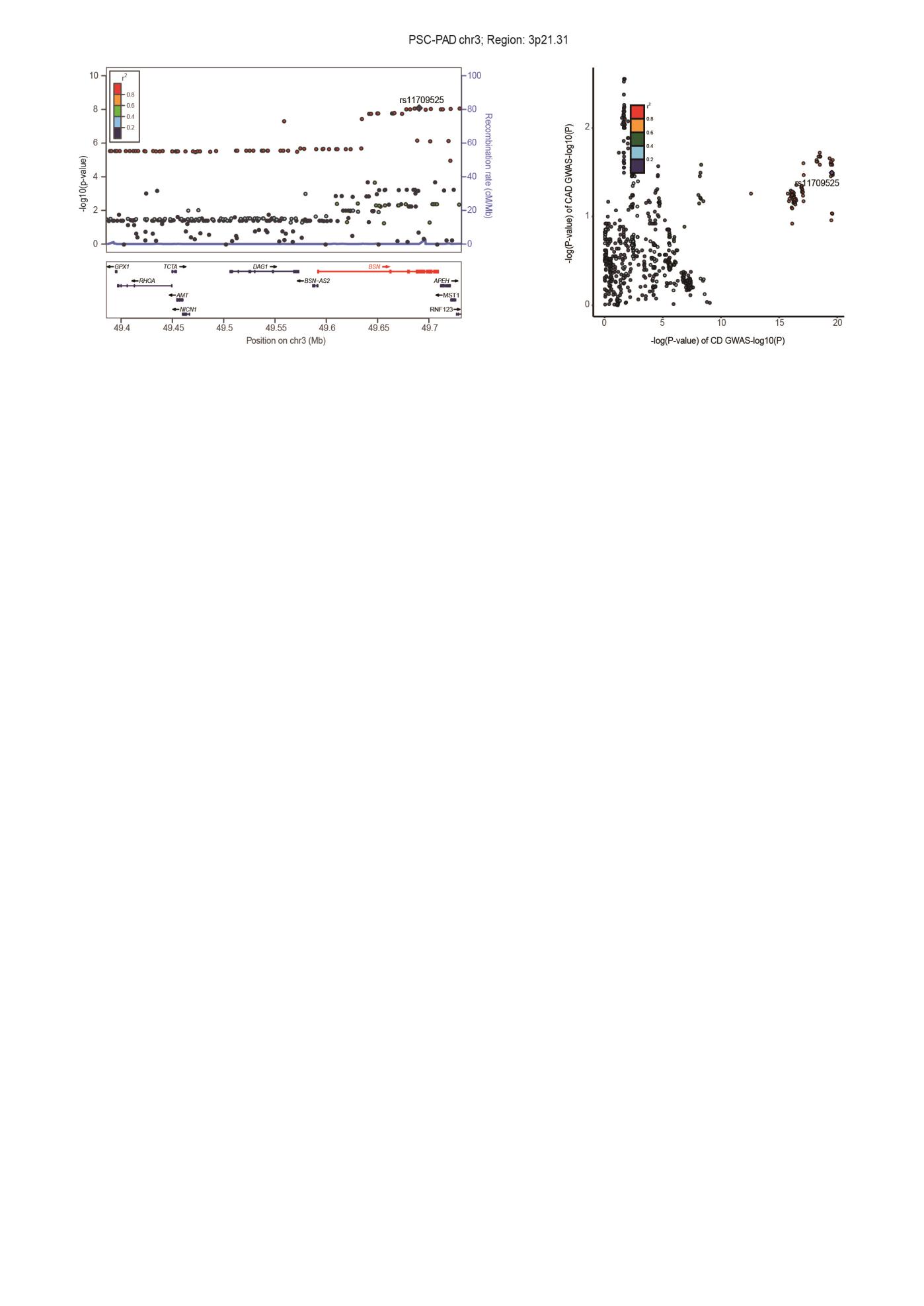

**Supplementary Fig. 4. Locus comparing plots for the shared causal variant for the associations of six autoimmune diseases and six cardiovascular diseases.**

The left panel depicts the PLACO results using a LocusZoom plot, and the right panel compares two single-trait GWAS statistics of the corresponding trait pair for each variant using LocusCompare plot. Here, we highlight the genomic loci 12q24.11-q24.12, 17q12, and 3p21.31, which influence ADs and CVDs through shared SNPs. For the LocusZoom plot, the x-axis shows the genomic position for each variant, and the y-axis shows -log10 P values from PLCAO results. The top variant with the smallest PPLACO in each locus is indicated in purple diamond. The color of each variant represents its LD relationship with the top variant. For the LocusCompare plot, each dot represents a variant; the x-axis shows the -log10 PGWAS from the corresponding GWAS of LTL, and the y-axis shows -log_10_ PGWAS from the corresponding GWAS trait. A purple diamond also indicates the candidate-shared causal variant identified by pairwise colocalization analysis. The color of each variant represents its LD relationship with the candidate-shared causal variant. All genomic locations are based on reference genome hg19, and the LD calculation is based on the 1000 Genomes Project of the European population. Detailed descriptions were provided in Supplementary Table 7. Rheumatoid arthritis, RA; Systemic lupus erythematosus, SLE; Type 1 diabetes, T1D; Crohn's disease, CD; Ulcerative colitis, UC; Primary sclerosing cholangitis, PSC; AF, Atrial fibrillation; CAD, Coronary artery disease; VTE, Venous thromboembolism; HF, Heart failure; PAD, Peripheral artery disease.
